# Forecasting the trajectory of the COVID-19 pandemic into 2023 under plausible variant and intervention scenarios: a global modelling study

**DOI:** 10.1101/2023.03.07.23286952

**Authors:** COVID-19 Forecasting Team, Robert C. Reiner, James K Collins, Chris J.L. Murray

## Abstract

**Background:** The recent Omicron-related waves of the COVID-19 pandemic have resulted in unprecedented levels of population transmission due to the variant’s high level of infectiousness across most of the world. China, the last large country to end its “zero-COVID” policies, is currently facing its own massive Omicron-related wave, and the final impact of that wave remains uncertain. We have seen repeatedly that the epidemiological characteristics of new variants can have profound impacts on global health outcomes. While the characteristics of these new variants are difficult to predict ahead of their emergence, considering the impact of potential future scenarios is of central importance for prudent planning and policy making. This paper samples across a range of potential variant-level characteristics to provide global forecasts of infections, hospitalisations, and deaths in the face of ongoing Omicron-related transmission and waning levels of past immunity and evaluates a range of interventions that may diminish the impact of future waves.

**Methods:** We created a susceptible-exposed-infectious dynamic model that accounts for vaccine uptake and effectiveness, antiviral administration, the emergence of new variants, and waning protection from both infection- and vaccine-derived immunity. Using this model, we first estimated past infections, hospitalisations, and deaths by variant, location, and day. We used these findings to more fully understand the global progression of the COVID-19 pandemic through December 12, 2022. Second, we forecasted these same outcome measures under five potential variant emergence scenarios. Third, we evaluated three different interventions in isolation and in concert within each potential variant scenario, to assess the impact of available intervention strategies through June 30, 2023.

**Findings:** We estimated that from November 15, 2021, through December 12, 2022, there were 8.60 billion (95% uncertainty interval [UI] 6.37–11.7) SARS-CoV-2 infections, 13.1 million (10.6–16.5) hospitalisations, and 3.04 million (2.65–3.55) deaths, the majority of which were attributable to Omicron variants (98.5% [97.4–99.1] of infections, 82.6% [76.7–86.3] of hospitalisations, and 72.4% [66.4–76.0] of deaths). Compared to the pre-Omicron pandemic period from January 1, 2020, to November 15, 2021, we estimated that there were more than twice as many infections (214% [163–286]) globally from November 15, 2021, to December 12, 2022, but only 20.6% (19.8–21.4) of the estimated deaths. The massive Omicron waves and high vaccination rates in many high-income countries have together contributed to high levels of immunity against SARS-CoV-2 infection, leaving only 97.3% (96.3–98.2) of the global population with no protection as of December 1, 2022. Concurrently, however, China, where only 17.6% [5.28–34.8] of the population have ever experienced infection due to its zero-COVID policy, requires special attention over the next few months, as all our future scenarios predict substantial increases in transmission, hospitalisation, and death in China in now that zero-COVID policies have been relaxed. Under the future scenario we consider most plausible (a scenario with another new Omicron-like variant emerging and reference levels of the drivers of transmission), we estimated there will be an additional 5.19 billion (3.11–7.78) infections, 13.6 million (8.50–21.8) hospitalisations, and 2.74 million (1.40–5.68) deaths between December 12, 2022, and June 30, 2023, with the Western Pacific region projected to sustain the highest rates of additional deaths, driven primarily by the uncontained outbreak in China. By comparison, a baseline scenario in which no new variant emerges results in 3.54 billion (2.24–5.43) infections, 6.26 million (4.11–9.65) hospitalisations, and 1.58 million (0.829–3.95) deaths in the same forecast period. The ability for a new variant to break through past infection- and vaccine-derived immunity greatly influences future outcomes: we estimate a new variant with the high severity of Delta, but correspondingly moderate immunity breakthrough rates will have difficulty overtaking current variants and will result in similar outcomes to the Omicron-like variant scenario with 3.64 billion (2.26–5.83) new infections, 7.87 million (4.81–13.0) new hospitalisations, and 2.87 million (1.03–5.56) new deaths. Finally, if we consider a variant that combines the high infectiousness and breakthrough rates of Omicron with the high severity of Delta, we again estimate 5.19 billion (3.11–7.78) new infections, but due to the presumed increase in severe outcomes, we estimate 30.2 million (13.4–51.2) new hospitalisations and 15.9 million (4.31–35.9) deaths over the forecasted period. The impacts of interventions vary by variant characteristics and region of the world, with increased mask usage and reimplementation of some mandates having massive impact in some regions while having less impact in others. Finally, assuming variant spread was as rapid as observed for Omicron, we find almost no impact of a rapidly developed and deployed variant-targeted booster.

**Interpretation:** As infection-derived and vaccine-conferred protection wanes, we expect infections to rise, but as most of the world’s population has some level of immunity to SARS-CoV-2 as of December 12, 2022, all but the most pessimistic forecasts in this analysis do not predict a massive global surge by June 30, 2023. Paradoxically, China, due to its lower levels of population immunity and effective vaccination will likely experience substantial numbers of infections and deaths that, due to its large population size, will adversely affect the global toll. This could be substantially mitigated by existing intervention options including masking, vaccination, health-care preparedness, and effective antiviral compounds for those at most at risk of poor outcomes. While still resulting in morbidity and mortality, this endemic transmission provides protection from less transmissible variants and particularly protects against sub-lineages of the more severe pre-Omicron variants. In the scenarios where a new variant does emerge and spread globally, however, the speed of this spread may be too fast to rely on even the most quickly developed mRNA vaccines to provide protection soon enough. Existing vaccines and boosters have played an important role in increasing immunity worldwide, but the continued contribution of mask usage (both past and future) in the prevention of infection and death cannot be understated. The characteristics of future COVID-19 variants are inherently difficult to predict, and our forecasts do show considerable differences in outcomes as a function of these variant properties. Given the uncertainty surrounding what type of variant will next emerge, the world would be wise to remain vigilant in 2023 as we move to the next phase of the COVID-19 pandemic.

**Funding:** Bill & Melinda Gates Foundation, J. Stanton, T. Gillespie, and J. and E. Nordstrom.

**Research in context:** *Evidence before this study:* Since the beginning of the COVID-19 pandemic, there have been a plethora of COVID-19 models developed; most were designed to focus on a specific location (or small set of locations) and a short time horizon (usually less than a month). A number of modelling consortiums were created to develop ensemble predictions across models of this sort (e.g., the COVID-19 Forecast Hub [maintained by the Reich Lab of the University of Massachusetts Amherst in collaboration with USA CDC (Centers for Disease Control)] or the European COVID-19 Forecast Hub [created by a multitude of infectious disease modelling teams and coordinated by ECDC (European Centre for Disease Prevention and Control)]), and the final results typically predicted four weeks, and at most, six weeks forward. The models combined for these ensembles ran the spectrum from transmission dynamic models that incorporated complex mixing patterns between individuals, to machine learning models that were agnostic of the fact that the input and output were associated with infectious diseases. Moreover, most of these models were designed to predict the most likely outcome as opposed to evaluate potential future scenarios. A small subset of these models were created with this sort of flexibility, though they have primarily been applied to limited global regions (e.g., USA CDC scenarios) and they typically do not evaluate multiple potential scenarios three to six months into the future. The Institute for Health Metrics and Evaluation (IHME) COVID-19 model has been generating and publishing forecasts of SARS-CoV-2 infections and COVID-19 deaths globally with four-month time horizons and making these available at mostly weekly intervals on its website since March 26, 2020 (https://covid19.healthdata.org). The cadence has now slowed to monthly updates as in many parts of the world, data needed to support the modelling of COVID-19 have reduced and/or ceased to be collected as the attention of policy makers and funders is drawn elsewhere. Several epidemiological scenarios have been evaluated in these online estimates, but the outcomes have not been formally compared across these scenarios globally into 2023. This article is also the first full formal documentation of the IHME-SEI model incorporating foundation work on infection–fatality ratio, more robust cumulative infection calculations, as well as more recently developed work that allows for waning immunity.

*Added value of this study:* To our knowledge, this study is the first to forecast multiple future COVID-19 scenarios of variant emergence against a background of high rates of past SARS-CoV-2 exposure globally, nationally, and for a set of subnational locations, six months into the future. It is also one of the first to forecast the impact of China relaxing its zero-COVID policy. The scenarios considered were selected to represent a range of realistic potential futures and are directly comparable by region, country, and territory (and in many instances subnational units), to identify future risk as well as inform on the effectiveness of potential intervention strategies. In particular, we directly compared scenarios where a future variant is presumed to be similar to Omicron (high infectiousness, low severity, high immune-breakthrough), Delta (moderate infectiousness, high severity, moderate immune-breakthrough), a Delta with increased immune escape (moderate infectiousness, high severity, high immune-breakthrough), or the worst of both (high infectiousness, high severity, high immune-breakthrough) to a scenario where no new variant emerges. We then evaluated several interventions against each potential variant future, each in isolation and in concert. In addition to providing timely predictions for China as they remove restrictions, we provide insight into which locations may be at highest risk for future COVID-19 infections, hospitalisations, and deaths, and what they might do to mitigate the worst possible outcomes.

*Implications of all the available evidence:* The Omicron waves have already resulted in an estimated 8.60 billion (95% uncertainty interval [UI] 6.37–11.7) infections in the past 13 months globally (from November 15, 2021, through December 12, 2022). Previous exposure to other variants and vaccination have together resulted in 97.3% (96.3–98.2) of the global population being estimated to have some immunity to SARS-CoV-2 as of December 12, 2022. While infection- and vaccine-derived immunity has and will continue to wane, this protection and ongoing transmission of currently circulating variants will mitigate the scale of the next COVID-19 wave. The scale of mitigation possible is highly dependent on the characteristics of the next variant. To assess the potential for a COVID-19 surge in early 2023, we evaluated several future variant scenarios, as well as the unlikely baseline scenario of no new variant emerging. In the absence of any new variant, our baseline model predicts 1.58 million (0.829–3.95) deaths globally between December 12, 2022, and June 30, 2023. If a variant with similar characteristics to Omicron (eg, high infectiousness and low severity) emerges on January 15, 2023, our model predicts 2.74 million (1.40–5.68) additional deaths over the same period. A variant with the characteristics of Delta is predicted to have difficulty overtaking current variants and past immunity, and despite its substantial severity, our model predicts a number of deaths similar to an Omicron-like new variant (2.87 million [1.03–5.56]). In the worst-case scenario considered, a variant with the transmission and breakthrough characteristics of Omicron and the severity of Delta would result in 15.9 million (4.31–35.9) deaths, 14.3 million (3.33–32.7) more than a scenario where no new variant emerges. In China, the potential morbidity and mortality in 2023 is high, due to a combination of pandemic history and policy that has kept levels of population immunity to COVID-19 low. In our “worst case” variant scenario, we estimate initiatives to return mask use to 80% of the population (or the location-specific current level, if higher) as well as the reimplementation of moderate mandates would avert 32.8% (18.7–51.3) of the predicted deaths, with maximal impact occurring in the European region (44.8% [28.7–61.6]). In every variant scenario, given the estimated speed of global spread, we predict that variant-targeted mRNA boosters are not able to be deployed soon enough to have a substantial impact. While there is considerable uncertainty in the future of COVID-19 variant characteristics, this study demonstrates a range of plausible outcomes expected across a spectrum of future realities. Although it would require nations to react quickly to newly detected threats, our predictions show that increased mask use and (where necessary) reimplementation of moderate social distancing mandates can mitigate much of any future challenge.

## Introduction

Since COVID-19 was declared a global pandemic by the World Health Organization (WHO) on Mar 11, 2020,^1^ evaluating and predicting future patterns in SARS-CoV-2 infections, hospitalisations, and deaths has been essential for decision-making by policy, professional, and private audiences alike. While the complex dynamics of disease transmission and human behaviours have made forecasting future patterns challenging, particularly early in the pandemic when little was known about the disease, COVID-19 projections have nonetheless been an important tool for policy makers around the world, to help inform the decisions on whether to implement different social distancing, masking, and other mandates.^2,3^ Forecasts for COVID-19 and other infectious diseases also help hospitals anticipate surges in hospital resource demands;^3–6^ help governments and corporations make decisions around regulating gatherings and closing and opening schools, offices, and other facilities;^7–9^ help policy makers and agencies distribute scarce resources such as vaccines and ventilators to high-risk areas;^10^ and help individuals assess their own behaviours and risk tolerances in a world of increasing misinformation.^11^

At the global level, estimated daily infections remained relatively steady through the spread of ancestral SARS-CoV-2 and the emergence of the first several variants of concern (VOCs; including Alpha, Beta, and Gamma),^12^ fluctuating between approximately 4.47 and 7.89 million new infections per day between mid-May 2020 and mid-March 2021 (Figure 1).^13^ Daily estimated COVID-19 death patterns varied more between the ancestral phrase of the pandemic and the “early-variants” phase (pre-Delta) than did daily estimated infections, however, with particularly dramatic fluctuations in the WHO regions of the Americas, Africa, and Europe (Figure 1). During this early-variants phase of the pandemic, daily estimated deaths hit a then-peak of more than 32,500 deaths per day in late January 2021, compared to approximately 21,600 deaths per day in early August 2020, before the Alpha variant was first detected (https://covid19.healthdata.org/global?view=daily-deaths&tab=trend). Likewise, with the emergence of new VOCs with more varied epidemiological characteristics than earlier variants—including unpredictable timing of emergence, transmissibility, and severity^14^—forecasting efforts have become even more complicated. The Delta variant was first detected in late 2020,^12^ and by mid-April 2021, the Delta surge in India had driven estimated daily global infections to a then-peak of approximately 16.5 million (Figure 1).^13^ Global estimated daily deaths subsequently peaked in early May 2021, at nearly 60 000 per day (https://covid19.healthdata.org/global?view=daily-deaths&tab=trend). During the pre-Omicron period of the pandemic, many countries experienced multiple large COVID-19 outbreaks, though several remained committed to and were able to maintain a “zero-COVID” approach (Australia, Iceland, Japan, New Zealand, South Korea, China, and several others).^13,15–17^ Over this period, vaccine distribution in many high-income countries became constrained by factors including hesitancy, while other countries were still constrained by inequitable distribution and other supply issues.^18, 19^

**Figure 1.**
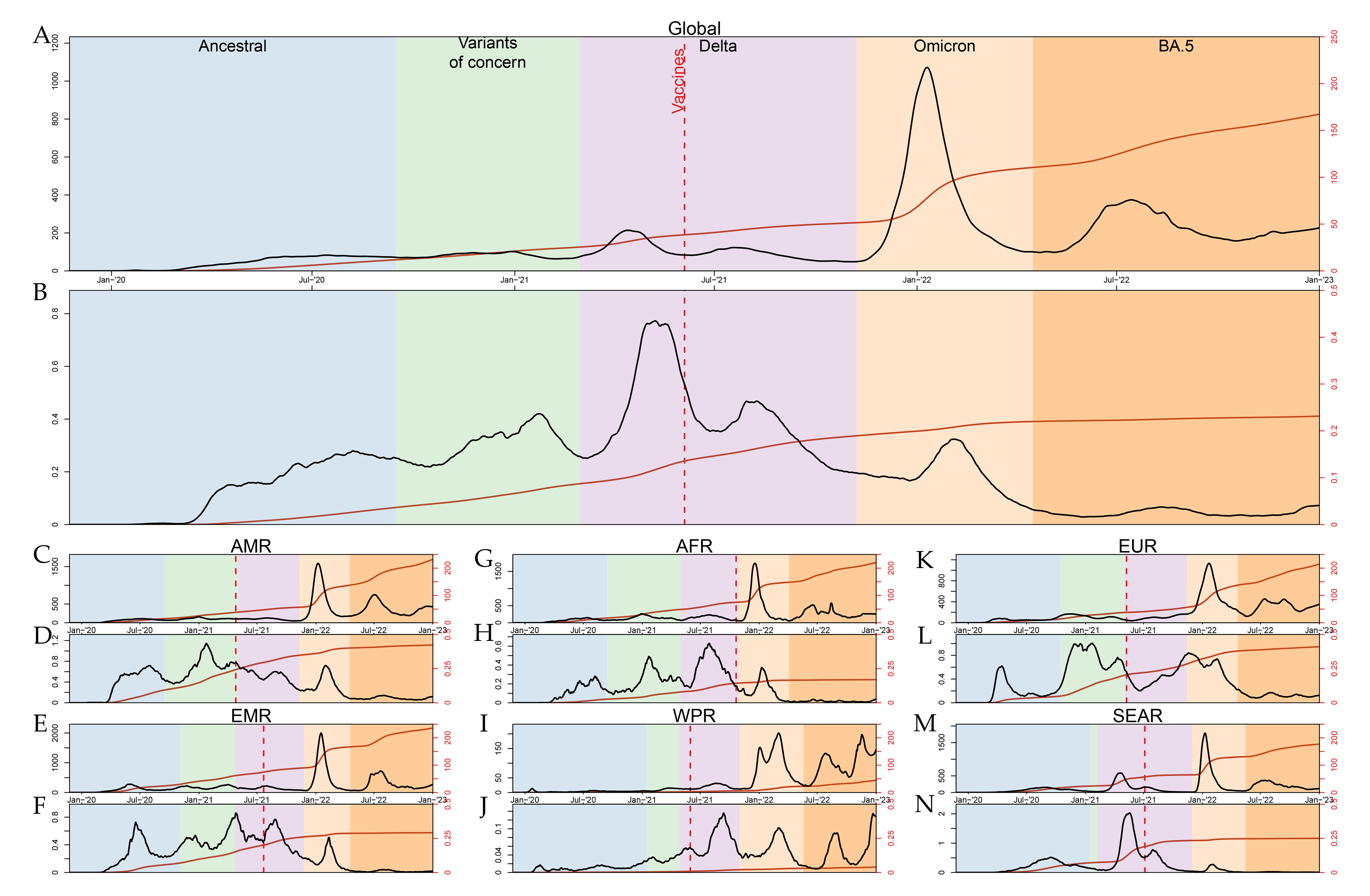
Per capita daily (black) and cumulative (brown) SARS-CoV-2 infections (A) and COVID-19 deaths (B) globally, and by WHO region (C–N) For each set of panels, by location, variant era is defined by first date when 5% of all variants were (i) Alpha, Beta, or Gamma variants of concern (green); (ii) Delta (pink); or (iii) Omicron (orange). The vertical dashed red line corresponds to the timing when 5% of the population of the corresponding panel location has been vaccinated. Both initial infections and reinfections are counted toward the total rate of infections. AMR = Region of the Americas. AFR = African Region. EMR = Eastern Mediterranean Region. EUR = European Region. SEAR = South-East Asia Region. WPR = Western Pacific Region.

The Omicron variant wave, which was first detected in November 2021,^12^ while many countries were still experiencing a Delta outbreak and dealing with the high severity associated with that variant, fundamentally changed the global transmission landscape. The previous peak of 16.5 million estimated infections per day during the Delta wave was dwarfed by a peak of nearly 83.0 million estimated cases per day at the height of Omicron (early January 2022) (https://covid19.healthdata.org/global?view=infections-testing&tab=trend&test=infections). As the first Omicron wave ebbed, countries reacted to the presumed increase in immunity by removing restrictions that have served to protect the community from SARS-CoV-2 infection and associated COVID-19 death. This policy relaxation, coupled with the emergence of newer sub-lineages of Omicron (particularly the BA.5 variant), resulted in additional waves. While most countries globally have experienced an Omicron or Omicron-related wave, the magnitude to which this wave has augmented immunity is heterogeneous, in part because the baseline immunity of countries was so varied before Omicron emerged.

New Omicron sub-variants continue to be associated with new cases and deaths, but given the general relaxation of mandates, it is difficult to parse the relative roles of behaviour and (the removal of) interventions on these smaller, localised outbreaks.^8^ In the past, mutations have led to variants that can break through past immunity, but fortunately, the severity of the subsequent infections has generally been lessened.^20–22^ The epidemiological characteristics (as well as the timing of emergence) of each of the five main variants (Alpha, Beta, Gamma, Delta, and Omicron) have been almost impossible to predict.^14,23^ While past immunity will likely dampen the impact of the next variant, given differential timings of past waves as well as differential vaccine efficacy and distribution, it remains unclear which regions of the world are most vulnerable to the next variant.

Numerous infectious disease and other modelling teams have produced COVID-19 forecasts throughout the pandemic, with wide-ranging approaches, data sources, assumptions, timeframes, locations, and other variables.^24^ Most of these models have been limited in scope, focusing on a single location or small set of locations and a short timeframe under a single plausible scenario. A number of modelling consortiums including the COVID-19 Forecast^25^ and the European COVID-19 Forecast Hub^26^ collate forecasts from dozens of models into an ensemble model and produce weekly updates on forecasted infections, hospitalisations, and deaths. Ensemble models incorporate information and uncertainty from multiple forecasts and have demonstrated superior performance over single models in this and previous outbreaks.^25^ However, ensemble COVID-19 models have generally produced forecasts for only up to four to six weeks in the future, or are limited in geographical scope to a particular country or region, and most only forecast the most plausible scenario based on current patterns.

In this study, we estimate the impact of Omicron and its descendent variants on COVID-19 infections, hospitalisations, and deaths and the resulting levels of immunity as of December 12, 2022. We also evaluate five potential future realities, each tied to a different variant emergence scenario. To conduct this evaluation, we developed a new transmission model that can accommodate variant competition, waning immunity due to vaccines and past infections, and the impact of both past and ongoing government-imposed mandates as well as transmission modifiers such as mask usage. Finally, we estimate avertable burden associated with three specific interventions: providing boosters to all previously vaccinated individuals, targeted social distancing mandate reimpositions, and returning mask usage to 80% of the population in each location, or the current location-specific level, whichever is higher. For both the past and the future, we estimate infections, hospitalisations, and deaths by day, location, variant, and vaccine status. This landscape of COVID-19 futures was chosen specifically to illuminate the wide variation in outcomes that might be expected, and hence planned for, rather than trying to identify exactly what the future will be.

## Methods

### Overview of SEI transmission model

We estimated and forecasted COVID-19 outcomes using a collection of interconnected sub-models. The core model in the process was a susceptible-exposed-infectious (SEI) transmission model that accounts for vaccination, boosters, multiple infections, antiviral treatments, and the differential waning of vaccine- and infection-derived immunity against infection and severe disease. The transmission model is structured to accept three parameterisations, each of which is used at a different phase of the modelling process. The first parameterisation takes as inputs a paired epidemiological measure (infections, hospitalisations, or deaths) and estimation of the ratio (infection–detection, infection–hospitalisation, or infection–fatality, respectively) of that measure to infections among the COVID-naive and unvaccinated population. This parameterisation is used to produce measure-specific estimates of transmission intensity and infections. The second parameterisation accepts a single estimate of transmission intensity and all available information about infections, hospitalisations, and deaths. The transmission intensity estimate is derived from an average of the measure-specific estimates produced with the first model parameterisation. This second parameterisation is used to produce final historical estimates of infections, hospitalisations, and deaths and of the infection–detection ratio (IDR), infection–hospitalisation ratio (IHR), and infection–fatality ratio (IFR), for each modelled location. The third model parameterisation takes transmission intensity and the epidemiological ratios as inputs and produces cases, hospitalisations, and deaths as outputs. This last parameterisation is used to forecast the future of the pandemic. A detailed description of the transmission model can be found in appendix 1 section 6.

### Time periods, locations, age groups, and outcome measures

Our historical model begins in December 2019, when the first COVID infections emerged in the Wuhan province of China.^27^ Most of our historical analysis is focused on the Omicron era of transmission, as we have provided a detailed account of historical COVID-19 infections and outcomes prior to the Omicron variant in Barber and colleagues.^13^ Based on GISAID Initiative data,^28^ we considered the start of the Omicron wave to be November 15, 2021. Our historical model spans roughly 13 months of the Omicron period of the pandemic and ends on December 12, 2022. We then forecast 30 scenarios spanning the six-month period of December 13, 2022, through June 30, 2023.

All the outcome measures in this study—historical and projected COVID-19 infections, hospital admissions, and deaths—are estimated for all ages and sexes combined. Estimates are presented at the global and WHO regional levels, for 176 countries and territories and for 206 subnational locations in 14 of those countries and territories.

### Estimating and forecasting key model drivers

Before implementing our epidemiological SEI model, we began by estimating and forecasting several key model drivers. First, we modelled the invasion date and rate of invasion of the most prevalent SARS-CoV-2 variants using data primarily sourced from the GISAID Initiative^28^ and performed a secondary analysis to match invasion dates with reported cases, deaths, and hospital admissions (appendix 1, section 5.1). Second, using data from Our World in Data, Linksbridge, the Duke Global Health Innovation Center (https://launchandscalefaster.org/COVID-19), the COVID-19 Trends and Impact Surveys for the USA (https://delphi.cmu.edu/covidcast/survey-results/) and globally (https://covidmap.umd.edu/), and other sources detailed in appendix 1 (section 5.2), we modelled the supply, delivery, and demand of available vaccines against SARS-CoV-2 to estimate and forecast the number of full vaccine courses and booster doses delivered in each location by brand (appendix 1, section5.2). Third, we estimated brand-specific waning vaccine efficacy against each variant^29^ and used these estimates to transform vaccine delivery into transmission risk-reduction curves (appendix 1, section 5.3.2). Fourth, we estimated the waning of infection-derived immunity and protection from severe disease.^30^ Fifth, we estimated the ratio of location-specific weekly pneumonia deaths to the annual average as a proxy for seasonal trends in COVID-19 transmission based on weekly vital registration data (appendix 1, section 5.4). Sixth, using reports from government health authorities and data from Our World in Data, we estimated the per capita testing rate and forecasted its growth up to a location-specific threshold (appendix 1, section 5.5). Seventh, we compiled a comprehensive database of the application of 21 detailed NPIs (non-pharmaceutical interventions, eg, closing primary school or non-essential retail) representing six NPI categories (eg, education and business closures) from January 2020 to October 2022 that are standardised across all modelled countries and subnational units (appendix 1, section 5.7). Eighth, using survey data from the USA and Global COVID-19 Trends and Impact Surveys, the PREMISE Behavior Survey, and the YouGov COVID-19 Behaviour Tracker Survey, we estimated the percentage of the population regularly wearing masks (averaged across different mask types and settings based on survey participants’ own interpretations of what “always” wearing a mask means), and projected mask use by location (appendix 1, section5.6). Full data and modelling details for the model drivers are available in appendix 1, section 5. In addition to the model drivers we estimated, we also leveraged demographic data and several time-invariant covariates from the Global Burden of Diseases, Injuries, and Risk Factors Study (GBD)^31–33^ 2019 to better inform location-level heterogeneity in many of our sub-models (appendix 1, section 5.8).

### Estimating historical COVID-19 outcomes by location

Model drivers in hand, our next task was to estimate historical transmission intensity in each location using all available reported data. We parameterised our model of variant-specific IDR, IHR, and IFR relative to the IDR, IHR, and IFR among the infection- and vaccine-naive population experiencing an infection with ancestral-type (D614G) COVID-19. We then paired a database of bias-corrected cases, hospital admissions, and excess deaths with SARS-CoV-2 seroprevalence surveys adjusted for waning antibody sensitivity, vaccination, and escape variant reinfection. These paired data were subset to the first period of the pandemic when no vaccines or variants were present and used to produce an initial empirical estimate of the infection- and vaccine-naive IDR, IHR, and IFR using a statistical model of the ratios that incorporated several of the model drivers to fill data gaps (appendix 1, section 4). The input measure and resulting ratio from each ratio model were then run through the first parameterisation of our transmission model using all historical data, allowing us a first-pass estimate of the fraction of COVID-19 infections, hospital admissions, and deaths due to ancestral COVID-19 among the infection- and vaccine-naive population. These naive measures were then paired with the full seroprevalence dataset and run through the ratio estimation process a second time to produce a robust empirical estimate of the epidemiological ratios. The updated ratios and original measures were run again through the transmission model to produce the final measure-specific estimates of transmission intensity. The next task was to use measure-specific models to produce a single, coherent estimate of historical COVID-19 outcomes in all locations. To do so, the measure-specific estimates of transmission intensity were averaged into a single transmission intensity per location and then input with all available infection, hospitalisation, and death estimates into the second parameterisation of the transmission system. This produced our final historical estimates of infections, hospital admissions, and deaths due to all variants among all vaccine- and SARS-CoV-2-exposure population subgroups. This step also implicitly produced our final estimates of the IDR, IHR, and IFR for each location so that each was consistent with the combined estimate of transmission intensity. This model of historical outcomes is detailed in appendix 1, section 7.

### Forecasting future COVID-19 outcomes by location

The final step in our modelling process was to forecast the disease dynamics in every location. To do this, our final estimate of transmission intensity was regressed against several of the key model drivers and then forecasted based on those relationships. We also produced simple forecasts of the COVID-19- and vaccine-naive ratios assuming they would transition linearly to their prior 180-day average over the next 30 days and then hold constant. The forecast transmission intensity and ratios, future estimates of vaccine and antiviral coverage, and future dates of new variant emergence were then input into the third parameterisation of the transmission model to produce forecasts of infections, deaths, cases, and admissions. See appendix 1, section 8 for full details on these methods.

### Forecasting variant scenarios

We projected 30 scenarios in this study. These scenarios combine five potential future variant scenarios with six possible intervention responses.

Given the stochastic nature of viral evolution, it is impossible to say what kind of variant will emerge and when and where that emergence will take place. Within the bounds of that uncertainty, it is our aim to provide a set of plausible future realities in which to ground the discussion of COVID-19 interventions. The first (baseline) variant scenario introduces no new variant and allows the continued dominance of Omicron to shape the future of the COVID-19 pandemic. The remaining four scenarios all share several key characteristics. They all start in South Africa on January 15, 2022, and follow the location-to-location invasion pattern as the Omicron variant. We elected to use the Omicron invasion pattern (ie, variant is first detected in South Africa) because it allowed us to use a historically realistic location-to-location spread in what we expect is a representative behavioural and mandate policy environment (appendix 1, section 5.1.4). These scenarios differ in their parameterisation of transmission intensity and severity. The first has an Omicron-like (high infectiousness, moderate severity, high immune-breakthrough) variant emerge, while the second has a Delta-like (moderate infectiousness, high severity, moderate immune-breakthrough) variant emerge. The third scenario has a close-to-worse-case enhancement added to the Delta-like scenario (moderate infectiousness, high severity, high immune-breakthrough). The last scenario is a worst-case scenario where a variant with Omicron-like transmission intensity and breakthrough capabilities and Delta-like severity emerges.

### Forecasting intervention scenarios

The four intervention scenarios are reference, increased mask usage (to 80% of the total population, or location-specific current level, if higher than 80%), targeted mandate reimposition, and variant-specific boosters rolled out at a country-specific or regional maximum rate. Our reference scenario is reflective of historical patterns in the implementation of these interventions, and therefore reflects what we believe to be the most plausible future trajectory of the model drivers. The reference scenario predicts vaccination and booster levels based on the best available supply, manufacture, hesitancy, and effectiveness data. Our model assumes that boosters restart the recipient’s once-waning immunity (appendix 1, section 5.3). It also forecasts mask use under the assumption that use will remain constant into the future. This forecast is based on the assumption that the population risk tolerance is at a steady state relative to the current climate of high transmission and low severity variants of SARS-CoV-2 circulating (appendix 1, section 5.6.2). This assumption is not specific to a particular mask type (eg, cloth, surgical, or N95) or setting (eg, indoor or universal), and instead assumes that the people who continue to wear a mask will do so under the same parameters they set in the past. Finally, it assumes mandate levels will likewise remain constant into the future under the same assumption of steady-state risk tolerance.

The increased mask use scenario has mask use increase linearly over the first week of the projected period to 80% of the population for each location (or the current level, if higher). This scenario assumes that the population that reverts back to mask wearing does so in the same manner as when their location was at peak mask use, eg, if they wore cloth masks previously and only wore a mask indoors, they would again wear cloth masks and only indoors; likewise, if they wore N95s both indoors and out, they would do so again.

The targeted mandate reimposition scenario has countries and subnational units reimpose social distancing mandates at location-specific levels based on location-specific thresholds of daily reported deaths (described in appendix 1, section 5.7.2).

The variant-specific booster scenario introduces a new vaccine with a high efficacy against the new variants introduced in our variant scenarios. It begins distributing vaccines three months after a new variant emerges to examine what happens if there is a globally coordinated effort to build targeted mRNA vaccines on an expedited timeline. There are two variations of this scenario, one in which targeted boosters are distributed at the same rate as the initial course of vaccination in each country or subnational unit, and one in which each country or subnational unit distributes the targeted boosters at the maximum rate observed in their respective WHO region.

### Uncertainty estimation

We propagated uncertainty from the drivers of the pandemic as well as the model parameters by representing model outputs with 100 iterations of the posterior distribution (draws) for all estimated quantities of interest, reported here as 95% uncertainty intervals (UIs).

### GATHER compliance

This study complies with the Guidelines for Accurate and Transparent Health Estimates Reporting (GATHER) recommendations (see appendix 1, section 3). All code used in the analysis can be found online https://github.com/ihmeuw/covid-model-seiir-pipeline.

## Results

### The Omicron waves (so far): November 15, 2021–December 12, 2022

Our model estimated that Omicron became the majority variant globally (by number of infections) on November 27, 2021 (Figure 6a), just 12 days after we consider the first Omicron wave to have begun. From November 15, 2021, to December 12, 2022, there were an estimated 8.60 billion (95% UI 6.37–11.7) SARS-CoV-2 infections globally, with Omicron (and its descendants such as BA.5) responsible for 8.47 billion ([6.20–11.6]; 98.5% [97.4–99.1]) (Figure 6d). Due to the higher severity of Delta and the lag between infection, severe symptoms, and death, larger proportions of all COVID-19 hospitalisations and deaths during this period were attributable to Delta compared to infections: 2.11 million ([1.50–3.02]; 16.1% [11.8–22.4]) of the estimated 13.1 million (10.6–16.5) COVID-19 hospitalisations and 0.755 million ([0.585–1.01]; 24.8% [20.5–31.4]) of the estimated 3.04 million (2.65–3.55) COVID-19 deaths in this period were attributable to Delta (Figure 6e, 6f).

Almost all 176 countries and territories in our analysis experienced at least one substantial Omicron wave over the historical study period (Figure 3a–3d), with 113 countries and territories seeing more than twice as many infections during the Omicron waves as had been previously estimated due to ancestral SARS-CoV-2 and all variants combined (Figure 3e). Underscoring Omicron’s lower severity compared to previous variants, many of these same locations saw only a fraction of deaths during this period compared to the pre-Omicron pandemic period (Figure 3f). Relative to previous waves, the WHO regions of the Americas and Europe continued to experience the largest per person death rates during the Omicron waves, while the WHO Western Pacific Region and South-East Asia Region avoided much of the severe outcome of the Omicron waves (Figure 2).

**Figure 2.**
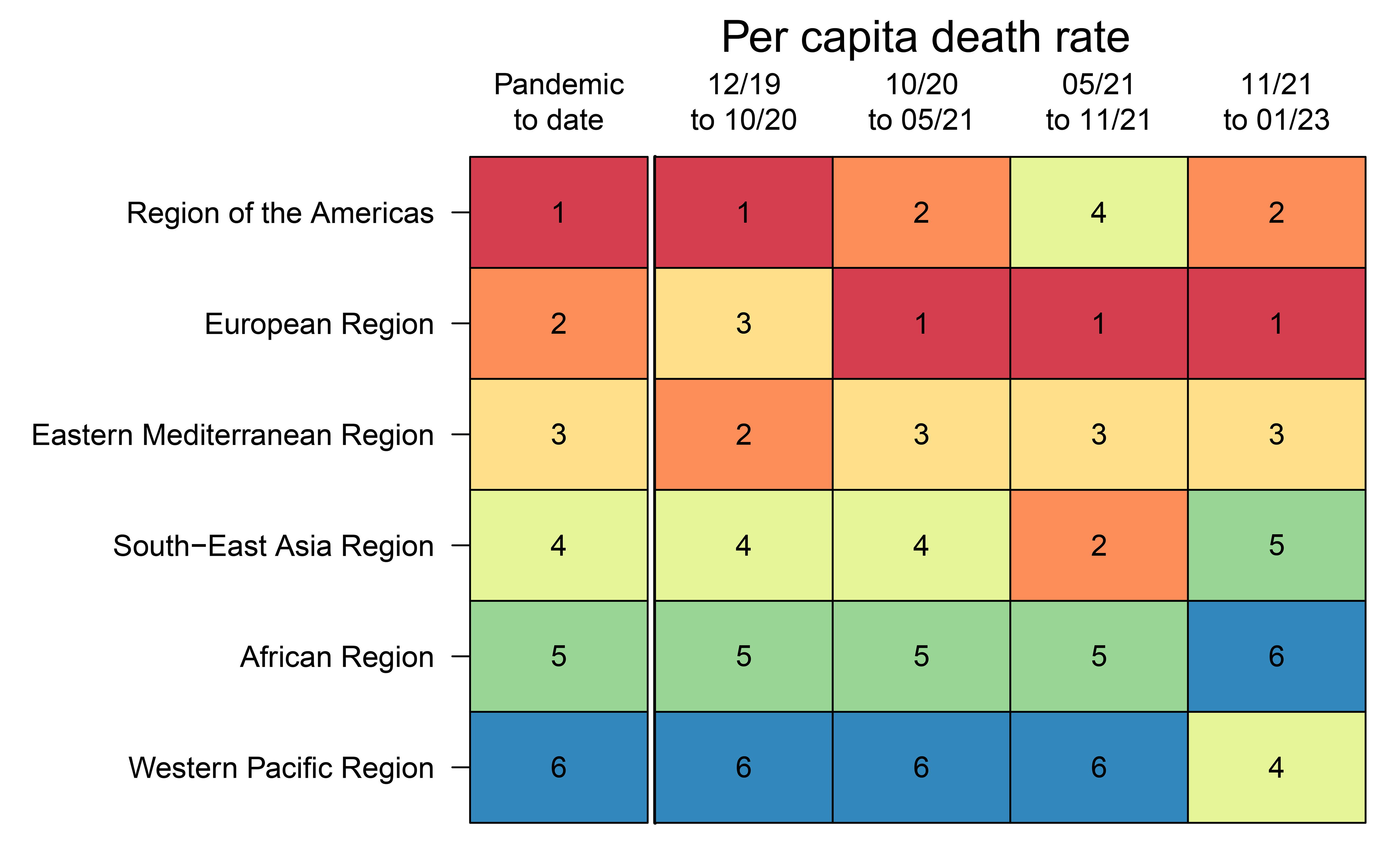
Per capita COVID-19 death rate ranking by WHO region. The far left “pandemic to date” column ranks the per capita death rate for each region for the entire pandemic up to December 12, 2022, by WHO region. This ranking determines relative ordering of WHO regions on the y-axis. The right four columns display relative rankings for four partitions of the pandemic, roughly corresponding to (a) pre-variant of concern phase, (b) Alpha, Beta, Gamma phase, (c) Delta phase, and (d) Omicron phase.

**Figure 3.**
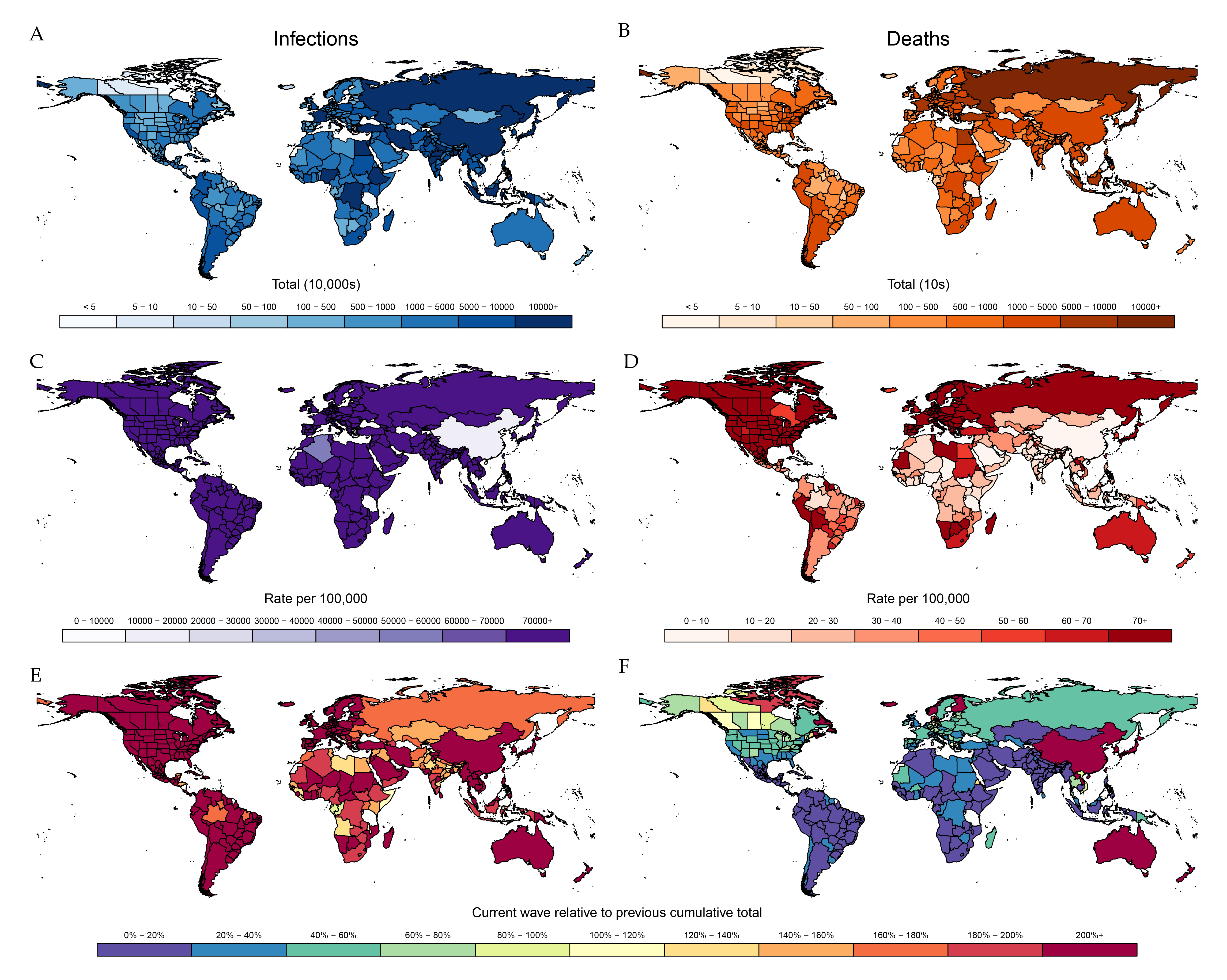
Cumulative total SARS-CoV-2 infections and COVID-19 deaths for Omicron wave from November 15, 2021, to December 12, 2022. Left column displays cumulative SARS-CoV-2 infections by location by count (A), rate (C), and relative size compared to entire pandemic up to November 15, 2021 (E). Corresponding values for COVID-19 deaths are shown in panels B, D, and F, respectively.

By December 12, 2022, an estimated 97.3% (95% UI 96.3–98.2) of the global population had experienced SARS-CoV-2 infection or vaccination or both, up from 83.2% (81.6–84.9) as of November 14, 2021 (Figure 4a, Movie S1). Notably, the proportion of the global population that had been both infected and vaccinated rose substantially, from 17.4% (16.5–18.3) on November 14, 2021, to 53.9% (49.9–58.1) on December 12, 2022. While all six WHO regions had exposure levels (from infection or vaccination) of at least 90% by December 12, 2022 (Region of the Americas: 99.5% [99.1–99.9; Figure 4b]; African Region: 97.2% [94.6–99.5; Figure 4e]; European Region: 99.5% [99.0–99.9, Figure 4c]; Eastern Mediterranean Region: 98.1% [97.9–99.8; Figure 4f]; Western Pacific Region: 94.0% [93.1–95.2; Figure 4d]; South-East Asia Region: 98.3% [97.0–99.5; Figure 4g]), the timing and composition of that exposure varied substantially. In the Western Pacific Region, the vast majority of all exposure was vaccine-derived because China’s zero-COVID policy has, to date, kept infections low (Figure 4d, Figure 5. Conversely, due to limited access to vaccines, the majority of all exposure (56.2% [53.6–58.5]) in the African Region was derived from infections alone. In the remaining regions, the majority of individuals were estimated to have been exposed from both vaccination and past infection (Region of the Americas: 81.1% [79.0–83.1]; European Region: 69.5% [66.3–72.3]; Eastern Mediterranean Region: 58.1% [55.4–60.4]; South-East Asia Region: 68.5% [63.6–73.1]; Figure 4b, 4c, 4f, 4g).

**Figure 4.**
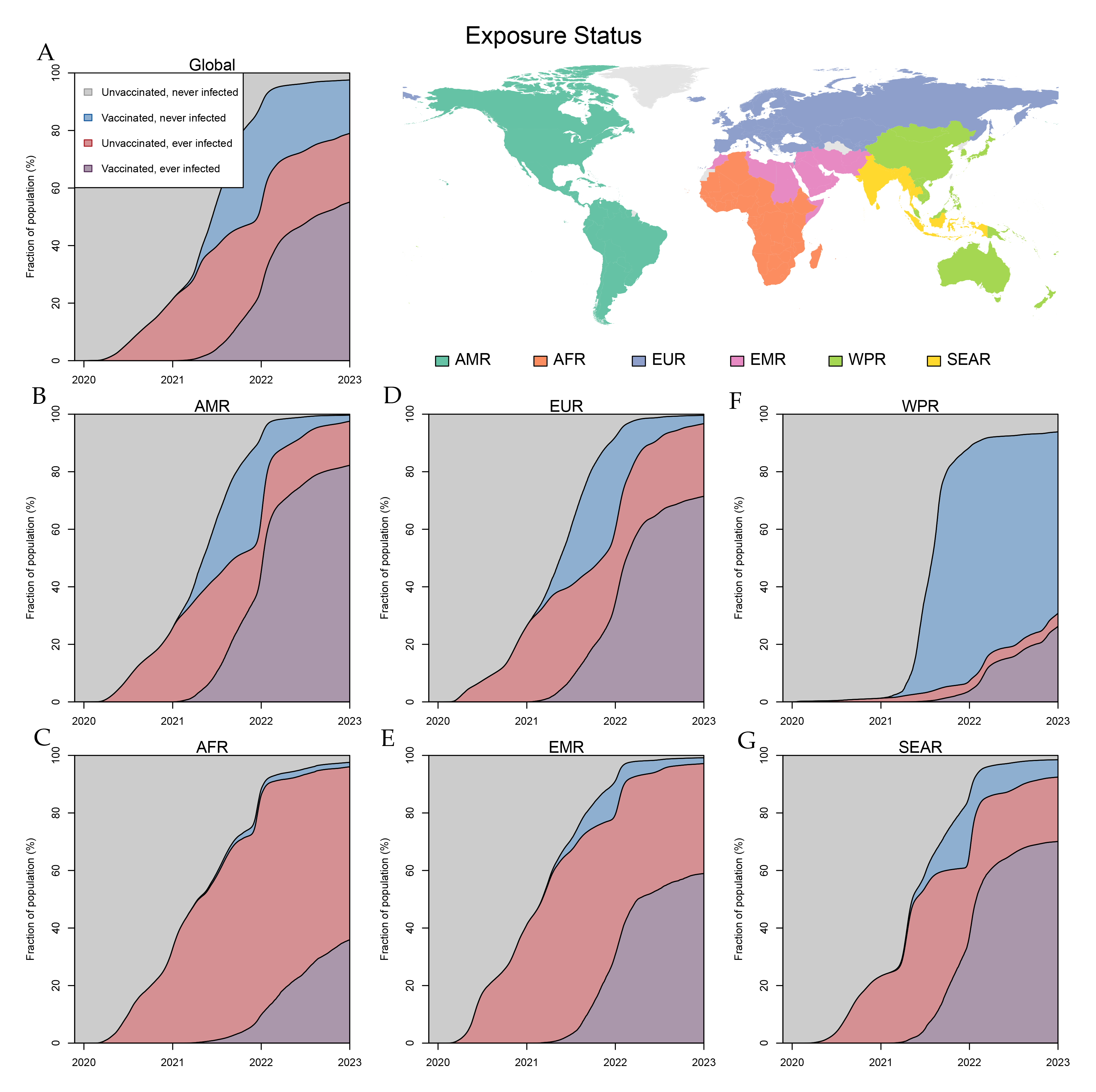
SARS-CoV-2 exposure status, December 2019 through December 12, 2022. Panel A: Global relative fractions of the population who have never been exposed to SARS-CoV-2 (grey), been vaccinated but never infected (blue), been previously infected but never vaccinated (red), or both vaccinated and previously infected (purple) are plotted through time from December 2019 through December 12, 2022. Panels B-G display corresponding information by WHO region. AMR = Region of the Americas. AFR = African Region. EMR = Eastern Mediterranean Region. EUR = European Region. SEAR = South-East Asia Region. WPR = Western Pacific Region.

**Figure 5.**
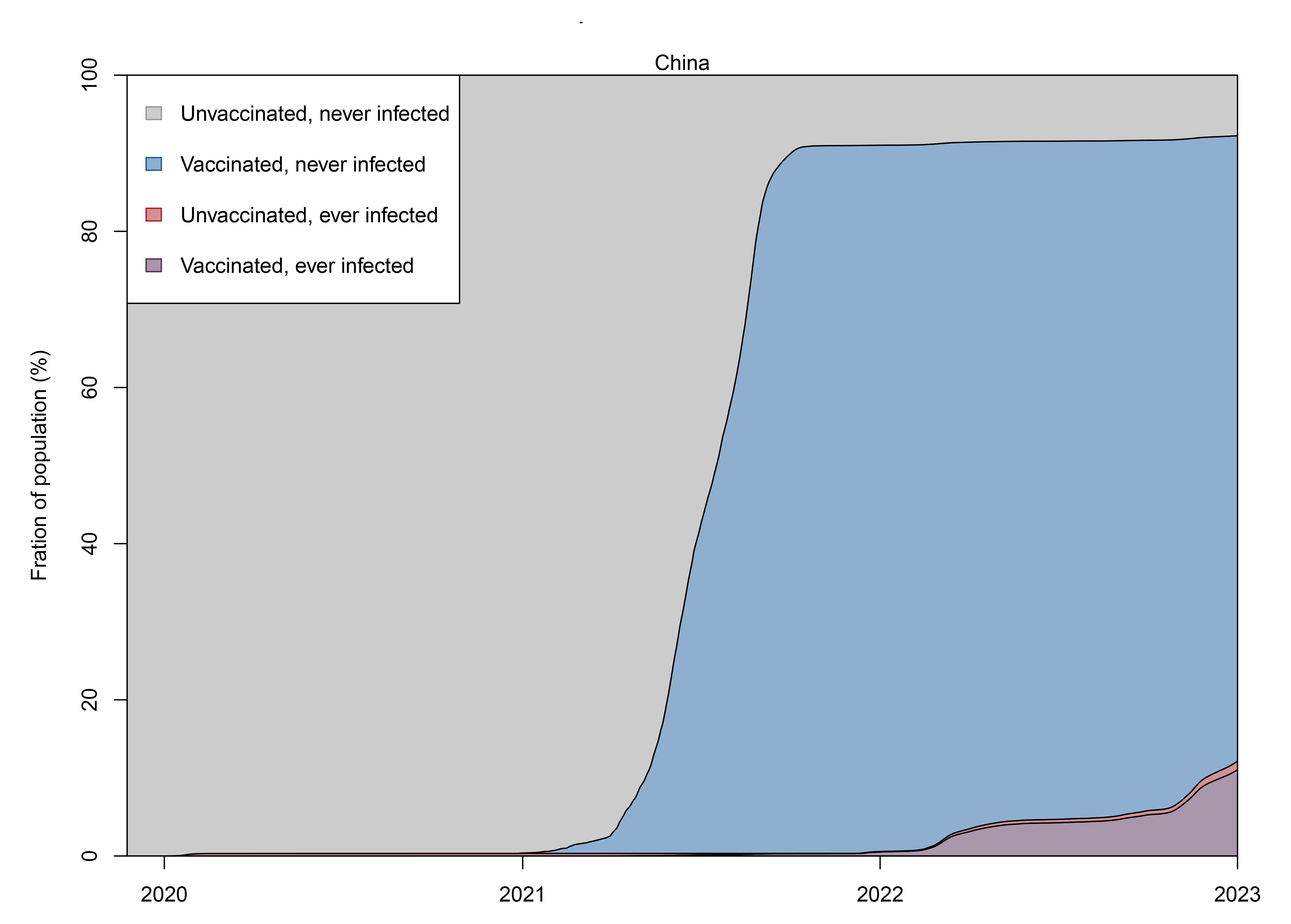
SARS-CoV-2 exposure status, December 2019 through December 12, 2022, in China. China relative fractions of the population who have never been exposed to SARS-CoV-2 (grey), been vaccinated but never infected (blue), been previously infected but never vaccinated (red), or both vaccinated and previously infected (purple) are plotted through time from December 2019 through December 12, 2022.

**Figure 6.**
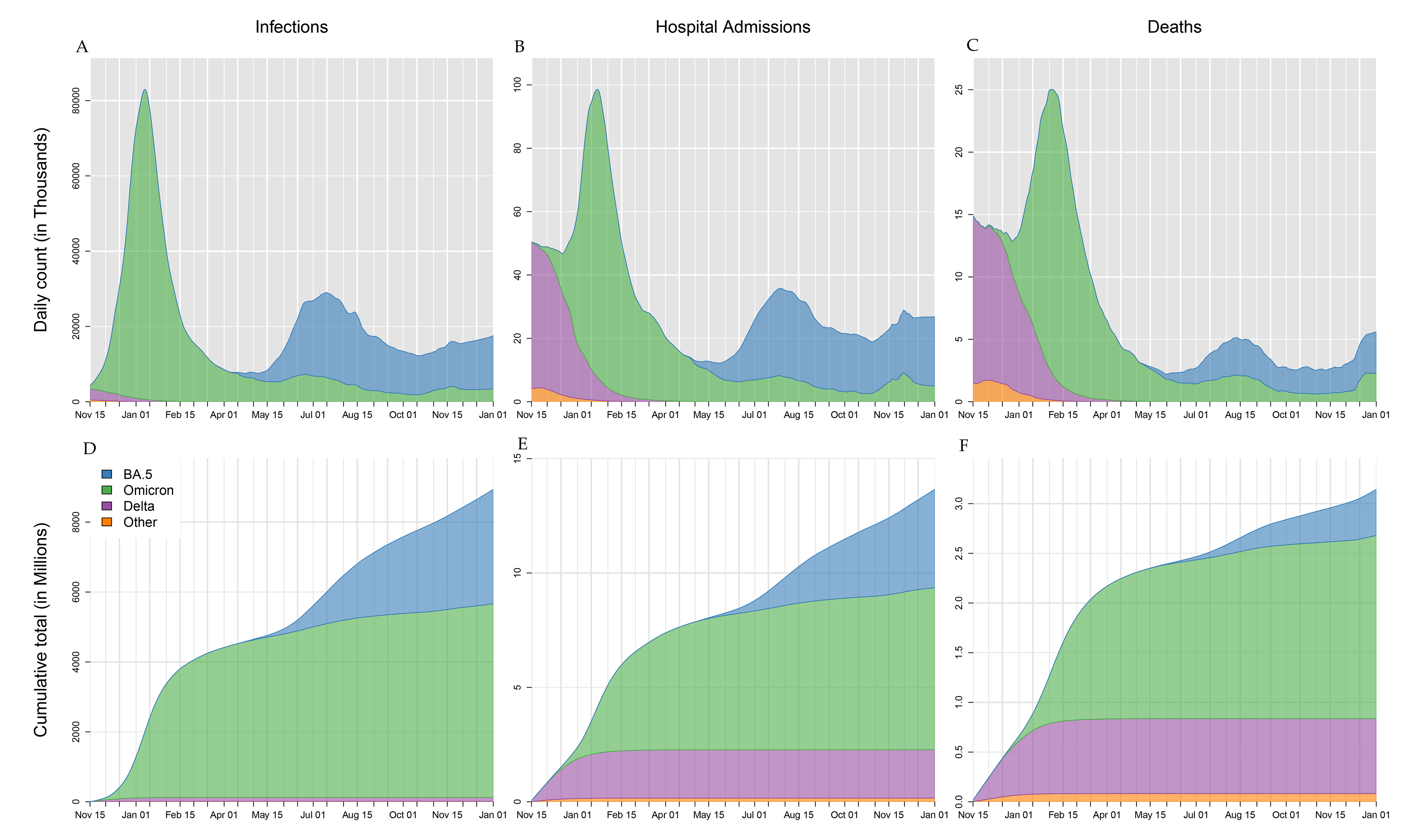
Global SARS-CoV-2 infections, COVID-19 hospital admissions, and COVID-19 deaths from November 15, 2021, through December 12, 2022. Infections are plotted by variant type: BA.5 subvariant of Omicron (blue), previous Omicron sub-variants (green), Delta (purple), other (orange). Daily counts are plotted in panels A, B, and C, while cumulative totals are plotted in panels D, E, and F.

### Five potential future variant scenarios under the reference intervention scenario

#### Variant scenario one: baseline

As a baseline, we evaluated a scenario where no new variant emerged—an improbable scenario if the past rates of variant emergence are maintained—and where mask usage, vaccine booster uptake, distribution of antivirals, and other drivers of transmission are set to plausible future levels (reference levels). Even in this scenario, due to incomplete or waning immunity and ongoing transmission of currently circulating variants, we estimated there will be 3.54 billion (95% UI 2.24–5.43) infections that result in 6.26 million (4.11–9.65) hospitalisations and 1.58 million (0.829–3.95) deaths between December 12, 2022, and June 30, 2023 (Figure 7d–f, red lines; Table 1). In this scenario, the daily infection rate will continue to increase until February and then slightly decline from March to June due to both seasonality in the Northern Hemisphere (Figure 7a, red line) and the expected continued removal of restrictions in China. As mentioned above, this scenario does not account for additional booster doses or an increase in mask usage due to perceived risk, which may ameliorate some of the predicted burden.

**Figure 7.**
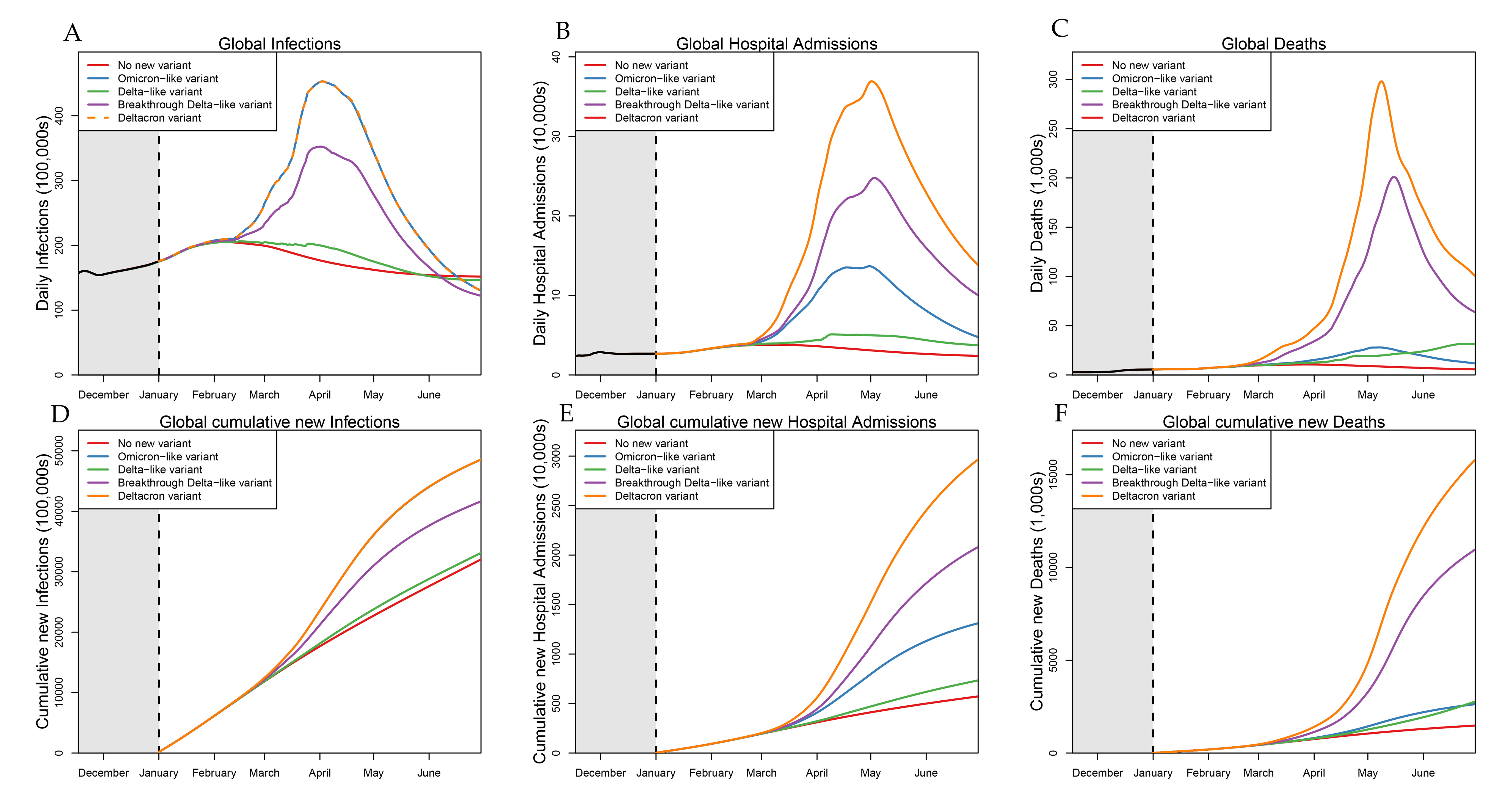
Global future variant scenarios for daily and cumulative COVID-19 infections (A, D), hospital admissions (B, E), and deaths (C, F), December 13, 2022–June 30, 2023. Panel A, global SARS-CoV-2 daily infections December 13, 2022 – June 30, 2023, for the scenario with no new variant (red line), an “Omicron-like” variant (blue line), a “Delta-like” variant (green line), a Breakthrough “Delta-like” variant (purple line) and DeltaCron (orange line). Panel B, daily COVID-19 hospital admissions; panel C, daily global COVID-19 deaths. The black line in panels A, B, and C depicts historical daily infections, hospital admissions, and deaths. Panels D, E, and F, display cumulative totals.

**Table 1.** Projected number of COVID-19 deaths for the reference intervention scenario under each variant scenario, by WHO region and for 177 countries and territories, December 13, 2022–June 30, 2023.

|  | No new variant | Breakthrough Delta-like variant | Delta-like variant | Omicron-like variant | DeltaCron variant |
| --- | --- | --- | --- | --- | --- |
| **Global** | **1,580,000**<br>**(791,000-4,500,000)** | **11,100,000**<br>**(2,390,000-46,000,000)** | **2,870,000**<br>**(1,010,000-7,840,000)** | **2,730,000**<br>**(1,310,000-6,730,000)** | **15,900,000**<br>**(3,950,000-73,800,000)** |
| **African Region** | **70,000**<br>**(16,100-180,000)** | **1,060,000**<br>**(133,000-3,880,000)** | **194,000**<br>**(39,200-836,000)** | **134,000**<br>**(34,100-461,000)** | **1,470,000**<br>**(211,000-6,000,000)** |
| Algeria | 56<br>(4-252) | 2,300<br>(13-25,200) | 371<br>(11-3,460) | 71<br>(4-334) | 5,990<br>(28-55,500) |
| Angola | 1,860<br>(131-8,950) | 13,700<br>(396-56,800) | 4,380<br>(246-21,100) | 1,970<br>(203-8,660) | 19,200<br>(518-61,500) |
| Benin | 118<br>(7-490) | 443<br>(39-1,680) | 156<br>(18-514) | 120<br>(11-480) | 1,320<br>(94-4,650) |
| Botswana | 1,180<br>(176-2,100) | 9,570<br>(849-18,600) | 2,100<br>(232-7,000) | 2,660<br>(879-4,760) | 12,400<br>(2,700-19,800) |
| Burkina Faso | 566<br>(48-1,420) | 4,810<br>(99-11,400) | 1,030<br>(67-3,130) | 1,040<br>(91-2,220) | 6,800<br>(145-14,500) |
| Cabo Verde | 17<br>(0-80) | 320<br>(0-1,270) | 95<br>(0-508) | 23<br>(0-77) | 420<br>(0-1,360) |
| Cameroon | 604<br>(161-1,380) | 5,420<br>(233-20,600) | 1,250<br>(184-5,410) | 1,300<br>(192-3,210) | 8,870<br>(346-28,800) |
| Central African Republic | 96<br>(1-580) | 775<br>(1-4,420) | 307<br>(1-2,380) | 169<br>(1-861) | 882<br>(1-4,460) |
| Chad | 411<br>(8-1,680) | 1,530<br>(13-6,390) | 517<br>(10-1,850) | 826<br>(10-3,080) | 3,340<br>(20-13,000) |
| Congo | 95<br>(1-314) | 619<br>(1-4,360) | 164<br>(1-630) | 154<br>(1-469) | 1,210<br>(1-5,220) |
| Côte d'Ivoire | 813<br>(1-2,020) | 9,700<br>(1-24,400) | 2,570<br>(1-9,980) | 2,060<br>(2-4,350) | 12,800<br>(2-29,500) |
| Democratic Republic of the Congo | 4,090<br>(403-13,500) | 24,900<br>(600-112,000) | 8,650<br>(421-34,500) | 10,500<br>(660-48,700) | 31,600<br>(1,100-167,000) |
| Equatorial Guinea | 86<br>(0-330) | 332<br>(0-1,330) | 116<br>(0-347) | 185<br>(0-481) | 1,100<br>(0-3,190) |
| Eritrea | 586<br>(81-2,820) | 13,900<br>(339-85,500) | 1,460<br>(199-6,750) | 1,110<br>(140-4,650) | 21,700<br>(482-119,000) |
| Eswatini | 9,120<br>(157-41,500) | 286,000<br>(703-1,180,000) | 33,300<br>(236-139,000) | 24,400<br>(331-118,000) | 341,000<br>(2,020-1,360,000) |
| Ethiopia | 4,560<br>(775-15,900) | 37,500<br>(5,660-106,000) | 7,150<br>(1,670-32,900) | 5,980<br>(1,850-17,300) | 61,300<br>(12,500-156,000) |
| Gabon | 18<br>(0-65) | 208<br>(0-1,430) | 39<br>(0-199) | 35<br>(0-125) | 453<br>(0-1,950) |
| Gambia | 234<br>(30-522) | 1,290<br>(82-4,140) | 348<br>(32-1,100) | 603<br>(85-1,570) | 1,860<br>(167-5,530) |
| Ghana | 135<br>(1-669) | 4,140<br>(1-26,800) | 621<br>(1-4,350) | 337<br>(1-1,890) | 6,300<br>(2-32,500) |
| Guinea | 1,080<br>(0-4,910) | 8,190<br>(0-41,800) | 2,330<br>(0-9,430) | 2,140<br>(0-10,500) | 10,700<br>(0-50,800) |
| Guinea-Bissau | 194<br>(4-630) | 1,520<br>(36-5,330) | 530<br>(10-2,190) | 310<br>(12-865) | 1,810<br>(67-5,620) |
| Kenya | 3,140<br>(76-13,500) | 36,800<br>(452-88,000) | 8,690<br>(139-36,700) | 2,790<br>(124-11,700) | 49,100<br>(1,100-107,000) |
| Lesotho | 659<br>(23-2,600) | 6,730<br>(158-27,900) | 1,450<br>(38-6,520) | 1,700<br>(53-6,710) | 8,170<br>(254-30,600) |
| Liberia | 266<br>(34-1,700) | 923<br>(122-4,320) | 319<br>(63-2,110) | 379<br>(77-2,660) | 1,580<br>(169-5,190) |
| Madagascar | 3,470<br>(386-14,500) | 19,600<br>(5,010-50,700) | 7,170<br>(1,590-21,000) | 4,050<br>(941-13,400) | 24,600<br>(7,260-55,500) |
| Malawi | 1,930<br>(10-9,680) | 10,500<br>(13-48,000) | 4,930<br>(11-30,000) | 1,940<br>(10-9,590) | 13,000<br>(15-62,600) |
| Mali | 264<br>(50-783) | 2,630<br>(71-21,400) | 615<br>(65-4,220) | 313<br>(53-883) | 3,490<br>(86-23,700) |
| Mauritania | 1,940<br>(0-12,900) | 24,000<br>(0-99,200) | 2,860<br>(0-14,600) | 12,600<br>(0-58,700) | 94,500<br>(0-190,000) |
| Mozambique | 12,600<br>(453-61,600) | 293,000<br>(9,240-1,610,000) | 53,500<br>(1,830-309,000) | 18,900<br>(819-95,800) | 355,000<br>(16,600-1,650,000) |
| Namibia | 442<br>(106-1,120) | 7,390<br>(218-26,900) | 1,540<br>(127-6,780) | 950<br>(162-2,740) | 10,600<br>(649-36,600) |
| Niger | 230<br>(48-504) | 1,380<br>(87-7,150) | 336<br>(61-768) | 331<br>(52-743) | 2,310<br>(116-9,210) |
| Nigeria | 298<br>(29-939) | 12,700<br>(88-81,100) | 1,420<br>(43-11,900) | 574<br>(33-1,880) | 26,800<br>(167-102,000) |
| Rwanda | 484<br>(44-1,430) | 3,600<br>(72-14,100) | 758<br>(49-2,430) | 875<br>(54-1,970) | 6,400<br>(148-18,900) |
| Senegal | 155<br>(22-367) | 7,610<br>(82-32,500) | 2,030<br>(39-10,200) | 266<br>(46-679) | 9,740<br>(106-35,100) |
| Sierra Leone | 122<br>(5-1,150) | 725<br>(14-5,250) | 145<br>(7-1,190) | 421<br>(12-2,020) | 1,660<br>(21-8,470) |
| South Africa | 5,100<br>(1,000-18,500) | 39,500<br>(4,450-106,000) | 9,950<br>(1,590-45,200) | 5,540<br>(1,610-16,400) | 58,300<br>(10,100-128,000) |
| South Sudan | 49<br>(0-186) | 1,270<br>(0-5,090) | 291<br>(0-1,510) | 133<br>(0-400) | 1,810<br>(0-6,040) |
| Togo | 250<br>(49-823) | 1,540<br>(76-6,130) | 448<br>(59-2,070) | 572<br>(80-1,870) | 2,450<br>(160-9,060) |
| Uganda | 4,150<br>(37-35,800) | 28,900<br>(172-278,000) | 7,770<br>(82-45,500) | 4,430<br>(64-36,800) | 52,900<br>(504-452,000) |
| Zambia | 1,480<br>(0-12,400) | 40,400<br>(0-295,000) | 4,540<br>(0-45,100) | 2,000<br>(0-18,900) | 55,000<br>(1-385,000) |
| Zimbabwe | 7,080<br>(579-62,400) | 96,900<br>(1,180-783,000) | 18,000<br>(676-83,300) | 18,900<br>(1,210-163,000) | 145,000<br>(2,750-1,440,000) |
| Eastern Mediterranean Region | 43,000<br>(12,700-163,000) | 913,000<br>(60,500-3,390,000) | 192,000<br>(24,500-1,170,000) | 73,100<br>(23,400-312,000) | 1,160,000<br>(114,000-3,790,000) |
| Afghanistan | 5,460<br>(818-22,400) | 98,100<br>(5,340-486,000) | 22,200<br>(1,540-185,000) | 9,940<br>(1,630-37,100) | 118,000<br>(10,400-528,000) |
| Bahrain | 79<br>(6-173) | 666<br>(10-2,570) | 134<br>(6-475) | 244<br>(12-582) | 1,010<br>(38-2,850) |
| Egypt | 1,250<br>(249-3,970) | 17,900<br>(689-78,600) | 2,050<br>(471-6,930) | 1,580<br>(404-3,760) | 30,400<br>(1,090-92,700) |
| Iran (Islamic Republic of) | 4,860<br>(1,940-8,950) | 48,500<br>(2,820-173,000) | 6,210<br>(2,190-15,700) | 10,800<br>(2,290-23,800) | 81,800<br>(3,790-199,000) |
| Iraq | 1,340<br>(79-5,040) | 17,300<br>(840-85,600) | 5,480<br>(305-35,100) | 1,460<br>(153-4,720) | 30,000<br>(1,370-102,000) |
| Jordan | 479<br>(127-1,290) | 12,600<br>(1,290-41,300) | 3,140<br>(310-15,300) | 840<br>(333-1,960) | 16,300<br>(3,390-47,100) |
| Kuwait | 32<br>(4-126) | 1,660<br>(9-17,300) | 93<br>(5-532) | 83<br>(5-445) | 3,530<br>(13-22,900) |
| Lebanon | 876<br>(27-2,000) | 9,920<br>(117-35,100) | 1,810<br>(35-8,330) | 2,500<br>(136-6,800) | 14,300<br>(159-39,500) |
| Libya | 210<br>(10-942) | 6,120<br>(20-46,900) | 2,610<br>(18-18,300) | 232<br>(16-985) | 6,880<br>(31-48,400) |
| Morocco | 3,660<br>(313-14,600) | 32,500<br>(1,040-161,000) | 6,630<br>(493-22,800) | 3,990<br>(427-13,800) | 50,900<br>(2,440-181,000) |
| Oman | 38<br>(0-183) | 301<br>(0-2,760) | 49<br>(0-245) | 105<br>(0-491) | 960<br>(1-5,150) |
| Pakistan | 8,860<br>(2,470-18,500) | 173,000<br>(11,400-917,000) | 25,600<br>(3,370-97,400) | 15,800<br>(5,310-55,500) | 236,000<br>(27,500-1,030,000) |
| Palestine | 263<br>(5-728) | 3,850<br>(7-12,800) | 1,140<br>(6-6,060) | 482<br>(6-1,170) | 5,160<br>(10-12,900) |
| Qatar | 42<br>(8-99) | 657<br>(83-1,830) | 87<br>(10-366) | 116<br>(24-279) | 859<br>(118-2,350) |
| Saudi Arabia | 915<br>(336-2,370) | 20,300<br>(444-88,200) | 3,200<br>(367-16,400) | 2,590<br>(373-7,830) | 32,900<br>(789-109,000) |
| Somalia | 375<br>(0-1,850) | 19,700<br>(1-115,000) | 2,790<br>(0-18,600) | 613<br>(0-3,240) | 27,900<br>(5-176,000) |
| Sudan | 825<br>(52-5,880) | 37,700<br>(54-354,000) | 10,000<br>(52-81,300) | 1,870<br>(52-18,200) | 47,000<br>(56-500,000) |
| Syrian Arab Republic | 74<br>(4-263) | 2,340<br>(37-28,900) | 412<br>(19-2,640) | 111<br>(9-510) | 4,320<br>(67-36,100) |
| Tunisia | 255<br>(2-1,490) | 5,790<br>(13-40,600) | 1,020<br>(3-10,300) | 364<br>(5-1,470) | 8,110<br>(21-45,700) |
| United Arab Emirates | 102<br>(3-284) | 3,270<br>(39-18,400) | 474<br>(12-3,130) | 253<br>(18-918) | 5,780<br>(118-22,200) |
| Yemen | 13,000<br>(13-136,000) | 401,000<br>(50-2,150,000) | 96,800<br>(20-1,040,000) | 19,200<br>(14-258,000) | 437,000<br>(88-2,320,000) |
| European Region | 215,000<br>(147,000-302,000) | 1,600,000<br>(414,000-3,380,000) | 365,000<br>(186,000-879,000) | 571,000<br>(301,000-968,000) | 2,340,000<br>(734,000-4,190,000) |
| Albania | 401<br>(4-727) | 3,300<br>(23-10,400) | 688<br>(4-2,750) | 1,030<br>(61-2,210) | 5,040<br>(251-12,000) |
| Andorra | 24<br>(12-45) | 273<br>(29-527) | 57<br>(14-210) | 65<br>(25-119) | 327<br>(68-565) |
| Armenia | 611<br>(224-1,820) | 6,740<br>(485-23,800) | 1,930<br>(325-7,420) | 1,140<br>(352-3,600) | 8,750<br>(789-33,000) |
| Austria | 1,630<br>(1,090-2,600) | 9,100<br>(1,440-25,600) | 1,910<br>(1,230-3,530) | 4,560<br>(1,360-9,530) | 15,300<br>(1,790-34,700) |
| Azerbaijan | 2,200<br>(373-6,710) | 13,700<br>(400-84,000) | 3,750<br>(384-13,700) | 4,320<br>(378-20,500) | 19,300<br>(427-118,000) |
| Belarus | 4,690<br>(1,580-12,000) | 53,600<br>(17,200-87,100) | 12,500<br>(2,390-32,100) | 7,730<br>(3,420-13,800) | 58,800<br>(28,000-97,300) |
| Belgium | 1,750<br>(651-5,030) | 11,900<br>(1,390-30,200) | 2,030<br>(726-5,200) | 4,950<br>(1,300-12,200) | 19,700<br>(3,250-40,000) |
| Bosnia and Herzegovina | 649<br>(118-1,850) | 6,010<br>(478-16,500) | 1,840<br>(175-5,960) | 1,200<br>(295-2,630) | 7,780<br>(504-18,000) |
| Bulgaria | 2,180<br>(691-4,830) | 15,900<br>(838-67,900) | 3,400<br>(743-9,460) | 4,670<br>(753-13,500) | 27,400<br>(1,030-90,400) |
| Croatia | 1,730<br>(903-2,880) | 9,590<br>(1,330-22,200) | 2,980<br>(1,060-8,060) | 3,780<br>(1,200-6,580) | 12,700<br>(2,230-26,700) |
| Cyprus | 134<br>(38-236) | 963<br>(61-4,300) | 227<br>(53-732) | 328<br>(57-805) | 1,960<br>(67-5,890) |
| Czechia | 1,390<br>(235-4,810) | 11,000<br>(1,270-28,900) | 3,400<br>(404-14,300) | 1,420<br>(415-4,400) | 14,700<br>(2,490-30,500) |
| Denmark | 5,190<br>(2,030-11,100) | 20,700<br>(3,000-60,700) | 5,590<br>(2,100-12,200) | 12,600<br>(3,170-27,600) | 33,700<br>(4,400-85,000) |
| Estonia | 600<br>(374-861) | 2,500<br>(501-8,470) | 666<br>(379-1,300) | 1,420<br>(488-2,770) | 4,860<br>(680-11,200) |
| Finland | 4,130<br>(1,200-7,570) | 17,100<br>(1,500-40,400) | 5,500<br>(1,260-17,200) | 10,500<br>(1,520-21,300) | 20,800<br>(1,560-42,800) |
| France | 19,600<br>(13,900-30,100) | 95,700<br>(19,300-203,000) | 21,300<br>(14,500-31,500) | 56,900<br>(20,000-104,000) | 145,000<br>(27,100-257,000) |
| Georgia | 353<br>(22-809) | 6,100<br>(42-19,500) | 2,660<br>(27-12,200) | 472<br>(33-964) | 7,320<br>(80-19,700) |
| Germany | 31,700<br>(20,000-44,700) | 112,000<br>(27,400-357,000) | 34,200<br>(20,800-48,300) | 98,000<br>(32,300-209,000) | 200,000<br>(40,300-480,000) |
| Greece | 6,160<br>(3,570-10,300) | 23,600<br>(6,270-59,600) | 6,710<br>(3,790-11,400) | 18,400<br>(6,630-37,500) | 37,500<br>(8,520-77,100) |
| Hungary | 1,980<br>(927-3,680) | 21,400<br>(1,850-55,800) | 6,760<br>(1,090-23,800) | 3,920<br>(1,510-7,440) | 28,500<br>(2,220-60,100) |
| Iceland | 42<br>(0-155) | 334<br>(0-1,640) | 65<br>(0-275) | 96<br>(0-315) | 630<br>(0-2,230) |
| Ireland | 1,620<br>(224-4,870) | 4,530<br>(442-17,300) | 1,690<br>(232-5,090) | 4,480<br>(557-14,000) | 8,240<br>(726-26,700) |
| Israel | 1,330<br>(880-2,100) | 7,440<br>(1,610-16,900) | 1,930<br>(1,130-3,640) | 3,890<br>(1,610-7,530) | 10,900<br>(2,630-21,700) |
| Italy | 32,700<br>(22,200-54,800) | 222,000<br>(45,600-466,000) | 39,700<br>(24,700-80,900) | 97,900<br>(41,300-191,000) | 325,000<br>(75,600-560,000) |
| Kazakhstan | 708<br>(0-1,850) | 9,930<br>(1-51,800) | 1,660<br>(0-8,430) | 1,610<br>(0-5,290) | 16,900<br>(5-58,700) |
| Kyrgyzstan | 532<br>(4-7,020) | 11,500<br>(5-123,000) | 1,790<br>(4-24,900) | 920<br>(4-9,830) | 17,700<br>(8-163,000) |
| Latvia | 694<br>(82-2,660) | 6,120<br>(335-25,800) | 2,520<br>(215-12,600) | 724<br>(130-2,580) | 8,200<br>(429-27,600) |
| Lithuania | 632<br>(53-1,810) | 6,370<br>(385-43,100) | 2,160<br>(129-21,200) | 639<br>(87-1,800) | 8,650<br>(607-42,600) |
| Luxembourg | 115<br>(32-205) | 771<br>(93-2,570) | 134<br>(39-246) | 332<br>(92-654) | 1,290<br>(206-3,130) |
| Malta | 57<br>(3-166) | 693<br>(4-2,260) | 110<br>(3-433) | 95<br>(4-224) | 1,010<br>(7-2,390) |
| Monaco | 9<br>(2-22) | 144<br>(7-465) | 23<br>(3-105) | 25<br>(4-60) | 208<br>(27-533) |
| Montenegro | 130<br>(20-369) | 770<br>(28-2,790) | 194<br>(22-663) | 254<br>(25-613) | 1,130<br>(43-3,100) |
| Netherlands | 1,160<br>(131-3,270) | 28,700<br>(216-160,000) | 4,910<br>(156-31,600) | 2,920<br>(168-8,550) | 51,000<br>(614-188,000) |
| North Macedonia | 812<br>(224-1,420) | 6,590<br>(953-15,200) | 1,460<br>(317-4,780) | 1,860<br>(603-3,660) | 8,690<br>(1,800-17,300) |
| Norway | 2,370<br>(645-6,700) | 4,510<br>(908-12,800) | 2,400<br>(658-6,720) | 5,900<br>(1,100-17,200) | 7,040<br>(1,220-22,900) |
| Poland | 5,160<br>(1,820-12,000) | 78,400<br>(3,300-211,000) | 16,400<br>(1,960-71,700) | 9,880<br>(2,190-19,200) | 106,000<br>(6,280-245,000) |
| Portugal | 5,140<br>(2,230-8,910) | 18,500<br>(4,660-40,200) | 5,430<br>(2,380-9,100) | 16,600<br>(5,640-29,300) | 29,500<br>(7,500-52,600) |
| Republic of Moldova | 333<br>(3-2,010) | 6,390<br>(14-67,500) | 2,410<br>(3-34,500) | 470<br>(6-2,500) | 10,400<br>(84-80,700) |
| Romania | 2,650<br>(920-7,090) | 30,600<br>(2,000-128,000) | 9,690<br>(1,350-55,600) | 3,820<br>(1,300-8,210) | 41,300<br>(2,970-136,000) |
| Russian Federation | 23,600<br>(9,450-51,500) | 313,000<br>(19,300-743,000) | 67,500<br>(11,600-285,000) | 55,300<br>(15,000-133,000) | 373,000<br>(36,600-780,000) |
| San Marino | 9<br>(4-18) | 96<br>(14-251) | 17<br>(6-66) | 22<br>(8-43) | 140<br>(34-291) |
| Serbia | 3,000<br>(1,450-5,290) | 20,000<br>(3,290-50,600) | 6,270<br>(1,710-19,900) | 7,220<br>(2,200-14,100) | 26,400<br>(5,220-57,300) |
| Slovakia | 1,800<br>(317-5,870) | 18,800<br>(465-89,300) | 4,950<br>(358-15,700) | 4,630<br>(417-19,700) | 25,000<br>(846-121,000) |
| Slovenia | 443<br>(223-664) | 8,750<br>(431-22,800) | 1,880<br>(284-8,200) | 1,210<br>(366-2,120) | 12,000<br>(1,080-24,900) |
| Spain | 17,300<br>(9,100-32,100) | 98,400<br>(21,700-266,000) | 20,600<br>(10,500-38,400) | 52,200<br>(20,000-90,500) | 156,000<br>(39,400-314,000) |
| Sweden | 2,970<br>(23-7,530) | 7,890<br>(27-31,800) | 3,100<br>(23-7,650) | 7,250<br>(39-20,100) | 15,100<br>(61-54,600) |
| Switzerland | 583<br>(137-1,220) | 12,700<br>(350-41,400) | 1,230<br>(166-4,760) | 1,510<br>(237-3,430) | 22,500<br>(1,470-55,700) |
| Turkey | 5,240<br>(1,040-12,400) | 97,100<br>(6,230-384,000) | 21,900<br>(1,920-124,000) | 7,700<br>(2,160-15,400) | 140,000<br>(11,700-423,000) |
| Ukraine | 2,010<br>(261-9,720) | 23,800<br>(651-208,000) | 4,880<br>(397-27,000) | 2,480<br>(330-12,100) | 45,100<br>(839-252,000) |
| United Kingdom | 13,100<br>(4,810-27,100) | 97,200<br>(9,570-328,000) | 18,600<br>(6,340-48,400) | 40,000<br>(9,190-91,100) | 174,000<br>(21,200-466,000) |
| Uzbekistan | 1,340<br>(82-4,530) | 13,600<br>(1,160-36,200) | 1,740<br>(182-4,980) | 1,440<br>(194-3,990) | 23,600<br>(4,080-47,300) |
| Region of the Americas | 209,000<br>(139,000-310,000) | 2,010,000<br>(707,000-3,350,000) | 358,000<br>(172,000-812,000) | 559,000<br>(333,000-847,000) | 2,720,000<br>(1,360,000-4,080,000) |
| Antigua and Barbuda | 15<br>(6-27) | 136<br>(12-360) | 30<br>(7-105) | 39<br>(10-76) | 207<br>(18-421) |
| Argentina | 3,360<br>(1,130-9,300) | 94,100<br>(5,810-212,000) | 13,700<br>(1,620-71,100) | 5,730<br>(2,120-11,700) | 131,000<br>(13,100-272,000) |
| Bahamas | 41<br>(10-94) | 317<br>(18-1,020) | 66<br>(10-251) | 95<br>(16-224) | 537<br>(37-1,380) |
| Barbados | 742<br>(285-1,540) | 1,490<br>(476-2,990) | 799<br>(308-1,760) | 1,880<br>(693-3,480) | 1,800<br>(652-3,220) |
| Belize | 36<br>(2-80) | 380<br>(4-1,020) | 92<br>(2-432) | 94<br>(6-194) | 538<br>(11-1,170) |
| Bolivia (Plurinational State of) | 3,210<br>(603-7,180) | 26,700<br>(786-62,700) | 6,400<br>(614-22,500) | 6,550<br>(903-14,400) | 32,800<br>(2,330-73,000) |
| Brazil | 33,700<br>(20,700-52,900) | 373,000<br>(152,000-604,000) | 66,000<br>(29,100-168,000) | 90,200<br>(55,900-147,000) | 451,000<br>(255,000-672,000) |
| Canada | 9,550<br>(5,730-16,200) | 55,400<br>(10,100-138,000) | 13,700<br>(6,020-33,000) | 23,400<br>(8,900-42,500) | 90,600<br>(18,300-180,000) |
| Chile | 2,950<br>(1,920-4,190) | 36,200<br>(3,690-92,000) | 5,880<br>(2,580-21,300) | 7,280<br>(2,950-13,100) | 60,600<br>(7,260-117,000) |
| Colombia | 1,770<br>(5-3,510) | 18,300<br>(38-104,000) | 2,080<br>(5-3,880) | 3,290<br>(26-8,210) | 38,400<br>(548-127,000) |
| Costa Rica | 522<br>(270-1,110) | 3,430<br>(435-10,100) | 587<br>(279-1,390) | 1,770<br>(495-3,540) | 5,710<br>(616-13,800) |
| Cuba | 306<br>(39-717) | 2,630<br>(153-10,900) | 538<br>(59-1,430) | 448<br>(50-1,160) | 8,730<br>(562-40,500) |
| Dominica | 22<br>(0-65) | 55<br>(0-252) | 25<br>(0-84) | 70<br>(0-265) | 88<br>(0-316) |
| Dominican Republic | 162<br>(7-399) | 9,190<br>(99-26,200) | 1,030<br>(9-5,730) | 300<br>(57-576) | 13,800<br>(817-27,800) |
| Ecuador | 1,670<br>(974-2,560) | 44,400<br>(6,330-80,400) | 5,390<br>(1,260-21,100) | 4,310<br>(2,220-7,170) | 55,500<br>(24,900-88,400) |
| El Salvador | 835<br>(384-1,590) | 10,600<br>(753-33,000) | 2,090<br>(474-9,880) | 2,170<br>(564-4,630) | 15,800<br>(1,320-37,000) |
| Grenada | 23<br>(10-54) | 88<br>(11-316) | 29<br>(12-68) | 74<br>(16-200) | 143<br>(18-368) |
| Guatemala | 1,810<br>(53-3,720) | 15,500<br>(326-40,500) | 2,970<br>(69-10,500) | 4,950<br>(384-10,300) | 21,900<br>(934-45,100) |
| Guyana | 335<br>(66-3,300) | 1,860<br>(143-5,730) | 539<br>(74-3,440) | 609<br>(133-4,010) | 2,650<br>(259-10,000) |
| Haiti | 177<br>(4-970) | 1,700<br>(9-12,700) | 388<br>(7-1,550) | 273<br>(7-1,320) | 3,070<br>(19-19,200) |
| Honduras | 1,860<br>(382-6,340) | 23,200<br>(920-71,000) | 6,010<br>(406-29,500) | 3,960<br>(644-13,600) | 27,300<br>(1,620-75,300) |
| Jamaica | 401<br>(4-2,060) | 983<br>(5-5,760) | 463<br>(4-2,340) | 1,040<br>(6-5,840) | 1,990<br>(7-10,500) |
| Mexico | 18,400<br>(9,990-32,200) | 269,000<br>(60,200-536,000) | 44,000<br>(12,500-135,000) | 43,700<br>(23,900-72,000) | 360,000<br>(131,000-593,000) |
| Panama | 712<br>(488-1,070) | 7,900<br>(1,510-14,700) | 1,080<br>(571-2,850) | 2,130<br>(1,250-3,310) | 10,500<br>(4,310-16,800) |
| Paraguay | 1,430<br>(379-4,260) | 7,200<br>(3,530-17,800) | 3,730<br>(1,090-7,940) | 2,100<br>(729-5,940) | 7,750<br>(3,670-19,100) |
| Peru | 12,100<br>(7,130-25,700) | 188,000<br>(51,800-327,000) | 33,100<br>(10,300-96,500) | 34,500<br>(18,100-60,300) | 221,000<br>(113,000-364,000) |
| Saint Kitts and Nevis | 7<br>(2-19) | 65<br>(3-222) | 10<br>(2-26) | 13<br>(2-38) | 117<br>(5-328) |
| Saint Lucia | 21<br>(4-60) | 71<br>(4-452) | 26<br>(4-90) | 84<br>(5-394) | 174<br>(5-973) |
| Saint Vincent and the Grenadines | 40<br>(12-121) | 216<br>(23-820) | 57<br>(15-197) | 99<br>(23-324) | 336<br>(36-1,410) |
| Suriname | 72<br>(5-260) | 875<br>(5-3,060) | 223<br>(5-1,250) | 119<br>(5-375) | 1,210<br>(7-3,340) |
| Trinidad and Tobago | 701<br>(82-1,790) | 2,740<br>(109-9,470) | 883<br>(97-2,120) | 2,560<br>(129-8,300) | 4,670<br>(116-15,100) |
| United States of America | 111,000<br>(66,600-201,000) | 782,000<br>(206,000-1,420,000) | 142,000<br>(80,900-299,000) | 314,000<br>(156,000-567,000) | 1,090,000<br>(375,000-1,850,000) |
| Uruguay | 480<br>(202-966) | 8,160<br>(393-20,600) | 1,780<br>(237-7,500) | 844<br>(339-1,330) | 11,700<br>(750-22,400) |
| Venezuela (Bolivarian Republic of) | 258<br>(9-1,240) | 25,900<br>(77-145,000) | 2,480<br>(22-14,800) | 549<br>(17-2,440) | 52,800<br>(236-272,000) |
| **South-East Asia Region** | **71,500**<br>**(18,300-492,000)** | **3,410,000**<br>**(58,800-22,100,000)** | **646,000**<br>**(25,500-1,170,000)** | **179,000**<br>**(27,300-1,470,000)** | **4,830,000**<br>**(110,000-48,600,000)** |
| Bangladesh | 1,630<br>(110-5,430) | 28,200<br>(149-217,000) | 3,020<br>(117-13,200) | 4,390<br>(134-17,400) | 57,500<br>(213-268,000) |
| Bhutan | 39<br>(1-171) | 437<br>(2-1,930) | 149<br>(1-851) | 58<br>(1-211) | 526<br>(2-1,960) |
| India | 50,900<br>(5,270-470,000) | 3,130,000<br>(14,600-22,000,000) | 578,000<br>(7,470-1,120,000) | 141,000<br>(6,760-1,420,000) | 4,370,000<br>(28,900-48,000,000) |
| Indonesia | 12,200<br>(2,930-26,300) | 178,000<br>(5,800-752,000) | 54,200<br>(3,090-295,000) | 21,300<br>(4,860-54,300) | 247,000<br>(6,870-819,000) |
| Maldives | 10<br>(0-36) | 138<br>(1-954) | 17<br>(0-63) | 36<br>(1-174) | 284<br>(1-1,760) |
| Myanmar | 133<br>(5-623) | 8,510<br>(13-90,700) | 513<br>(7-2,840) | 285<br>(5-1,480) | 16,300<br>(45-117,000) |
| Nepal | 896<br>(168-2,480) | 14,200<br>(325-63,700) | 1,440<br>(204-5,740) | 1,760<br>(205-7,910) | 25,400<br>(599-91,000) |
| Sri Lanka | 292<br>(50-644) | 11,400<br>(130-58,800) | 995<br>(83-4,470) | 795<br>(76-2,610) | 19,200<br>(373-65,200) |
| Thailand | 5,390<br>(1,220-9,570) | 37,700<br>(2,280-166,000) | 8,310<br>(1,580-20,800) | 9,010<br>(1,680-19,600) | 93,200<br>(2,980-293,000) |
| Timor-Leste | 51<br>(10-201) | 140<br>(20-626) | 64<br>(13-237) | 101<br>(18-371) | 314<br>(38-1,370) |
| **Western Pacific Region** | **963,000**<br>**(312,000-3,840,000)** | **2,040,000**<br>**(463,000-8,130,000)** | **1,100,000**<br>**(371,000-4,100,000)** | **1,200,000**<br>**(405,000-4,590,000)** | **3,350,000**<br>**(656,000-12,200,000)** |
| Australia | 8,560<br>(4,880-14,000) | 47,800<br>(9,900-89,400) | 9,630<br>(5,750-15,900) | 21,600<br>(10,200-34,600) | 68,300<br>(20,600-110,000) |
| Cambodia | 325<br>(43-629) | 6,850<br>(264-26,700) | 615<br>(64-1,800) | 577<br>(121-1,400) | 14,200<br>(1,570-35,900) |
| China | 849,000<br>(191,000-3,710,000) | 1,480,000<br>(265,000-7,290,000) | 958,000<br>(217,000-3,930,000) | 986,000<br>(219,000-4,380,000) | 2,420,000<br>(329,000-11,700,000) |
| Fiji | 41<br>(9-97) | 492<br>(15-1,860) | 145<br>(10-726) | 71<br>(14-176) | 643<br>(25-1,920) |
| Japan | 89,900<br>(53,700-130,000) | 314,000<br>(105,000-855,000) | 105,000<br>(59,000-173,000) | 163,000<br>(84,900-306,000) | 535,000<br>(152,000-1,250,000) |
| Lao People's Democratic Republic | 86<br>(35-195) | 898<br>(62-4,270) | 154<br>(48-775) | 173<br>(52-445) | 1,540<br>(72-4,960) |
| Malaysia | 1,570<br>(862-2,620) | 15,500<br>(1,610-67,000) | 2,450<br>(1,100-7,630) | 3,050<br>(1,180-6,510) | 27,200<br>(2,620-78,200) |
| Mongolia | 71<br>(2-193) | 1,310<br>(2-6,590) | 616<br>(2-3,590) | 95<br>(2-242) | 1,520<br>(3-6,740) |
| New Zealand | 897<br>(347-1,430) | 6,080<br>(1,050-17,300) | 1,160<br>(420-2,290) | 1,760<br>(686-3,020) | 10,600<br>(1,630-21,100) |
| Papua New Guinea | 240<br>(26-775) | 712<br>(31-3,640) | 324<br>(29-976) | 290<br>(26-866) | 1,380<br>(34-7,330) |
| Philippines | 5,520<br>(4-25,300) | 82,700<br>(12-392,000) | 10,500<br>(4-56,700) | 12,000<br>(10-55,900) | 134,000<br>(36-671,000) |
| Republic of Korea | 6,590<br>(4,150-9,880) | 63,700<br>(6,480-226,000) | 12,800<br>(5,000-65,800) | 12,100<br>(5,830-24,900) | 103,000<br>(9,240-257,000) |
| Singapore | 168<br>(62-293) | 4,020<br>(120-14,100) | 395<br>(78-1,450) | 503<br>(90-1,260) | 7,620<br>(199-18,400) |
| Viet Nam | 419<br>(15-2,590) | 13,000<br>(32-156,000) | 2,620<br>(16-9,620) | 511<br>(15-2,770) | 22,900<br>(57-192,000) |

The largest country in the world, China, also has done one of the best jobs of avoiding a large Omicron wave. As China drops its zero-COVID policy, 1.17 billion (95% UI 0.928–1.35) individuals who have never been infected before will be potentially exposed to an incredibly infectious pathogen. While Omicron-related variants have substantially lower severity than some other variants, given the sheer number of potential infections, our model predicts substantial future COVID-19 burden. In the absence of any new variant emerging, between December 12, 2022, and June 30, 2023, we predict there will be 580 million (276–839) new infections in China. These infections are estimated to result in 2.35 million (1.23–3.77) COVID-19-related hospitalizations and 0.849 million (0.217–3.10) COVID-19-related deaths. It is important to note that these predictions are based on specific assumptions on when (if ever) China would revisit mandate reimposition and should be interpreted as one of many possibilities.

#### Variant scenarios two and three: Omicron-like and Delta-like

A new variant emerging on January 15, 2023, that has similar epidemiological characteristics to Omicron (high infectiousness, moderate severity, high immune-breakthrough) would result in 1.65 billion (95% UI 0.504–3.06) more infections attributable to all variants than the baseline scenario (5.19 billion [3.11–7.78] total) (assuming reference levels of other transmission drivers). Currently circulating variants would still contribute 2.53 billion (1.75–3.56) infections, with the new variant associated with 2.66 billion (0.975–4.56) infections. The moderate severity of this potential future variant would increase total hospitalisations to an estimated 13.6 million (8.5–21.8) new hospitalisations over the forecasted period (an increase of 7.39 million [2.57–12.9]) compared to the baseline scenario) and total deaths to 2.74 million (1.40–5.86) (an increase of 1.15 million [0.360–2.28]; Figure 7d–f, blue lines).

A key difference between our Omicron-like and Delta-like new variant scenarios is the level of immune breakthrough we assume each new variant can achieve (and how this interacts with existing immunity levels due to vaccination and ongoing transmission). To visualize this difference, we consider estimated globally susceptibility to either new variant before the Omicron wave on November 15, 2021 (Figure 11). On that date we estimate 71.0% (69.1–73.0) of the world’s population were susceptible to a new variant with moderate immune breakthrough capacity (similar to Delta, Figure 11a) and 77.6% (75.9–79.3) of the world’s population were susceptible to a new variant with high immune breakthrough capacity (similar to Omicron, Figure 11c). If we re-estimate these quantities after the massive Omicron (and descendants) waves, on December 12, 2022, the population-level susceptibility is much lower for a variant with moderate breakthrough except in China, which avoided the worst of the past 18 months (Figure 11b versus Figure 11d).

Combining a lower capacity to break through past immunity with currently high immunity (and predicted future transmission of currently circulating variants), a new variant emerging with the characteristics of Delta (moderate infectiousness, high severity, moderate immune breakthrough) would have difficulty fully dominating transmission in all locations. If a new variant with Delta-like characteristics emerged on January 15, we predict there would be 3.64 billion (95% UI 2.26–5.83) new infections between December 12, 2022, and June 30, 2023, but 3.22 billion (2.17–4.59) of those would be attributable to currently circulating variants, with 423 million (39.4–1340) attributable to the new variant. Although Omicron is substantially less severe compared to Delta, due to the incomplete global dominance of a Delta-like variant, hospitalisations are predicted to be lower in the Delta-like variant scenario than the Omicron-like new variant scenario: 7.87 million (4.81–13.0) compared to 13.6 million (8.5–21.8), respectively. Total deaths, for which the severity difference between Delta and Omicron is greater, are comparable between the two scenarios, with a predicted 2.87 million (1.03–5.56) new deaths in the Delta-like variant scenario (Figure 7d–f, green lines).

#### Variant scenarios four and five: Enhanced Delta-like and DeltaCron

Both our enhanced Delta-like scenario and our DeltaCron scenario assume high severity and high immune breakthrough. DeltaCron goes one step further by assuming the new variant is as infectious as Omicron. Both scenarios are simulating the spread and impact of a new pathogen, and both could be considered as worse-case scenarios (with DeltaCron being “even worse”). Even with only moderate infectiousness, due to its high immune breakthrough, an enhanced Delta-like new variant would cause a massive new wave of COVID-19 burden, resulting in an estimated 4.50 billion (95% UI 2.69–7.18) infections, 21.3 million (9.54–37.2) hospitalisations, and 11.1 million (2.67–22.0) deaths; Figure 7d-f, purple lines). As expected, DeltaCron would match the infection numbers of the Omicron-like new variant (5.19 billion [3.11–7.78]) but would result in catastrophic morbidity and mortality. In the extremely unlikely scenario that fundamental changes would be made to reduce transmission (eg, a repeat of the lockdowns of early 2020), DeltaCron would result in 30.2 million (13.4–51.2) new hospitalisations and 15.9 million deaths (4.31–35.9, Figure 7d-f, red lines). Detailed results for all five variant scenarios by both WHO region (Figure 8) and for all national and subnational locations are available in Appendix 3.

**Figure 8.**
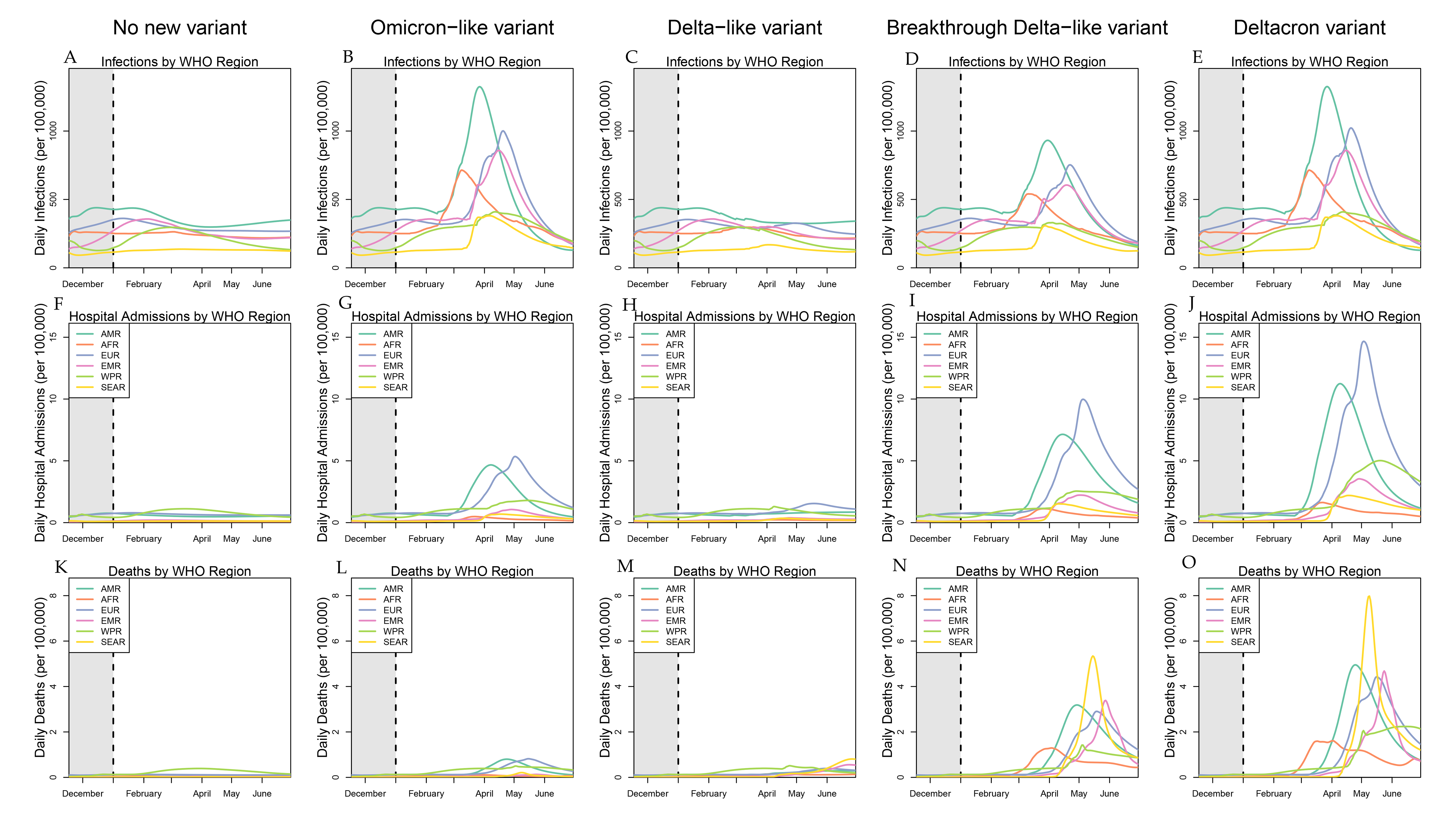
Daily infections, hospitalisations, and death rates under five future variant scenarios by WHO region. The first column (A, F, and K) displays daily SARS-CoV-2 infections, COVID-19 hospital admissions, and COVID-19 deaths by WHO region for the no new variant future scenario. The second column (B, G, and L) displays similar information for “Omicron-like” variant scenario, the third column (C, H, and M) displays similar information for “Delta-like” variant scenario, the fourth column (D, I, and N) displays similar information for a Breakthrough “Delta-like” variant scenario, and the fifth column (E, J, and O) displays similar information for DeltaCron scenario. AMR = Region of the Americas. AFR = African Region. EMR = Eastern Mediterranean Region. EUR = European Region. SEAR = South-East Asia Region. WPR = Western Pacific Region.

#### Three potential future intervention scenarios

##### Intervention scenario one: historical high mask usage

The mask scenario—in which mask use increases to 80% of the population or the location-specific current high, if higher—averted at least 15% of forecasted deaths across variant scenarios and regions. At the global level, depending on the characteristics of the future variant, deaths were reduced by 23.0% (95% UI 14.1–33.8) to 28.1% (16.2–40.8) compared to reference levels of transmission drivers (Figure 10a). For the baseline variant scenario, this translated into 339,000 averted deaths (160,000–709,000) over the forecasted period, while for the DeltaCron scenario, over 3.23 million (1.16–8.21) deaths would be averted (Figure 9). Masks had the greatest impact in the Delta-like new variant scenario where the new variant struggles to compete, averting 909,000 (231,000–1,240,000) deaths (Figure 9). Across all variant scenarios, the lowest impact of mask levels returning to their previously observed high was in the Western Pacific Region; for the DeltaCron scenario, increased mask usage reduced deaths in the Western Pacific Region by only 15.2% (6.84–24.6), Figure 10). Irrespective of the variant scenario, increased mask usage had the largest impact in the European Region, where mask use was particularly low at 12.3% as of December 12, 2022, averting a minimum of 38.6% (23.0–55.4) of deaths in the DeltaCron new variant scenario and up to 44.1% (26.6–63.2) of deaths in the Breakthrough Delta new variant scenario. Mask use was also very effective in the South-East Asia and Eastern Mediterranean regions, averting at least 30% of deaths across all variant scenarios. In contrast, the increased mask use intervention consistently had the smallest impact in the Western Pacific Region, where mask use was quite high at 60.7% at the beginning of the forecast period, averting a maximum of 17.2% (8.83–30.6) of deaths in our baseline scenario where no new variant invades (Figure 10).

**Figure 9.**
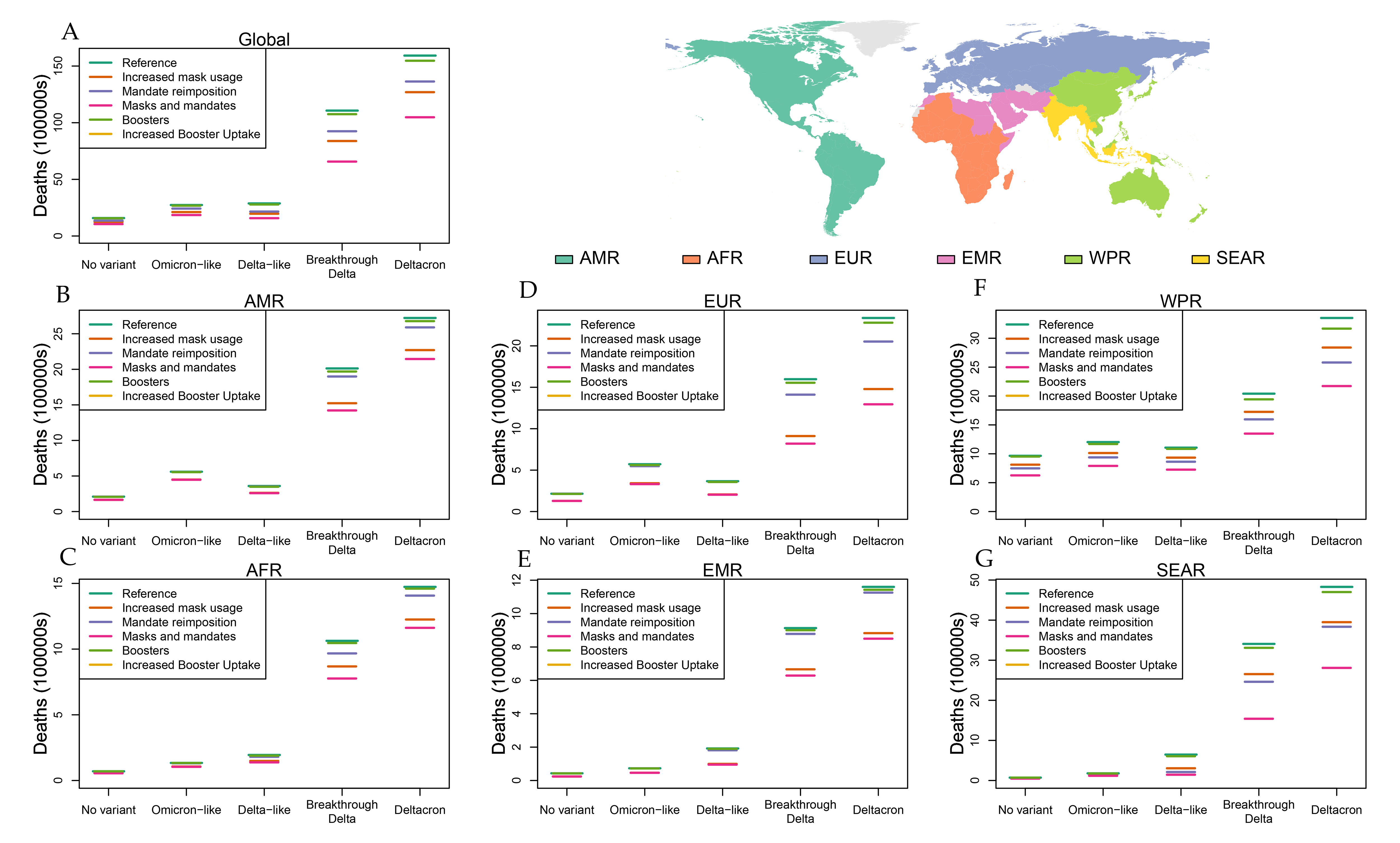
Total COVID-19 deaths December 13, 2022, through June 30, 2023, across four intervention scenarios, globally and by WHO region. Panel A: global cumulative COVID-19 deaths from December 13, 2022, through June 30, 2023, are plotted for each of the five variant scenarios. For each variant scenario, reference (green line) is comparable to the increased mask usage scenario (orange line), globally distributed antivirals (purple line), and boosters for all previously vaccinated individuals who are not already boosted (magenta line). Similar information is presented in Panels B-G for each WHO region. The upper and lower whiskers of each line depict 95% uncertainty intervals. AMR = Region of the Americas. AFR = African Region. EMR = Eastern Mediterranean Region. EUR = European Region. SEAR = South-East Asia Region. WPR = Western Pacific Region.

**Figure 10.**
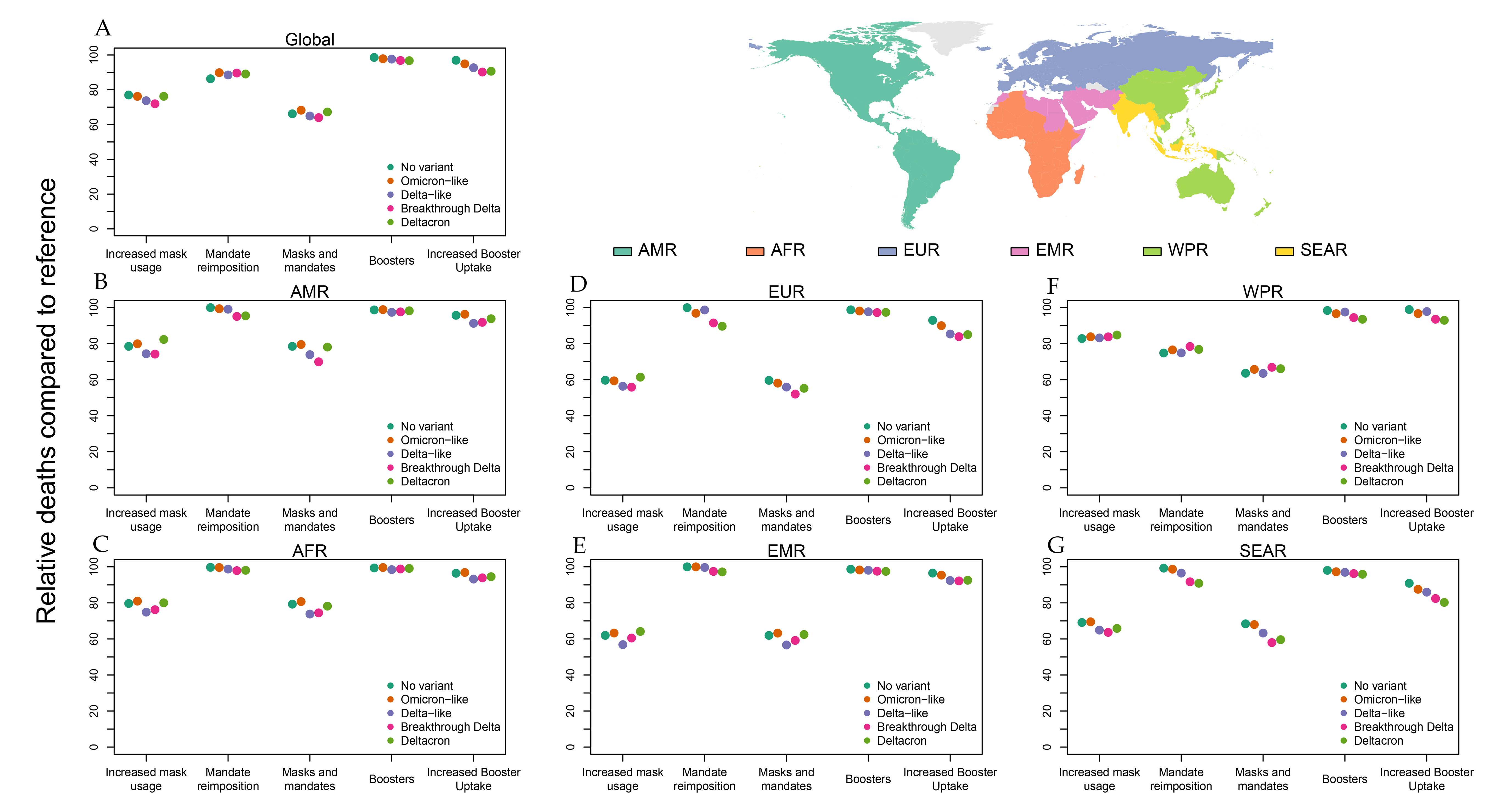
Reduction in COVID-19 deaths by intervention scenario over forecasted period compared to the reference intervention scenario, globally and by WHO region. Panel A: global relative reduction in COVID-19 deaths from December 13, 2022, through June 30, 2023, are plotted for each of the three intervention scenarios relative to the reference scenario across the five variant scenarios. For each intervention scenario (left: Increased mask usage, middle: globally distributed antivirals, right (boosters for all previously vaccinated individuals who are not already boosted), relative reductions within each variant scenario compared to the reference scenario are plotted for no new variant (dark green), “Omicron-like” new variant (orange), “Delta-like” new variant (purple), Breakthrough “Delta-like” variant (red), and DeltaCron (light green). Similar information is presented in Panels B-G for each WHO region. The upper and lower whiskers of each line depict 95% uncertainty intervals. AMR = Region of the Americas. AFR = African Region. EMR = Eastern Mediterranean Region. EUR = European Region. SEAR = South-East Asia Region. WPR = Western Pacific Region.

##### Intervention scenario two: Mandate re-imposition

As described in the methods, the mandate re-imposition intervention reflects policy mandate levels in the post-Ancestral era rather than the severe lockdowns imposed by much of the world at the outset of the pandemic in 2020, as we believe this better represents the level of intervention governments are willing to impose as the pandemic nears its third year. The scenario also requires that a country or subnational unit reach a high daily death rate relative to rates observed in the Omicron era (when mandate levels in most locations hit historic lows) before mandates are reimposed. As such, this intervention is a conservative one along a spectrum of potential algorithms that could be used to trigger new mandates. In many regions and variant scenarios, this means the mandate reimposition intervention is only mildly to moderately effective. In the minimum case, mandates are never reimposed in the Eastern Mediterranean Region in our baseline scenario where no new variant is introduced. On the other hand, in the Western Pacific Region, mandate reimposition averts at least 21% of deaths in all scenarios, primarily due to the presumed impact of China reverting to lockdowns in the presence of a massive surge in COVID infections and deaths (averting at least 30% of deaths in every variant scenario; Figure 10f). Importantly, our mandate reimposition intervention is a discontinuous one, occurring only in the fraction of our forecast trajectories where outcomes are most severe. That is, in a given country and variant scenario, only a fraction of the distribution forecast trajectories we generate will reach the threshold for mandate reimposition in that location, which means our distributions of averted deaths have a large weight at zero for many regions and drastically shrink the lower bound of our predicted impact size.

Globally, mandate reimposition matters most in absolute terms in the DeltaCron variant scenario, where it averts 2.29 million (95% UI 0.924–9.69) deaths (Figure 9), the majority of those deaths coming from the Western Pacific Region (771,000 [1450–3.02 million]; Figure 9f) and the South-East Asia Region (994,000 [0–4.71 million]; Figure 9g) with China (724,000 [0–2.93 million]) and India (957,000 [0–4.70 million]) delivering the bulk of the impact in those regions, respectively. In the Western Pacific Region, mandate reimposition always matters, averting at a minimum of 21.6% (0–47.8) of deaths in the Breakthrough Delta scenario and up to 25.2% (0–64.8) of deaths in the baseline scenario (Figure 10f). Mandate reimposition has a minor to negligible effect in other regions due primarily to a historical hesitancy to implement strong mandates in the face of severe COVID outbreaks (Figure 10).

#### Intervention scenario three: variant-targeted boosters

One of the benefits of mRNA vaccines is the faster turnaround from pathogen/variant/strain identification and the production of a vaccine. For a new vaccine specifically designed to target a newly emerged variant, however, the speed of development and distribution would have to compete with the speed at which the variant spread. The Omicron variant spread globally very quickly, and in the absence of international travel restrictions, we assume a new variant would spread at the same speed. The second challenge associated with a new vaccine would be convincing individuals at risk to take it. In almost every setting, the hesitancy to take a booster against SARS-CoV-2 was substantially larger than for the first round of doses. In our base booster intervention scenario, we assumed uptake would match that observed for the first booster dose. In this intervention scenario, we found very little impact of the targeted booster, with global reductions in COVID-19 mortality no greater than 3.28% (1.86–4.85), a reduction of 453,000 (169,000–1.34 million) deaths in the DeltaCron variant scenario (Figure 10a, Figure 9a). The largest regional impact was in the Western Pacific Region, where a targeted booster averted 6.47% (2.61–10.3) of deaths in the DeltaCron scenario (Figure 10f). In all other regions, the maximum relative impact of a fast but historically plausible vaccine rollout is less than 5% in all variant scenarios (Figure 10).

We also considered an idealistic second booster scenario, where we recalculated future booster uptake by imagining every country in a WHO region would achieve the highest uptake levels seen in any country in that region. While it would take significant effort to increase uptake to these levels, it nearly triples the impact at the global scale in all variant scenario, with a maximum relative reduction of 9.79% (95% UI 5.36–14.9) or 995,000 (241,000–3.05 million) deaths in the Breakthrough Delta scenario (Figure 10a, Figure 9a). The South-East Asia Region sees a mild to moderate impact in all variant scenarios, ranging from a mortality rate reduction of 9.13% (5.07–12.7) in the baseline scenario to a reduction of 19.7% (5.78–33.4) in the DeltaCron scenario (Figure 10g). The impact is also notable in the European Region, where a fast scale-up of targeted boosters can reduce mortality by up to 16.1% (10.6–21.9) under a Breakthrough Delta-like variant (Figure 10d). The impact is more muted in other regions, with a maximum mortality rate reduction of 6.78% (2.81–14.5) in the African Region with a Delta-like variant, 7.82% (3.85–12.1) in the Eastern Mediterranean region with a Breakthrough Delta-like variant, 8.73% (3.97–15.7) in the Region of the Americas with a Delta-like variant, and 7.05% (1.8–12.6) in the Western Pacific Region with a DeltaCron variant (Figure 10).

#### Combined interventions

Focusing on the two interventions with the largest impact (increased mask usage and mandate re-imposition), we further investigate the impact of these two interventions deployed simultaneously. This intervention scenario has similar caveats to the mandate re-imposition scenario as mandates won’t be re-imposed in many of our forecast trajectories unless things get quite severe—the distributions of outcomes here have a large weight at the level where only the mask use intervention is implemented. This effect is even larger here as mask use alone mitigates many of the more severe outcomes in most situations. At the global level, together these interventions avert between 31.8% and 36.0% of all projected future COVID-19 deaths by June 30, 2023 (Figure 10a). The largest relative impact is seen if the new emerging variant has characteristics similar to Delta with increased breakthrough, with 4.50 million (95% UI 0.733–15.2) deaths averted, while even in the most cataclysmic case of DeltaCron, this combined intervention scenario is predicted to avert 5.44 million (1.20–17.5) deaths. Across all regions, in every variant scenario this combined intervention averted at least 19% of future deaths, with significantly larger effects in most regions (a minimum mortality rate reduction of 19.3% [8.45–34.3] in the African Region; 37.5% [15.2–61.6] in the Eastern Mediterranean Region; 40.4% [26.6–54.4] in the European Region; 20.5% [12.0–32.5] in the Region of the Americas; 31.6% [13.9–54.3] in the South-East Asia Region; and 33.1% [11.4–63.0] in the Western Pacific Region). For projected hospitalisations under each intervention scenario across all variant scenarios, see appendix 1, section 13 and appendix 3, tables S2, S4, S7, S10, S13, and S1.

## Discussion

The ongoing Omicron waves have likely provided humanity with the additional immunity necessary to shift COVID-19 from an epidemic to an endemic infection. The extraordinary spread of Omicron contributed to 8.47 billion (95% UI 6.20–11.6) new infections from November 15, 2021, through December 12, 2022. While this period of the COVID-19 pandemic resulted in the loss of an estimated 3.04 million (2.65–3.55) lives directly attributed to SARS-CoV-2 infection, it has added to previous exposure due to past infection and vaccination, resulting in an estimated 6.39 billion (6.05–6.58) individuals (83.2% [81.6–84.9]) of the global population) having gained some level of immunity to SARS-CoV-2 as of December 12, 2022. This immunity, while varying in strength due to vaccine efficacy or the recency of past infections, is directly responsible for the relatively low forecasted COVID-19 burden across all but the worst-case scenarios we considered.

As past immunity wanes and individuals choose to participate in more risky behavior (such as not wearing masks), we expect transmission to continue. Even under a scenario in which no new variants emerge between December 12, 2022, and June 30, 2023, we estimate more than 3.54 billion (95% UI 2.24–5.43) new infections and 1.58 million (0.829–3.95) deaths, moderately lower than the 2.08 million (1.80–2.44) deaths observed from December 12, 2021, to June 30, 2022, the same period a year earlier. If another Omicron-like variant emerges, a scenario we view most likely, we expect instead 2.74 million (1.40–5.86) deaths in the next six months, a moderate increase from the same period a year ago. This increase is primarily explained by a much lower vaccine-derived protection to severe COVID outcomes, though overall lower mask use and mandate levels also contribute. In the situation where a variant with Delta-like severity emerges, the outcomes range from a moderate increase in deaths (2.87 million [1.03–5.56]), comparable to an Omicron-like variant, to a potentially massive surge in deaths (15.9 million [4.31–35.9]) if the variant also shares the transmission and breakthrough characteristics of an Omicron-like variant.

While ongoing transmission of currently circulating variants will continue to add to the burden associated with COVID-19 and SARS-CoV-2, this endemic transmission setting does provide some collateral population immunity benefits. The currently circulating variants are considerably less likely to result in severe outcomes compared to the Delta variant,^45^ and by continually infecting, mutating, and then breaking through past immunity, these variants appear to maintain higher levels of immunity to observed and potential variants with less capacity to break through immunity. In our Delta-like new variant scenario, we saw the new variant had difficulty in monopolising transmission in most locations, and although an infection with this hypothetical new variant was more likely to result in morbidity and mortality, the overall estimated burden was similar to what we predict if another new Omicron-like variant emerges. It is, of course, possible, if a variant that is more severe also develops a high capacity to break through previous immunity, as can be seen in our Breakthrough Delta and DeltaCron variant scenarios, the predicted burden will still be catastrophic, without substantial policy interventions.

Given the number of susceptible individuals in China, and the recent change in policy to move away from a zero-COVID strategy, there is potential for a massive COVID-19 wave in China in the next month. Many sources indicate this acceleration of infections is beginning.^46^ In our most conservative scenario, we predict 2.35 million (95% UI 1.23–3.77) COVID-19-related hospitalisations and 849,000 (217,000–3,100,000) COVID-19-related deaths. It is crucial to note that although we investigate a handful of intervention scenarios, these are just a small subset of the possible approaches that the Chinese government could take to prevent a massive loss of life.^46^ Antivirals have proven effective in reducing the most severe outcomes and should be acquired and distributed to clinics and hospitals for administration to those most at risk. While mask use is already high in China, mandates that increase usage even higher will likely slow the potential oncoming wave. Targeted social distancing mandates have proven immensely successful in China in the past and could again be used in the hardest hit areas to further slow the rise in SARS-CoV-2 transmission. Finally, if the predicted wave can be slowed by other measures, a national booster policy may be able to further protect individuals. Many of the Chinese population were vaccinated over six months ago, so the protection they have derived from those vaccinations has likely greatly waned. Implementing a larger booster program, especially with more effective mRNA vaccines, could have substantial impact if other measures can hold off the worst until such a campaign could be mobilised. Given the immense uncertainty in both how China may react to the future threat as well as what may come in terms of future variants, there is a pressing need for careful surveillance of the current outbreaks in China, alongside the full repertoire of proven interventions to avoid substantial loss of life.

While increased mask usage did not eliminate the future risk from a newly emerging variant, in every scenario considered, resuming mask usage averted substantial burden, with average reductions of at least 15% (Figure 10). While reductions for a moderate mandate reimposition scenario were lower, the combination of these two intervention packages averted more deaths than each one alone. It is critical to note that the mandate reimposition scenario was more conservative that what was seen in 2020, and as such, if a new threat is detected, policy makers should reconsider mandate reimpositions as a viable tool to reduce transmission and avoid overwhelming the health-care system. In settings where the new variant is deadly and very infectious, combining masks and mandates can have dramatic impacts, reducing burden by up to 44.8% (95% UI 28.7–61.6), as seen in the European Region.

Our vaccine-specific booster scenarios attempted to leverage the speed at which mRNA vaccines can be developed. They took optimistic estimates for development, distribution, and uptake, but due to the incredibly fast rate at which Omicron spread globally (and our assumption that a new variant would spread equally as fast), the impact of these intervention scenarios was minimal as the majority of the impact of the new variant had already occurred before the booster reached market. In several specific locations, assuming an accelerated uptake schedule had moderate impact in averting burden in some variant scenarios. It is unclear that a new variant would spread as fast as Omicron, but in the absence of companion interventions that delay the spread or slow the location-specific invasion of a new variant, mRNA vaccines alone are unlikely to be available fast enough to blunt the first wave of a new variant.

There are several important limitations that must be considered when evaluating the outcomes of this and any infectious disease forecast at this time scale. There are several components of a model of this level of complexity, and the assumptions of each component, as well as how those components interact that can and will influence the robustness of conclusions associated with the model results. These are documented most completely in the appendix. First, there are critical assumptions associated with input data fidelity. For example, the vaccine efficacy tables presented in appendix 1 table S2 cannot be exclusively created from data, as there have not been clinical trials run on every vaccine–variant combination. Moreover, several parameters within the transmission dynamic model are poorly identified in isolation, such as variant-specific relative increases in infectiousness versus probability of breaking through past immunity. Second, the relative contribution of immunity escape versus increases in transmissibility of those already infected by some variants of concern is difficult to parse. Third, we encountered several limitations related to seroprevalence data, including that some surveys may be biased in a direction that is difficult to determine, and that we attempted to standardise for various corrections that had been applied to serosurvey estimates but could not always do so. These limitations are detailed in Barber and colleagues.^12^ Fourth, we used an estimate of the proportion of excess mortality attributable directly to SARS-CoV-2 infection to estimate COVID-19 deaths. This strategy is a response to an absence of reliable and complete cause-specific death data but yields highly uncertainty estimates (reflected in our 95% UIs).^12^ Fifth, due to data limitations, it is important to note that the forecasted burden estimates presented here ignore the potential reduction due to fourth doses (or second boosters) of vaccines. Other scenarios, such as a more targeted distribution of vaccine doses to those locations with the highest need, would provide insight on alternative approaches to preventing future loss of life and should be considered. Finally, and as stated above, our scenarios represent 30 specific potential realities, and the corresponding results for each scenario must be viewed as illustrating a range of possible outcomes. It is this universe, when considered with a very wide range of additional socioeconomic factors, in which policy makers must weigh the most prudent actions on behalf of the populations they represent.^24^

There are myriad directions of future work that would help us reduce future COVID-19 burden as well as prepare for the next pandemic. Our model currently fits a flexible transmission intensity trajectory for each location to past data, but there is an opportunity for a more detailed analysis of the drivers of the observed variation in transmission. Future work must look more specifically at the estimated impacts of non-pharmaceutical interventions, messaging and trust in local governmental officials and “science” in general, as well more broadly at the impact of human behaviour on transmission dynamics. Even though we cannot change the past, understanding which locations reacted to the pandemic well, which reacted poorly, which mandates were life-saving, and which recommendations may have hurt more than they helped will be important to help us prepare for the next pandemic. While the IHME COVID-19 forecasting model was one of a handful of models that forecasted the COVID-19 pandemic at the global level, there were hundreds of models that made projections of COVID-19 burden for some set of locations. Understanding what makes a “good” model (or what modelling approaches are better at short- or long-range forecasts) will be key for creating better ensemble models for future pandemics, and as such, systematic comparisons of all available COVID-19 models should be conducted. Finally, as exemplified by our baseline scenario forecasting more than 3 billion new infections, the disease burden associated with COVID-19 will continue to grow. Careful monitoring will be necessary to track and react to changes in COVID-19 epidemiology, and it is likely that more modelling alterations will be needed to incorporate whatever future comes next.

## Conclusion

There is considerable uncertainty in how future variants may alter the trajectory of the COVID-19 pandemic. In this analysis, we considered a range of future scenarios that reflect the potential emergence of new variants, in the context of several different intervention approaches. Paradoxically, the world has become better protected from COVID-19 due to the surge of infections associated with Omicron. As such, only in our “worst-case scenarios” did we did see a return to the global mortality levels seen over the same period in 2021. We found that regardless of the severity and transmissibility of existing and future variants, expedited variant-specific mRNA boosters had minimal impact due to the speed at which variants spread globally. High mask usage resulted in strong reductions, and moderate mandate reimposition strategies likewise reduced future burden; the combination of these two interventions had a greater impact than each alone. In addition to what future variants may emerge, how the current COVID-19 surges in China will unfold is uncertain, but it is clear there is potential for a mass loss of life. China, like the rest of the world, does have multiple tools at their disposal to reduce the worst of what is to come, but they will need to act quickly and decisively. Our analysis has shown that simple interventions, such as mask use, are expected to remain vital tools in our battle against COVID-19. As COVID-19 looks to be transitioning to a more endemic transmission regime, 30 plausible future scenarios remain an important piece of information for policy makers and the public alike.

## Data Availability

All code used in the analysis can be found online https://github.com/ihmeuw/covid-model-seiir-pipeline.

---

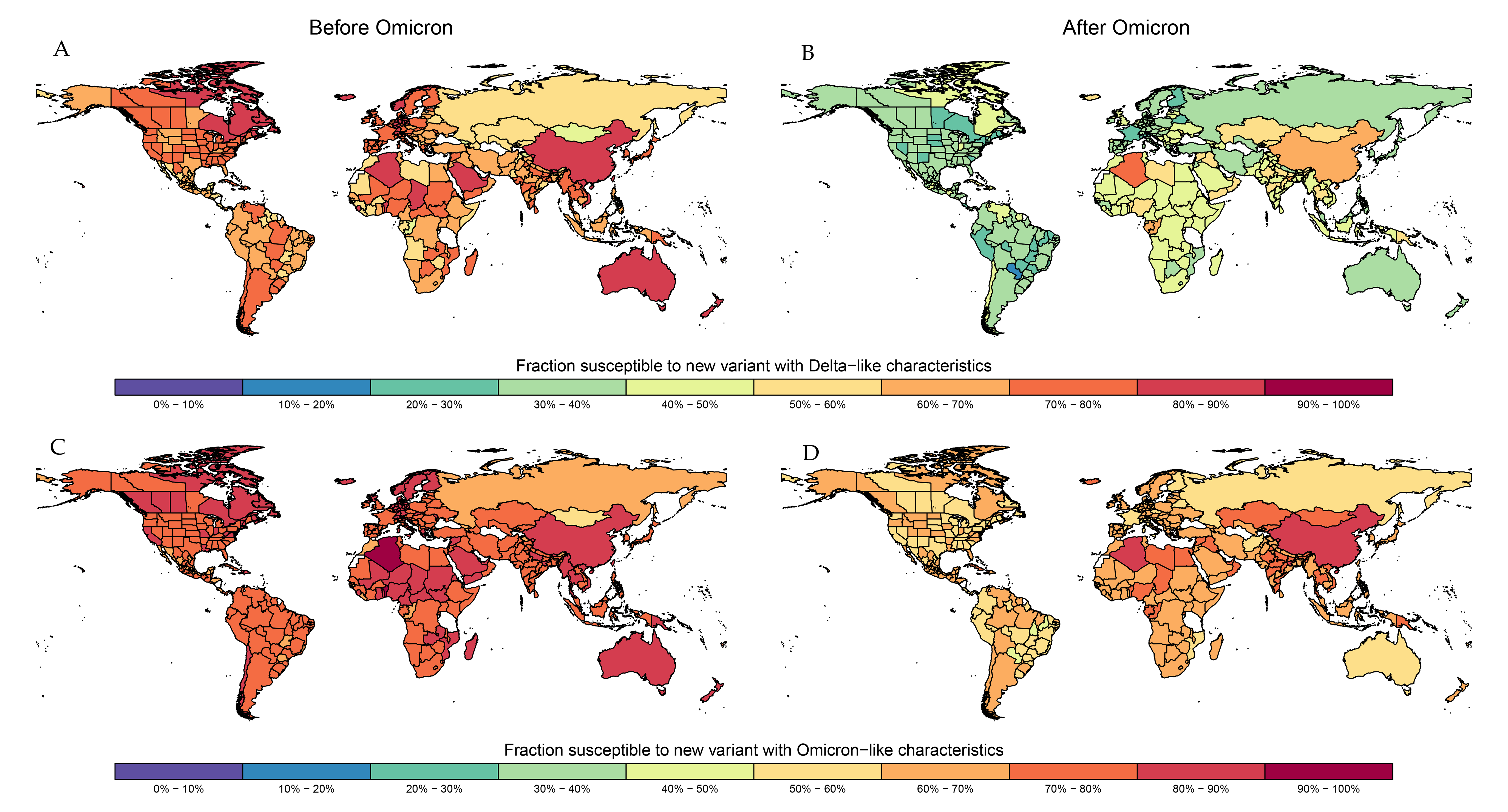

