## Appendix 1 for "Forecasting the trajectory of the COVID-19 pandemic into 2023 under plausible variant and intervention scenarios: a global modelling study"

Appendix 1: Supplementary methods and results for “Forecasting the trajectory of the COVID-19 pandemic into 2023 under plausible variant and intervention scenarios: a global modelling study”

This appendix provides further methodological details and supplementary results for “Forecasting the trajectory of the COVID-19 pandemic under plausible variant and intervention scenarios: a global modelling study”

Portions of this appendix have been reproduced or adapted from Reiner et al.^1^ and Barber et al.^2^ References are provided for reproduced sections.

### Section 1 Table of Contents

### List of abbreviations

| Abbreviation | Full phrase |
| --- | --- |
| B.1.1.7 | SARS-CoV-2 Alpha variant |
| B.1.351 | SARS-CoV-2 Beta variant |
| B.1.617.2 | SARS-CoV-2 Delta variant |
| COVID-19 | coronavirus disease 2019 |
| CDC | Centers for Disease Control and Prevention |
| GATHER | Guidelines for Accurate and Transparent Health Estimates Reporting |
| GBD | Global Burden of Diseases, Injuries, and Risk Factors Study |
| GHDx | Global Health Data Exchange website |
| GISAID | global initiative on sharing avian influenza data |
| IDR | infection–detection ratio |
| IFR | infection–fatality ratio |
| IHR | infection–hospitalisation ratio |
| J&J vaccine | Johnson & Johnson Janssen COVID-19 vaccine |
| LRI | lower respiratory infection |
| MR-BRT | meta-regression—Bayesian, regularised, trimmed |
| NPI | non-pharmeceutical intervention |
| P.1 | SARS-CoV-2 Gamma variant |
| PANGO | Phylogenic Assignment of Named Global Outbreak |
| R_effective_ | effective reproduction number |
| RT-PCR | reverse transcription-polymerase chain reaction |
| SARS-CoV-2 | severe acute respiratory syndrome coronavirus 2 |
| SEI | susceptible-exposed-infectious |
| UI | uncertainty interval |
| WHO | World Health Organization |

### Section 2 Guidelines for Accurate and Transparent Health Estimates Reporting (GATHER)

This study complies with GATHER recommendations.^3^ We have documented the steps in our analytical procedures and detailed the data sources used. See table S1 for the GATHER checklist. The GATHER recommendations can be found on the [GATHER website](http://gather-statement.org/).

Table S1 GATHER checklist

| Item # | Checklist item | Reported location |
| --- | --- | --- |
| Objectives and funding | | |
| 1 | Define the indicator(s), populations (including age, sex, and geographic entities), and time period(s) for which estimates were made. | Main text methods: “time periods, locations, age groups, and outcome measures” |
| 2 | List the funding sources for the work. | Main text summary & acknowledgments |
| Data Inputs | | |
| For all data inputs from multiple sources that are synthesized as part of the study: | | |
| 3 | Describe how the data were identified and how the data were accessed. | Main text methods “estimating and forecasting key model drivers”; detailed in appendix 1, sections 4, and data source sections within section 6 |
| 4 | Specify the inclusion and exclusion criteria. Identify all ad-hoc exclusions. | Appendix 1 |
| 5 | Provide information on all included data sources and their main characteristics. For each data source used, report reference information or contact name/institution, population represented, data collection method, year(s) of data collection, sex and age range, diagnostic criteria or measurement method, and sample size, as relevant. | Appendix 1, section 4; appendix 2; link to GHDx will be added at acceptance. |
| 6 | Identify and describe any categories of input data that have potentially important biases (e.g., based on characteristics listed in item 5). | Appendix 1, section 6; main text limitations section |
| For data inputs that contribute to the analysis but were not synthesized as part of the study: | | |
| 7 | Describe and give sources for any other data inputs. | N/A |
| For all data inputs: | | |
| 8 | Provide all data inputs in a file format from which data can be efficiently extracted (e.g., a spreadsheet rather than a PDF), including all relevant meta-data listed in item 5. For any data inputs that cannot be shared because of ethical or legal reasons, such as third-party ownership, provide a contact name or the name of the institution that retains the right to the data. | GHDx URL will be added at acceptance; all data inputs can be downloaded there |
| Data analysis | | |
| 9 | Provide a conceptual overview of the data analysis method. A diagram may be helpful. | Main text methods sections: “overview of the SEI transmission model” and “estimating and forecasting key model drivers” |
| 10 | Provide a detailed description of all steps of the analysis, including mathematical formulae. This description should cover, as relevant, data cleaning, data pre-processing, data adjustments and weighting of data sources, and mathematical or statistical model(s). | Appendix 1, sections 4–9 |
| 11 | Describe how candidate models were evaluated and how the final model(s) were selected. | Appendix 1, section 11 |
| 12 | Provide the results of an evaluation of model performance, if done, as well as the results of any relevant sensitivity analysis. | Appendices 4 and 5 |
| 13 | Describe methods for calculating uncertainty of the estimates. State which sources of uncertainty were, and were not, accounted for in the uncertainty analysis. | Main text methods section “uncertainty estimation”; appendix 1 section 10 |
| 14 | State how analytic or statistical source code used to generate estimates can be accessed. | Github link provided in main text methods “GATHER compliance” section |
| Results and Discussion | | |
| 15 | Provide published estimates in a file format from which data can be efficiently extracted. | Online data tools (data visualization tools, data query tools, and the Global Health Data Exchange, link to the GHDx to be provided upon acceptance) |
| 16 | Report a quantitative measure of the uncertainty of the estimates (e.g. uncertainty intervals). | All quantitative measures are reported with 95% UIs, including in the results section, tables, and figures (as appropriate) |
| 17 | Interpret results in light of existing evidence. If updating a previous set of estimates, describe the reasons for changes in estimates. | Main text RIC section “implications of all the available evidence,” and discussion |
| 18 | Discuss limitations of the estimates. Include a discussion of any modelling assumptions or data limitations that affect interpretation of the estimates. | Main text methods and discussion and appendix 1 |

### Section 3 Primary model inputs^2^

#### Section 1.1 Reported cases data

Data on reported cases primarily came from Johns Hopkins University,^4^ supplemented by location-specific datasets extracted either directly from ministries of health, departments of public health, or other third parties. Sources are outlined in appendix 2, section 8. Cases were defined, depending on the local context, as either an individual who has received a positive test result, whether RT-PCR, antigen, or antibody (regardless of symptoms status) or an individual who has symptoms consistent with a clinical definition of COVID-19. Adjustments to the time series were periodically required, either to account for interruptions in daily reporting due to, for instance, major public holidays, or more systematic issues, such as reporting backlogs of cases accumulated in laboratory processing, or adjustments due to changes in case definitions. A catalogue of these corrections is available through the associated GHDx record.

#### Section 1.2 Hospital admissions data

Data on reported daily admissions, or cumulative hospitalisations, was typically sourced from ministries of health, or multi-jurisdiction agencies such as the US Department of Health and Human Services (HHS), or the European Centres for Disease Control. Sources are outlined in appendix 2, sections 3, 4, 5, 6, and 7. Adjustments to the time series were periodically required, either to account for interruptions in daily reporting due to, for instance, major public holidays, or more systematic issues, such as changes in COVID case definitions. A catalogue of these corrections is available through the associated GHDx record.

For time series that were incomplete for the entirety of the pandemic, such as the US HHS admissions dataset, we imputed the missing time-series portions by first running separate linear regressions for each location with missingness. The dependent variable was admissions, and the independent variable was infections derived from both daily deaths divided by the infection–fatality ratio (IFR)^5^ and from cases divided by the infection–detection ratio (IDR) for the period in time that overlapped with the admissions data. We then used the coefficient from this model, a naïve infection–hospitalisation ratio (IHR) estimate, to predict out of sample for the period that was missing admissions, using the average of the two separately derived infections estimates. To avoid any disjoints at the day of transition from imputed data to the observed, we gradually transitioned from the former to the latter over the tail period of the imputation. We did this by determining the ratio of the average observed admissions over first week of data to the predicted admissions for that week, linearly interpolated from a ratio of 1 at 60 days before observed to the calculated residual ratio at the first day of observed and multiplied the imputation model predictions by that ratio during that period. We then included the imputed admissions along with the observed in our hospitalisations database.

#### Section 1.3 Reported deaths data

Data on reported daily deaths primarily came from Johns Hopkins University,^4^ supplemented by location-specific datasets extracted either directly from ministries of health, departments of public health, or other third parties. Sources are outlined in appendix 2, sections 1 and 2. Adjustments to the time series were periodically required, either to account for interruptions in daily reporting due to, for instance, major public holidays, or more systematic issues, such as reporting backlogs of deaths accumulated in vital registration system processing, or adjustments due to changes in case definitions and reconciliation of death certificates. A catalog of these corrections is available through the associated GHDx record.

#### Section 1.4 Seroprevalence data

Data on serosurveys reporting antibody positivity were collated on an ongoing basis. Two key data types were tracked—ongoing serological surveys conducted by governmental organisations and released periodically, and publications of antibody surveys published in preprint servers and traditional journals. For the latter, we leveraged existing published reviews^6,7^ and cross-referenced the Serotracker database.^8^ Sources used in this study and used as location-representative studies are outlined in appendix 2, sections 9 and 10. Data that were deemed not to be representative of the general population in the most-detailed geographical location in our modelling hierarchy covering the study site were excluded. Additionally, we excluded studies that reported less than 0.03 seroprevalence, as we found that measurements taken with little signal in the population resulted in empirical estimates of IFR not generalisable across locations, or even within location after broader exposure of the population.

### Section 2 Historical ratio estimation

Detailed descriptions of methods for estimating the infection–detection ratio (IDR), infection–hospitalisation ratio (IHR) and infection–fatality ratio (IFR) can be found in Barber et al.^2^ A summary of the current adaptation is provided here. Seroprevalence surveys from ongoing government surveillance programs and from preprint and peer-reviewed studies were collated. Surveys were adjusted to account for vaccination rates, where appropriate, and for decaying sensitivity over time due to seroreversion. These data were paired with cumulative cases, hospitalizations, and deaths to produce empirical estimates of the IDR, IHR, and IFR, respectively. Case, hospitalization, and death data were first adjusted to only be representative of individuals with no existing immunity from prior infection or vaccination using methods described in section 7.1. Models for each ratio were then fit in a cascading meta-regression framework using MRTool – an open-source Bayesian meta-regression framework developed by IHME^9,10^ – based on various combinations of predictive covariates listed in section 5. The model of IFR also included a time variable to capture the effect of improved treatment over time, and both the IFR and IHR models included a term to control for the severity risk ratio of non-ancestral variants relative to ancestral given the cumulative variant prevalence at the time of the survey. Predictions were made the variant effect, thus representing the ratio associated with ancestral infections among the unvaccinated with no prior infection. These predictions, made for all location-dates in the analysis, serve as the prior estimates of IDR, IHR, and IFR in the estimation of past infections described in section 7.

### Section 3 Model drivers

#### Section 3.1 Variants

Our base model of the invasion date and the rate of invasion of several severe acute respiratory syndrome coronavirus 2 (SARS-CoV-2) variants was based on sequence data from the GISAID Initiative^11^. The resulting models were then manually calibrated against reported case, hospitalisation, and death data in each location. The invasion dates for the new variant in our scenarios were determined by shifting the Omicron invasion dates forward so that South Africa is invaded on January 15, 2023.

##### Section 3.1.1 Data sources

Data on variants was primarily sourced from the GISAID Initiative.^11^ This group provides a centralized mechanism for the collation and curation of sequences of SARS-CoV-2 viruses. Key metadata are tracked, including date of specimen collection, and Phylogenic Assignment of Named Global Outbreak (PANGO) lineage assignment. For each estimation location, we track the daily number of SARS-CoV-2 sequences collected (starting from January 1, 2020), and the number of variant-specific sequences collected on that day. We specifically track: B.1.1.7 (Alpha), B.1.351 (Beta), P.1 (Gamma), B.1.617.1 (Kappa), and B.1.617.2 (Delta) using sequence data.

Case, hospitalisation, and death data were procured from a variety of sources described in section 3.

##### Section 3.1.2 Modelling sequence data

For the modelling of the GISAID data, we first apply inclusion / exclusion criteria by location. The primary reason to apply inclusion / exclusion rules is that the sequence data reported to GISAID is not guaranteed to representative. There are few locations globally that have only a small number of infections sequenced, and in some cases, these sequences were based on contact tracing of someone entering from a location with a specific variant of concern. As such, we apply 3 general rules for minimal data for modelling. We do not consider a location for sequence modelling unless: 1) they have one variant of concern with at least 200 sequences; 2) they have two variants of concern with at least 100 sequences each; or 3) they have one variant of concern with at least 50 sequences in the last six weeks. On a case-by-case basis we investigate each instance of a location almost passing a requirement (e.g., a location with 190 sequences of a single variant of concern will be researched to identify if there are other reasons to presume there is local transmission, in which case we would include it). We then only model those variants that satisfied one of the three requirements for that location, reclassifying all other variant sequences in that location as “other”.

Once the location/variant set is defined, we model each location’s included variants with a sequential logistic regression using the regression tool MR-BRT.^9^ In particular, by day, we model what fraction of all circulating infections are of each variant type. First, we identify the variant with the most sequences (which frequently is the “other” variant) and model what fraction of all variants are of that type by day. We then move to the next most common variant for a location, and model what fraction of the unaccounted-for variants each day are of that type. We continue in this manner until we have modelled 100% of each day’s variant distribution. Second, we use our fitted model to forecast into the future the fraction of each day’s variants that are of each type.

We model Delta and Kappa separately but given the ubiquity of the Delta variant, we combine their estimates. Finally, as there are many locations that began sequencing data well after their invasion, we anchor each regression by imputing an anchor for each variant at zero percent 3 weeks before the first included variant of concern data point.

##### Section 3.1.3 Model calibration

Our base model attempts to best capture the date and speed of a variant invasion according to the GISAID database. New variant invasions form a boundary condition in our SEI transmission model, and we found that the raw results from our initial model to be inconsistent with reported trends in cases, hospital admissions, and deaths. Part of the discrepancy is explained by differences in the reporting mechanisms. GISAID collates its own data sources with reporting mechanisms that are unclear to us, and so data may or may not be representative of the population of a country or subnational unit in question, the data may be lagged or aggregated from multiple data sources with different lags, and the data may be exposed to other issues that compromise the quality of the reports in ways we cannot accommodate. There are also just far, far fewer sequences of COVID-19 cases than there are cases (and often fewer sequences than hospitalisations or deaths). These data quality issues can lead the sequence model to predict variant invasions in the middle of downturns in transmission and invasions at periods where there is almost no reported COVID-19 transmission. The second issue is definitional: our model of GISAID sequences seeks to identify the date at which a new variant begins to compete with and dominate transmission in a particular country or subnational unit, assuming the sequences are representative of the location as a whole. Our SEI transmission model is looking for boundary conditions that mechanically explain sudden surges in transmission in a particular country or subnational unit, assuming current transmission is representative of the location as a whole. As a result, invasions according to sequence data can often occur weeks or months ahead of a surge in COVID-19 transmission.

To resolve the inconsistencies between the models in the service of making the best estimates and predictions for the geographies we model, we elected for a manual calibration procedure that uses unexplained surges in reported cases, hospital admissions, and deaths as the primary indicator of a new variant invasion and then used the sequencing model and GISAID data as an indicator of variant presence and timing. For this calibration, we built panel plots with the total daily sequences from GISAID, the sequence rate (daily sequences / daily cases), the variant proportion from our base sequence model and then the reported daily cases, hospital admissions, and deaths. Atop these plots, we drew vertical lines representing the base model predictions for the variant invasion date in purple and the post-calibration invasion dates in red. Invasion dates were then adjusted by hand to best align with transmission surges and variants were selected to best match our sequence model. For the two Omicron waves we model, we found the GISAID sequence data to generally be a poor match and we relied entirely on the manual calibration process for invasion date selection. Calibration plots are included for all locations in appendix 6, section 2. Our transmission model attributes reported cases, hospital admissions, and deaths to particular variants, and so we used the outputs of model runs in an iterative process to tweak variant invasion dates using area plots of the variant-split metrics (see appendix 6, section 3).

##### Section 3.1.4 New variant invasion scenario

New variants of SARS-CoV-2 are inevitable. The exact nature of the next variant of concern and its origin and timing are unknowable. To account for the uncertain nature of the next variant, we have constructed a set of parameterizations that blend the characteristics of both the Delta variant and the Omicron variant, as detailed in the main text. For the origin and timing of the next variant, we have decided to have the new variant first appear in South Africa on January 15, 2023, and then follow the same invasion pattern relative to this first invasion as the Omicron variant did. There are many options we could choose to use for origin and timing but using the Omicron invasion pattern lets us use a historically realistic pattern of location-to-location spread in what we expect is a representative behavioural and mandate policy environment.

#### Section 3.2 Vaccination rates

The rapid development and distribution COVID-19 vaccines has been one of the primary pharmaceutical interventions used to reduce SARS-CoV-2 transmission and mortality. Forecasting SARS-CoV-2 transmission therefore requires data on historical vaccine uptake as well as an accurate prediction of population level vaccination coverage in the future. Since COVID-19 vaccines were developed in response to the ongoing pandemic, quantifying vaccination coverage depends on a suite of factors which include the supply and manufacturing constraints of the vaccines, international purchasing agreements, which vaccine brands are available in each country, and the willingness to be vaccinated in the population (vaccine hesitancy).

To predict population vaccination coverage in the future, we constructed location-specific models that project the daily rate of vaccination as a function of vaccine supply, the current trajectory of vaccine administration (shots in arms), and vaccine hesitancy. More detailed descriptions of each component can be found in the following sections. For the overall mathematical framework of vaccine delivery, we define the proportion of the population that has received at least one dose in location $j$ and time $t$ as:

$p_{jt} =\frac{V_{jt}}{N_{j}}$.

Where $V_{jt}$ is the total number of individuals with a first vaccine dose and $N_{j}$ is the population size of location $j$. The value of $V_{jt}$ (past or future) is given by the cumulative sum of the daily rate of vaccination up to time $t$:

$V_{jt} = \sum_{t=1}^{t} r_{jt}^{'}$.

The effective daily rate of vaccination $r_{jt}^{'}$ is determined empirically from past observations, but future time steps are estimated using a flexible MR-BRT spline model fitted to observed values of $p_{jt}$ that asymptotes at the total vaccine-eligible population subject to available vaccine supply and population willingness to be vaccinated. This provides a constrained projection of vaccination rate $r_{jt}^{'}$ that is limited by both vaccine supply and vaccine demand:

$r_{jt}^{'} =\text{Min}\left\{ r_{jt}, a_{jt}, d_{jt} \right\}$.

The constraints on vaccination rate are defined by separate models and data for vaccine supply (described in Section 6.2.2) and vaccine demand (described in Section 6.2.3). The supply constraint is given as $a_{jt}$, which reflects the total number of doses available for administration in location at time $t$. Available doses are determined by vaccine supply that is dynamic over time with increases due to purchases and delivery of doses from manufacturers and decreases from administration to the population:

$a_{jt} = \sum_{t=1}^{t} s_{jt}-V_{j,t-1}$.

The daily rate of vaccination is also constrained by vaccine demand $d_{jt}$, which gives the total number of unvaccinated individuals that are willing to be vaccinated. Demand is estimated as:

$d_{jt}=N_{j}\left( 1-p_{jt} \right)w_{jt}$,

where the population size of location $j$is scaled by the proportion of the population that is unvaccinated $1-p_{jt}$ and the proportion of unvaccinated individuals $w_{jt}$ which is based on survey responses in the Facebook Global Symptom Survey reporting the number of unvaccinated individuals that would accept a vaccine (details below).

The above model provides estimates of initially vaccinated individuals (first dose). Quantities for subsequent vaccine doses (fully vaccinated and boosters) were estimated by lagging according to the corresponding dosing period and scaling by the dropout rate, which were both calculated empirically from observed data in each location. Models of vaccination coverage were applied to low and high-risk populations (under 65 and 65+ respectively) and stratified into brand-specific estimates based on the available proportions of each vaccine brand in location $j$and time $t$.

##### Section 3.2.1 Vaccine delivery

Data on vaccinations delivered were sourced from Our World in Data for most nations,^12^ apart from the United States, Mexico, Canada, Brazil, Germany, India, Italy, Spain, and China, where datasets tracking subnational vaccination numbers were used instead. Vaccination data for subnational locations was extracted from public government webpages or acquired through collaboration with national ministries of public health. Most sources provided time series of the number of people vaccinated (received any dose), fully vaccinated (one dose Janssen or two doses of other vaccines) and boosted (second dose Janssen or third dose of other vaccines). In cases where time series were incomplete, we assumed vaccination scale up followed a square root increase starting from the date of emergency use authorisation in that country. We found evidence that the US Centers for Disease Control (CDC) numbers lag behind state reports by 3 days, so we shift the CDC reported vaccinations by 3 days before adjusting our estimates. Data sources are listed in appendix 2, sections 12, 13, 14, and 15.

When data for fully vaccinated or boosted individuals was sparse or only a point estimate, we inferred the full time series by scaling the complete time series of fully vaccinated by the proportional relationship to the point estimate. When the data for fully vaccinated or boosted was sparse, we calculated the proportional relationship for each of the observed time points and then linearly interpolated the proportional relationship before scaling fully vaccinated numbers to account for changes in the administration of different vaccine courses over time. Thus, these data provide full time series for number of people initially vaccinated, fully vaccinated, and boosted for each location and time. The observed daily rate of vaccination described above ($r_{jt}$) therefore is the daily difference in reported cumulative timeseries ($V_{jt}-V_{j,t-1}$).

To estimate the trajectory of vaccination rate, we first find the ceiling for vaccination coverage in each location by estimating the percentage of the population that is either already vaccinated or willing to be vaccinated up to the current time point. Using MR-BRT,^9^ we fit a hierarchical [spline model](https://stash.ihme.washington.edu/projects/CVD19/repos/covid-beta-inputs/browse/src/covid_beta_inputs/vaccine_coverage/spline_fit_scaleup.R) in logit space with a linear offset and a monotonicity constraint where the independent variable is days since first vaccination. Specifically, we model the logit fraction of the population that has been vaccinated among the *total* willing population ($p_{jt}+d_{jt}/N_{j}$) that reported that they would definitely get vaccinated as a function of time since the first day of vaccination. The model was used to predict an average scale global curve for the predicted percentage of the population that is likely to be vaccinated. The resulting scale-up curve better approximates the observed slowing rate of vaccination as countries approach the maximum number of people who are willing to get vaccinated. The average scale-up curve was calibrated to the observed number of vaccinations reported as delivered in each location. This was done by calculating the ratio of the predicted cumulative percentage vaccinated over the observed percentage vaccinated for the most recent time period. This ratio was then used to adjust the average scale-up curve. For locations without observed data, we used the regional average ratio to calibrate the scale-up curve.

##### Section 3.2.2 Vaccine Supply

Data on vaccine purchasing unilateral and multilateral agreements are from the Duke Global Health Innovation Center (<https://launchandscalefaster.org/COVID-19>), Linksbridge, and relevant news articles^13^ . Supply data also include multilateral donation agreements that are part of the Gavi COVID-19 Vaccines Global Access Facility Advance Market Commitment (COVAX AMC; https://www.gavi.org/gavi-covax-amc). COVAX AMC provides financing and delivery mechanisms that facilitate donations of COVID-19 vaccines to 92 low- and middle-income countries. Together these data represent, by country or purchasing block, the number of doses secured and optioned from the primary approved and in development vaccines. Data on manufacturing capacity by quarter were from Linksbridge. Dates of initiation of vaccine delivery were from news reports when available and assumed to be the first date of daily vaccinations delivered based on Our World in Data when the date of emergency use authorization was not available. Starting in 2021, we have data on vaccinations delivered for most locations (100+) from [Our World in Data](https://ourworldindata.org/covid-vaccinations) and from the [United States CDC](https://covid.cdc.gov/covid-data-tracker/#vaccinations). To match the reported number of people vaccinated, we simply use reported numbers of people, when available. For missing dates, we linearly interpolate cumulative vaccinations. Together, these data sources provide the total number of doses supplied to a location up to timepoint, denoted as in the model above.

When constructing estimates of vaccine supply, we assume that the number of doses that can be manufactured per day is constant across quarters in 2021 (meaning that the doses per day is quarter capacity divided by days in the quarter). We also assume that when a vaccine first becomes available, there will be a stockpile that is equal to the previous quarter’s capacity. That stockpile is used over the first 30 days of the vaccine being available. There are two exceptions for the Pfizer/BioNTech and Moderna vaccines where we use the manufacturers’ stated capacity for the remainder of 2020. This leads to a large bump in available doses when a vaccine first becomes available. Otherwise, since we assume a constant number of doses manufactured per day, this is a step function over time. We then assign doses to locations based on purchasing and donation agreements and spread the number of secured doses across the stockpile doses and the daily manufacturing capacity for each manufacturer.

##### Section 3.2.3 Vaccine hesitancy

Willingness to accept the COVID-19 vaccine is based on data from the COVID-19 Trends and Impact Survey (CTIS). The CTIS is comprised of two data sources: national level data for over 190 countries collated by the University of Maryland (https://covidmap.umd.edu/) and state level data within the USA collated by the Delphi Group at Carnegie Mellon University (https://delphi.cmu.edu/covidcast/survey-results/)^14,15^ . The surveys are administered in collaboration with Facebook in 56 languages globally. To estimate vaccine willingness in the unvaccinated, we used survey question V1 in the global survey and V3 in the US survey, which ask “If a vaccine against COVID-19 were offered to you today, would you accept it?” to which respondents can select one of four answers: “Yes”, “Yes, probably”, “No, probably”, or “No”. Data from the surveys were tabulated by location-date. Note that this survey question is administered only to respondents who have previously answered that they are *not* vaccinated.

We estimate a smooth timeseries of vaccine willingness ($w_{jt}$) by fitting a Quasi-binomial Generalized Additive Model (GAM) with a thin plate smoothing term applied across dates.^16^ The spline model was fitted to tabulated data on the proportion of people, by location-date, who said they would accept a vaccine against COVID-19 if it were offered to them today. These data are weighted to be representative of the general population using weights derived by the survey-developers (more below). In locations where CTIS data is not available we used a regional model where the same GAM was fitted to a composite dataset comprised of several neighbouring countries in the region. However, for China we used a surrogate hesitancy model fitted to data from Singapore. When predicting future vaccination coverage, levels of vaccine hesitancy in the future were inferred by simply taking the average of the last three days of the smoothed time series to provide a single estimate of future vaccine acceptance by location. The value represents the fraction of adults on the model date that have would definitely receive the vaccine.

#### Section 3.3 Vaccine Efficacy and Waning Immunity

Vaccine efficacy refers to two aspects of a vaccine’s performance – first, its ability to prevent the transmission of COVID-19 from one person to another; and second, its ability to prevent an exposed person from developing severe symptoms leading to hospitalization and death. In our analysis, these features of a vaccine vary by the brand of vaccine delivered, by the number of doses an individual has received, by the COVID-19 variant an individual is exposed to, and by the time since the individual received their last course of vaccination. We assume booster doses of a particular brand of vaccine carry the same efficacy and waning pattern as the first full course of vaccination with the same brand, irrespective of time since the first course was completed or the brand used in the first course of vaccination.

##### Section 3.3.1 Maximum effect sizes

We have conducted a systematic review of literature on the effectiveness of all vaccines against infection and severe disease (interpreted as prevention of death and hospitalization). Our data sources include peer-reviewed publications, reports, preprints, medRxiv, and news reports, which are summarized in appendix 2, section 17. We track efficacy by the variants D614G (Ancestral), B.1.1.7 (Alpha), B.1.351 (Beta), P.1 (Gamma), B.1.617 (Delta), and BA.1 (Omicron). D614G is also referred to as “ancestral-type” COVID and was the dominant strain of SARS-CoV-2 infection before the Alpha variant spread widely. Due to limitations in available data, we have pooled the effect sizes into three groups – first, ancestral COVID and the Alpha variant; second, the Beta, Gamma, and Delta variants; and third, the Omicron variant. Maximum vaccine efficacy is summarized in table S2 which was retrieved from the IHME special analysis on vaccine efficacy on January 10, 2023, (<https://www.healthdata.org/covid/covid-19-vaccine-efficacy-summary)>.

##### Section 3.3.2 Waning immunity

To estimate waning protection from vaccination, we performed a systematic review and meta-analysis of scientific literature studies that estimated vaccine effectiveness by outcome, vaccine brand, and variant.^17^

##### Section 3.3.3 Reduction in transmission intensity

To generate inputs for our model, we constructed curves representing the average risk reduction among all people who last received a particular vaccine round. These risk reduction curves were constructed from the brand-specific vaccinations produced by the methods in Section 5.2 combined with the efficacy and waning curves derived from our systematic review and analysis. We first define the time-dependent effectiveness $\varepsilon_{x,v}^{Z}(t, b)$ of vaccine course $v$ of vaccine branch $b$ at preventing outcome Z against variant $x$ as

$$\varepsilon_{x,v}^{Z}\left( t, b \right)=\epsilon_{x,v}^{Z}\left( b \right)w^{Z}(t, b)$$

Where $\epsilon$ is the maximum effect size described in Section 5.3.1 and $w$is the relative waning curve described in Section 5.3.2. Then

$$\eta_{x, v}^{I}\left( t \right)=\frac{\int_{0}^{t} \sum_{b} \psi_{v}\left( t-\tau, b \right)\varepsilon_{x,v}^{I}\left( \tau, b \right)d\tau}{\int_{0}^{t} \sum_{b} \psi_{v}\left( t- \tau\right)d\tau}$$

is the average reduction in transmission intensity against variant $x$ among individuals whose last vaccine course (full first course, first booster, etc.) was $v$as a function of the current time $t.$ Similarly,

$$\eta_{x, v}^{Z}\left( t \right)=\frac{\int_{0}^{t} \sum_{b} \psi_{v}\left( t-\tau, b \right)(1- \frac{1- \varepsilon_{x,v}^{Z}\left( \tau, b \right)}{1-\varepsilon_{x,v}^{I}\left( \tau, b \right)})d\tau}{\int_{0}^{t} \sum_{b} \psi_{v}\left( t- \tau\right)d\tau}$$

is the average reduction in the infection-fatality ratio (when $Z=Death$) or the average reduction in infection-hospitalization ratio (when Z$=Hospitalization$) among individuals infected with variant $x$ whose last vaccine course was was $v$ as a function of the current time $t.$

##### Section 3.3.4 Waning infection-derived immunity

For waning of infection-derived immunity, we performed a systematic review and meta-analysis^18^ to establish the time pattern of waning protection from infection and severe disease due to prior infection $w^{Z}\left( t \right)$ where $Z$ corresponds to the outcome (infection, death, hospital admission).This waning pattern is then paired with the maximal cross-variant immunity, $\phi$, summarized in table S3 to form a pattern of waning protection specific to time since last exposure with variant $x$ when being challenged with variant y:

$$\varphi_{x,y}^{Z}\left( t \right)=\phi_{x,y}^{Z}w^{Z}(t)$$

#### Section 3.4 Pneumonia seasonality^1^

Pneumonia is one of the main clinical syndromes associated with respiratory SARS-CoV-2 infection and its seasonality is notable in many locations, particularly those far from the equator. This could be due to climatic variation (relative humidity, average air temperature) or due to human behaviour (greater time spent indoors). We modelled the ratio of pneumonia deaths in each week to the average weekly pneumonia deaths by location. As such, ratios above 1 indicate that more pneumonia deaths than the yearly average occur in that week, and ratios below 1 indicate that fewer deaths than the yearly average occur.

##### Section 3.4.1 Data sources

For locations where we have weekly vital registration data for pneumonia deaths, we used the data to directly model this ratio. Pneumonia deaths include all deaths classified by the full range of ICD codes in J12 - J18.9.

##### Section 3.4.2 Estimation process

To account for uncertainty in vital registration data and model type, all ratios were estimated 1000 times in a meta-regression model using MR-BRT.^9^ The meta-regression used a cubic spline on week and 1% trimming of the data inputs. The proportion of deaths in each week was calculated as the weekly number of deaths over the annual number of deaths in a location. The standard error was calculated using the formula for binomial variance:

$$variance=\sqrt{\frac{\frac{weekly death count}{annual death count}*(1 -\frac{weekly death count}{annual death count})}{annual death count}}$$

For locations without data on pneumonia deaths, the strategy included additional models and calculations to generate estimates for all locations. We modelled the global seasonality trend pooling all pneumonia deaths data, calculated the amplitude of the seasonality time series in specific locations to model and predict the relationship between amplitude and latitude, and then used the estimated amplitude values by latitude to manipulate the amplitude of the global pattern. As such all locations without data have the same general seasonality pattern (higher in October to April in Northern Hemisphere; higher in April to October in Southern Hemisphere), but the amplitude varies by location, depending on the latitude. In order to preserve the cyclical trend of the pneumonia deaths in the model, the same 52 weeks of data were triplicated, and added to the beginning and the end of the time series.

#### Section 3.5 Testing per capita^1^

Testing for COVID-19 can impact the epidemic both directly and indirectly. Directly, a positive test result alerts an individual to their need to self-isolate and for their contacts to quarantine. Indirectly, higher levels of testing ensure that policy makers and healthcare professionals have accurate information when making decisions about social distancing mandates and resource allocation.

##### Section 3.5.1 Data sources

Data on the number of tests administered were sourced from a combination of direct reports from government health authorities including the United States Department of Human and Health Services, and Our World in Data for all locations that were present in their database that we had not sourced from direct reports, supplemented by additional country resources when missing. Sources for these data are detailed in appendix 2, section 15.

##### Section 3.5.2 Descriptive model

When both daily and cumulative data were present on the same date for a given location, we gave preference to the cumulative data. When there were daily data reported in between gaps in cumulative data reports, we added the daily data to the preceding cumulative value to fill in the missing cumulative data. Dates where only positive tests were reported were dropped. Cumulative data preceded by days of no reports was shifted to the midpoint of the missing interval and scaled to equal the average daily tests over the interval. In locations where the date of the first confirmed case preceded the date of the first reported tests, we assumed that testing started two weeks before the first confirmed case and linearly interpolated to the first day of cumulative testing counts. We then aggregated to weekly intervals and linearly interpolated the weekly data with knots placed at the middle of each week. Finally, we smoothed the weekly interpolated data using ten iterations of smoothing with a uniform kernel and a three-day bandwidth. Finally, we assumed that daily tests were at least two times greater than reported cases (50% positivity rate), increasing daily testing counts in some locations early in the pandemic.

##### Section 3.5.3 Testing forecasting model

We projected levels of daily testing per capita using the location-specific mean daily difference in testing per capita for locations with data; in effect assuming that future growth in daily testing per capita will match past increases in testing. For locations that were missing testing data, we used the regional average growth of daily testing per capita and the date of the location-specific first reported case to impute both the past and the future values of daily testing per capita. Daily testing per capita increases in all locations until reaching a threshold value for the maximum daily tests per capita, a predicted level from a frontier analysis of testing rates by lag-distributed income per capita (LDI).

#### Section 3.6 Mask use

Mask use has proven to be an incredibly effective intervention strategy in reducing the transmission of COVID-19.^19,20^ While its importance appears to have waned in the presence of vaccines, the emergence of escape variants like B.1.617.2 which evade both natural and vaccine-based immune response indicates that mask use still has a vital role in the long-range trajectory of the pandemic. We analysed survey data on the levels and trends of mask use. This analysis of survey data sought to estimate the proportion of people who self-reported always wearing a facemask when outside their homes.

##### Section 3.6.1 Data sources

For mask use coverage levels, data was used from the The Delphi Group at Carnegie Mellon University and University of Maryland COVID-19 Trends and Impact Surveys (CTIS). The surveys are administered in collaboration with Facebook in more than 100 countries and in 56 languages globally. Mask use data are also from PREMISE Behavior survey, the Kaiser Family Foundation, and the YouGov COVID-19 Behaviour Tracker survey. The CTIS is comprised of two data sources: national level data for over 190 countries collated by the University of Maryland (<https://covidmap.umd.edu/>) and state level data within the USA collated by the Delphi Group at Carnegie Mellon University (<https://delphi.cmu.edu/covidcast/survey-results/>). There have been 13 waves of the CTIS and phrasing has changed slightly between waves, as of the current wave 13, the survey questions ask: “In the past 7 days, did you wear a mask most or all of the time in public?” (with the available answers being “Most of the time” and “All of the time”) and “In the past 7 days, when you were out in public, did most or all other people wear masks?”(with the available answers being “Most of the people” and “All of the people”). Respondents for “All of the time” and “All of the people” were the numerator in our proportion. For the PREMISE surveys, the following question was asked: “When you leave your home do you typically wear a face mask (SELECT_ONE)” with responses “Yes, always; Yes, sometimes; No never”. Respondents for “Yes, always” were the numerator in our proportion. From YouGov, the following question was asked: “Thinking about the last 7 days, have you worn a face mask outside your home (e.g. when on public transport, going to a supermarket, going to a main road)” with responses “Always”, “Frequently”, ”Sometimes”, ”Rarely”, and “Not at all”. Respondents for “Always” were the numerator in our proportion. Data sources are listed in appendix 2, section 16.

All surveys we used to estimate historical levels of mask usage ended well before the end of our reported case, hospitalisation, and death data (PREMISE on March 9, 2021, YouGov on April 6, 2021, and CTIS/Facebook on June 6, 2022). We use all available evidence and begin our mask use forecasts at the latest available date in each country or subnational unit, depending on survey coverage.

##### Section 3.6.2 Mask use forecasting model

We used a smoothing model to produce estimates of observed mask use. This smoothing process averages each data point with its neighbours. Locations with fewer than 30 daily respondents on average were smoothed to a greater degree than locations with more than 30 daily respondents.

We produced two scenarios of our forecasts. In our reference scenario we assume mask use remains constant into the future. This assumes that the world will continue to face waves of new very infectious but mild variants (mild relative to the severity of the original SARS-Cov-2 virus and early variants) and that population behaviours are at a steady state with respect to that level of risk. In our increased mask use scenario, we ramp up mask use to 80% linearly in the over the first week of the forecast. This scenario represents a policy response where masks are encouraged or mandated in order to prevent a large surge in COVID infections and related outcomes.

#### Section 3.7 Social distancing mandates

Some version of public health and social mandates have been implemented in nearly every country in the world. Some have presented as recommendations, others as requirements; some have presented as fragmented updates that escalate over a few days or weeks, others as discrete events where a nation transitions from no measures to full implementation of strict social distancing measures. We collected a global database containing social distancing mandates (also known as non-pharmaceutical interventions or NPIs) from January 1, 2022, to October 14, 2022, across 176 countries and 206 subnational units. We constructed 21 binary NPI variables which represent an individual government mandated intervention to reduce incidence of COVID-19, such as the closure of dining establishments or gathering restrictions of a specific size. The dates of initiation and cessation for each NPI were manually collected from academic organizations,^21,22,23^ business,^24^ non-governmental organizations,^25^ legal documentation hubs, and news outlets. Our mandate classification system emphasized two key aspects of social distancing mandates (i) policies that map to specific venues or populations i.e., closures of establishments for indoor dining, as opposed to general mandates such as “business restrictions” and (ii) policies that are binary in their hypothesized impact on the potential for transmission rather than a lessened effect i.e., we emphasize complete closures of non-essential retail, rather than partial capacity limits.

##### Section 3.7.1 Detailed mandate descriptions

In many circumstances, closures and reopenings were staggered, or partial for the entire sector we have outlined. This could be due to portions of the affected groups being included (e.g., school Grades 1 and 2 remained remote, but grades 3-6 were in-person) or due to geographic variations within the countries considered (e.g., specific counties within a state having local closures in the absence of a state-wide requirement). Whenever this was the case, given our desire to characterize the intent of the mandate, we utilized a fuzzy majority rule in determining whether the mandate was, or was not, still in place. This either was when most of the subunits were affected (e.g., four of six school grades constitute a majority), where most of geographic units (e.g., counties within a state) were affected, or it was clear that only a specific subset were outliers (e.g., bookstores were reopened, but all other retail remained closed). Where population tallies were made available, these were used to determine a majority.

The following mandates were tracked, with accompanying definitions below. They are collated by sector.

**Education**

Educational institutions have been subject to various mandates during the pandemic. Circumstances where schools served as essential services, such as in providing childcare support for essential workers, or in continuing lunch service for local communities were excluded.

*Primary education* – no in-person classroom activities for primary schools, grades 1-6.

*Secondary education* – no in-person classroom activities for secondary schools, grades 7-12.

*Higher education* – no in-person classroom activities for higher education institutions.

**Travel**

Entry restrictions included obligations to quarantine or abide by testing regulations prior to entry

*Borders closed to any* – entry restrictions imposed on any non-resident citizen group (mostly based upon nationality).

*Borders closed to all* – entry restrictions imposed on all non-resident citizens.

*Local travel restrictions* – citizens require specific permissions to travel beyond approximately 5km of their place of residence.

**Gathering Restrictions**

All gathering restrictions reference a specific threshold for which indoor and outdoor gatherings must not be greater than. The emphasis in these mandates is with respect to private gatherings. To be within a given tier, both of the two conditions must be satisfied. If a given tier of gathering restriction is in place, so too are all less stringent tiers. When a stay-at-home order is in place, so too are all tiers of gathering restriction. The following combinations were tracked, in order from most stringent, to least:

*Tier 1 gathering restrictions* – Indoor gathering limited to 6 persons, outdoor gatherings limited to 10 persons.
*Tier 2 gathering restrictions* – Indoor gathering limited to 10 persons, outdoor gatherings limited to 25 persons.
*Tier 3 gathering restrictions* – Indoor gathering limited to 25 persons, outdoor gatherings limited to 50 persons.
*Tier 4 gathering restrictions* – Indoor gathering limited to 50 persons, outdoor gatherings limited to 100 persons.
*Tier 5 gathering restrictions* – Indoor gathering limited to 100 persons, outdoor gatherings limited to 250 persons.

In addition, a non-thresholded “Any Gathering Restriction” was tracked.

**Stay-at-home orders**

*Stay-at-home order* – there is a mandate requiring all citizens to stay-at-home unless undertaking essential activities.

*Stay-at-home order with legal penalty* – there is an accompanying penalty (including financial penalties or imprisonment) for non-compliance with the stay-at-home order.

**Businesses**

“Non-essential” references local definitions rather than a universal global standard, due to the difficulties in cataloguing exhaustive lists. Most countries and territories retained a consistent core of services that were deemed “non-essential”. Exemplars of services that transitioned from non-essential to essential as the pandemic wore on are hardware stores and construction sites. These businesses included the following:

*Indoor dining closed* – catering establishments are closed to in-person indoor service.

*Indoor bars closed* – bars and other drinking establishments are closed to in-person indoor service.

*Gyms closed* – gyms and other personal fitness facilities are closed.

*Non-essential retail closed* – all non-essential retail is closed.

*Non-essential workplaces closed* – all non-essential workplaces (e.g., offices) are closed.

**Curfews**

Business curfews most often affected the restaurant and entertainment sectors as the emphasis was to minimize evening and night-time activities.

*Business curfews* – any business is prohibited from opening within their regular business hours

*Home curfews* – all citizens are unable to leave their homes unless for essential reasons within a certain time period.

##### Section 3.7.2 Mandate averaging and forecasting

To make the regression portion of our transmission model (Section 8.1) tractable, we collapse the 21 NPI variables into a single index of mandate intensity. We chose to do this with a simple averaging model. First, we computed the average of the sector-specific mandates by location and day to build a sector-specific index. Then we computed our mandate index as the average of the sector specific indices. This method of building a single mandate index is overly simplistic and is bound to over-estimate the effect sizes of some individual NPIs (such as gym closures) and under-estimate the effect sizes of others (such as stay at home orders), however disentangling the effect sizes of these individual mandates on transmission is problem riddled with definitional differences between locations and collinearity and is beyond the scope of our analysis.

We offer two forecast scenarios for mandates. The reference scenario, like our mask use reference scenario, assumes that the governmental risk tolerance is in a steady state in all locations relative to an ongoing level of infection consistent with highly transmissible and mildly severe (relative to Ancestral) COVID-19 variants, and thus is held constant into the future. In our mandate reimpositions scenario, we picked a location-specific daily death rate as a threshold for mandate reimpositions as well as a location-specific level at which to reimpose mandates.

For the threshold determination, we chose a candidate threshold for each location as five times the maximum reported daily death rate in the Omicron-era of transmission (the period in which our variant model, described in Section 5.1, says Omicron or one of its sub-lineages is the dominant variant). The candidate threshold was used if it was between 10 deaths per million and 30 deaths per million, otherwise the respective boundary was used for the location’s threshold. Our only exceptions to this algorithm were the provinces of China, where we manually set the threshold to 1 death per million, well above their historical lockdown thresholds during the zero-COVID era. The level of mandates reimposed in each location is the maximum level of mandates a location imposed in the post-Ancestral era in each location (the time span from when the first variant invaded said location and the end of our historical model for that location). Here, for Chinese provinces, we again make an exception and set the reimpositions level at its maximum to reflect the magnitude of the historical policy response. Diagnostics with location specific reimposition thresholds and reimposition levels can be found in Appendix 7.

Mandate reimpositions is done by running our transmission forecast model (Section 8) with our reference forecast of mandates, and then evaluating total daily deaths in each location. Any locations whose daily deaths exceed their threshold have mandates reimposed at their threshold level for 6 weeks and then turned off. Our forecast model is then run again on the subset of locations who reimposed mandates, their daily death rates are evaluated again, and then mandate levels are further adjusted among those locations who still exceed their mandate reimpositions threshold. This loop of transmission forecast and mandate reimpositions is repeated until all locations have forecast below their daily death rate thresholds for the entire forecast period, or mandates have been reimposed for the entire forecast period.

#### Section 3.8 Time-invariant drivers^1^

##### Section 3.8.1 Lower respiratory infection mortality

In the transmission model, the mortality rate due to lower respiratory infections (LRI) is captured as the location-specific age-standardized mortality death rate in the population 15 years or older. The 15+ years age-standardised LRI death rate is assumed to represent transmission of respiratory communicable diseases amongst adults.

Estimates of the LRI mortality rate come from the Global Burden of Diseases, Injuries and Risk Factors Study (GBD) and the methods for estimation are described in detail elsewhere^26–28^. Briefly, we used vital registration and verbal autopsy data in a Bayesian ensemble model which uses out of sample validity to produce a variety of plausible models which are weighted based on their performance in the final ensemble. Estimates are produced for each age, sex, year, and location. For this analysis, we used the age-standardized rate for both sexes by location in the year 2019 (most recent complete year of estimates).

##### Section 3.8.2 Altitude

The incidence and severity of lower respiratory infections, including pneumonia, is greater at higher elevation^29–31^. Altitude and humidity are believed to be a predictor of transmission and several studies have found greater mortality due to pneumonia at higher elevations, possibly due to decreased oxygen concentration at higher altitudes. The proportion of the population living below 100 meters above sea-level by location was obtained from GBD.^32^

##### Section 3.8.3 Smoking

The adult (15+ years) age-standardized tobacco smoking prevalence in 2019 was used as a covariate. This covariate is from GBD 2019 and estimation methods are described in detail elsewhere.^33^ Briefly, we estimated the prevalence of current smokers (daily or occasional) using individual-level and aggregated available survey data. The prevalence was modelled using space-time Gaussian Process Regression to produce smoothed estimates by space, time, age, and sex. For this analysis, we used age-standardized prevalence among both sexes. Smoking prevalence is location-specific.

##### Section 3.8.4 Ambient particulate matter pollution

Ambient particulate matter pollution is a covariate from GBD 2019^34^ and is defined as the population-weighted mean exposure to air particles with an aerodynamic diameter less than 2.5 micrometres per cubic meter of air. Input data for this model come from satellite observations, ground measurements, land use data, and chemical transport model simulations. Estimates are produced on a geospatial resolution and aggregated to the national level by population-weighting. This covariate is location-specific.

##### Section 3.8.5 Population density

Population density per pixel was calculated using Worldpop total population rasters and an area raster,^35^ and is represented as the percentage of the population living in areas denser than 2,500 people per square kilometre (km^2^) for a given country. By location, we determined the proportion of the population living in discrete categories of density and aggregated categories less than 2,500 per km^2^ for this analysis, using 2020 estimates to approximate population. The raster based Worldpop estimates were adjusted to match the country-totals from GBD.^36,37^

##### Section 3.8.6 Demography

Demographic data on national and subnational populations, namely the age structure of the population, is used to age-standardize several drivers and to split populations into high- and low-risk groups in the transmission model. Age distributions were obtained from GBD.^26^

##### Section 3.8.7 Lag-distributed income

Lag-distributed income is a moving average transformation of Gross Domestic Product per capita that uses a normalized ten-year lagged average. It was produced as a covariate for GBD 2019^37^ and used here to predict maximum testing per capita.

##### Section 3.8.8 UHC effective coverage

Estimates from the GBD 2019 were used to create an index which represents global progress towards universal health coverage (UHC) and specifically UHC effective coverage. The UHC effective coverage index is comprised of 23 indicators drawn across a range of health service areas and is meant to represent healthcare needs over the life course. It is one of the covariates in the production of location-specific infection fatality ratios for COVID-19.

##### Section 3.8.9 Healthcare Access and Quality index

Estimates of 32 causes of death that are amenable to healthcare from the GBD 2019^27^ were used to derive the Healthcare Access and Quality (HAQ) index, a summary measure of personal healthcare in each location. It is one of the covariates in the production of location-specific infection fatality ratios for COVID-19.

##### Section 3.8.10 Obesity

The age-standardised obesity prevalence covariate was estimated for each location as part of GBD 2019.^34^ It is one of the covariates in the production of location-specific infection fatality ratios for COVID-19.

##### Section 3.8.11 Diabetes

The age-standardised diabetes prevalence covariate was estimated for each location as part of GBD 2019.^27^ It is one of the covariates in the production of location-specific infection fatality ratios for COVID-19.

##### Section 3.8.12 Cancer

The age-standardised cancer prevalence covariate was estimated for each location as part of GBD 2019.^27^ It is one of the covariates in the production of location-specific infection fatality ratios for COVID-19.

##### Section 3.8.13 Cardiovascular disease

The age-standardised cardiovascular disease prevalence covariate was estimated for each location as part of GBD 2019.^27^ It is one of the covariates in the production of location-specific infection fatality ratios for COVID-19.

##### Section 3.8.14 Chronic obstructive pulmonary disease

The age-standardised chronic obstructive pulmonary disease prevalence covariate was estimated for each location as part of GBD 2019.^27^ It is one of the covariates in the production of location-specific infection fatality ratios for COVID-19.

##### Section 3.8.15 Chronic kidney disease

The age-standardised chronic kidney disease prevalence covariate was estimated for each location as part of GBD 2019.^27^ It is one of the covariates in the production of location-specific infection fatality ratios for COVID-19.

### Section 4 The SEI transmission model

The core component of both our past infections model and our forecasting model is a system of integro-differential equations. This system tracks groups of individuals by what variant (if any) of COVID-19 they were most recently exposed to, what round of vaccination (if any) they most recently completed, and whether they belong to a high risk (defined as over the age of 65) or low risk (all others) demographic group. The system also captures the waning of vaccine-derived and infection-derived protection from infection and severe disease. Figure S1 contains a compartment and transition diagram of the model structure.

The model uses three kinds of compartments to track individuals. $S$ compartments represent individuals with full or partial susceptibility to a COVID infection. Individuals in an $E$ compartment have been exposed to the virus but are currently asymptomatic and non-infectious. Finally, $I$ compartments contain those individuals with a current active case of COVID who are transmitting the virus to other people. Compartments are indexed by variant, vaccine status, and age group (e.g. $S_{x,v,a}$) . Suppressed indices imply summation (e.g. $S_{x,a}= \sum_{v} S_{x,v,a}$).

Variants are members of the set $X=\{\emptyset, A, \alpha, \beta, \gamma, \delta, o, ba5, \omega\},$ where $\emptyset$ is no variant, $A$ is the original ancestral COVID, Greek letters represent the WHO variants of concern, and $ba5$ is the Omicron subvariant we associate with the second large global wave in Omicron transmission. In the following description, we use $x$ and y as indices representing a variant. The variant index is contextual to the compartment. For susceptible compartments $S_{x,v,a}$ the variant $x$ represents the last variant an individual in the compartment was exposed to. In an exposed or infectious compartment, it instead represents the variant they are currently exposed to.

Vaccine statuses are members of the set $V=\{0, 1, 2, 3, 4\}$ and represent the number of courses of vaccination a person has completed, with $0$ representing no vaccination, $1$ representing a first full course of vaccination (with one or two doses depending on the vaccine), and $2, 3,$ and $4$ representing additional booster doses.. In the general model description, we use $v$ and $w$ as indices representing vaccine status. Finally, age groups are members of the set $A=\{under 65, over 65\}$ and are represented by an $a$ index. The age group index is suppressed in Figure S1, but there is a parallel group of compartments for each age group and no assumption of differential mixing between the groups.

We can then write the transmission model as

$$\frac{dS_{x,v,a}}{dt}= -\sum_{y} \lambda_{y}\hat{S}_{x,y,v,a}$$

$$+\frac{S_{x,v-1,a}\psi_{v,a}}{S_{v-1,a}}- \frac{S_{x,v,a}\psi_{v+1,a}}{S_{v,a}}$$

$$+\gamma I_{x,v}$$

$$\frac{dE_{x,v,a}}{dt}= -\sigma_{x}E_{x,v,a}$$

$$+\lambda_{x} \sum_{y} \hat{S}_{x,y,v,a}$$

$$\frac{dI_{x,v,a}}{dt}= -\gamma I_{x,v,a}$$

$$+\sigma_{x}E_{x,v,a}$$

All variables in these equations except $N$, $\sigma_{x}$, $\gamma$, and $\alpha$ depend on time, though we have suppressed that dependence in the notation. $N$ is the total population size of a location and is assumed to be invariant over the duration of the model. The other parameters are defined below, and their distributions are documented in table S3.

The first equation describes the change in the number of people susceptible based on their prior variant and vaccine exposure and can be broken down into three terms. The first term is new infections from the particular susceptible compartment due to each variant $y$ and has several key factors. We define

$$\hat{S}_{x,y,v,a}=\left( 1- \eta_{y,v,a}^{I} \right)\left( 1-\chi_{x,y,a}^{I} \right)S_{x,v,a}$$

as the count of people ­*effectively susceptible* to variant $y$ accounting for prior infection exposure to variant $x$ and vaccine exposure $v$ in age group $a$. As described in Section 5.3.3, the factor $\eta_{x,v,a}^{I}$ describes the average proportion reduction in transmission intensity against variant $x$ for people who have completed vaccine course $v$, accounting for time since vaccination and waning efficacy. The parallel factor for average infection-derived protection is $\chi_{x,y,a}^{I}$ which represents the proportion reduction in transmission intensity against variant $y$ due to prior exposure with variant $x$, accounting the time since last exposure. This term is calculated as

$$\chi_{x,y,a}^{I}\left( t \right)=\frac{\int_{0}^{t^{*}} E_{x,a}^{+}\left( t- \tau\right)\varphi_{x,y}^{I}\left( \tau\right)d\tau}{\int_{0}^{t^{*}} E_{x, a}^{+}\left( t- \tau\right)d\tau}$$

where $E_{x,a}^{+}(t)$ are new infections of variant $x$ in age group $a$, $\varphi_{x,y}^{I}\left( t \right)$ is the waning infection-derived protection of an individual with prior exposure to variant $x$ being challenged with variant $y$ as a function of time since last exposure, and

$$t^{*}=max_{t^{'}} \left\{ \int_{0}^{t^{'}} E_{x,a}^{+}\left( t- \tau\right)d\tau<S_{x, a}(t) \right\}$$

and represents an assumption that individuals who were infected the furthest in the past are the ones who will be next infected within each susceptible compartment. This assumption and the assumption that $\chi$ is independent of vaccine round were made to make the model computationally tractable. We also define

$$\lambda_{y}=\frac{\kappa_{y}^{I}\beta}{N}I_{y}^{\alpha}$$

which can then be interpreted as the force of infection due to variant $x$ among the effectively susceptible. The most important term in this equation for our model is the time-varying transmission intensity $\beta$. It is constructed to be independent of prior COVID exposure, vaccine status, age, and the variant an individual is being challenged with so that we can link it to other model drivers like mandate levels, mask usage, and seasonality. The relationships with vaccination and prior COVID exposure are captured in the definition of effectively susceptible. Then, $\kappa_{y}^{I}$ represents the increase in transmission intensity of variant $y$relative to Ancestral COVID and is time-invariant, and $I_{y}^{\alpha}$ describes count of people currently infectious and spreading variant $y$. This last term scaled down by a factor $\alpha$ which is used to account for imperfect mixing of population groups.

The second term in the first equation for $\frac{dS}{dt}$ represents entrances and exits from the susceptible compartment due to changes in vaccine status. Vaccine delivery is age specific but assumed to be independent of prior COVID exposure. This means that the count of new entrances into $S_{x,v,a}$ due to vaccination the total number of vaccines of round $v$ delivered to age group $a$, $\psi_{v,a}$, multiplied with the proportion of the vaccine eligible in with matching COVID exposure $\frac{S_{x,v-1,a}}{S_{v-1, a}}$. New exits due to vaccination with the next vaccine course $v+1$ follow the same logic. Here we use the arithmetic on the vaccine index to move positions within our ordered set of vaccinations $V$ and define any quantity with an index outside the set as 0. Our model assumes that vaccination and infection do not co-occur on a time step and that no one with an active infection (people in $E$ or $I$ compartments). The final term is the re-entry of infected people into the susceptible population ${\gamma I}_{x,a}$ where $\gamma$ is the reciprocal of the average duration of infectiousness.

The equation for $\frac{dE}{dt}$ describes the change in the count of people exposed to (but not spreading) variant $x$ with prior vaccination status $v$ in age group $a$. People exit the compartment at a rate $\sigma_{x}$ which is higher for the Omicron variant than other variants. The second term of this equation is parallel to the transmission term in the equation for the change in the susceptible population, except summed over all pools of effectively susceptible to the exposure variant.

The last equation in transmission model describes the change in the count of people infectious and spreading variant $x$ with prior vaccination status $v$ in age group $a$. People enter from the corresponding exposed compartment and exit at a rate $\gamma$ to the appropriate susceptible compartment when their infectiousness period ends.

The transmission dynamic equations are coupled with a set of lagged equations for the cases, deaths, and hospital admissions. Formally, we define the set of outcomes $O=\{D, C, H\}$ where $D$ is deaths, $C$ is cases, and $H$ is hospital admissions. For $Z\in O$, we can then say

$$\frac{dZ_{x,v,a}}{dt}=\lambda_{x}\hat{S}_{x,y,v,a}\theta_{x,y,v,a}^{Z}$$

where $\theta^{Z}$ is either the infection-fatality ratio, the infection-detection ratio, or the infection-hospitalization ratio for $Z$ equal to $D$, $C$, or $H$, respectively. The ratios and outcomes are indexed to the date of infection with outcome-specific lags drawn from uncertainty distributions. The force of infection and effectively susceptible have been defined above. The ratios are defined as

$$\theta_{x,y,v,a}^{Z}= \kappa_{y}^{Z}\left( 1- \eta_{y,v,a}^{Z} \right)\left( 1- \chi_{x,y,a}^{Z} \right)\zeta_{a}^{Z}\theta_{\emptyset,A,u}^{Z}$$

In this definition $\theta_{\emptyset,A, u}^{Z}$ is the all-age infection-outcome ratio among individuals with no infection- or vaccine-derived immunity experiencing an Ancestral-type infection. This ratio is initially estimated with the statistical model described in section 4, and has final estimates produced using the second parametrization of this transmission model. $\zeta_{a}^{Z}$ is an age-adjustment factor for the infection-outcome ratio that is produced as a part of the statistical model and is time-invariant. The $\eta^{Z}$ and $\chi^{Z}$ terms mirror their transmission reduction counterparts. $\eta^{Z}$ is defined in section 5.3.3 an $\chi^{Z}$ is defined as

$$\chi_{x,y,a}^{Z}\left( t \right)=\frac{\int_{0}^{t^{*}} E_{x,a}^{+}\left( t- \tau\right)\frac{(1-\varphi_{x,y}^{Z}\left( \tau\right))}{(1-\varphi_{x,y}^{I}\left( \tau\right))} d\tau}{\int_{0}^{t^{*}} E_{x, a}^{+}\left( t- \tau\right)d\tau}$$

where $\varphi_{x,y}^{Z}(t)$ is the waning protection offered against outcome $Z$ by variant $x$ among individuals with a variant $y$ infection at some time $t$ after the original infection with variant $x$, defined in section 5.3.4. Note that when $Z$ represents cases, both $\eta$ and $\chi$ are zero, meaning we assume prior infection or vaccination offers no differential reduction in cases beyond the reduction in transmission.

We use three parameterizations of the model in our estimation process. The first parameterization uses a paired input measure and epidemiological rate (deaths and the infection-fatality ratio, for instance) to estimate infections and transmission intensity. The second uses all input measures (cases, deaths, and admissions) available in a location and an estimate of transmission intensity to produce internally consistent final estimates of the epidemiological ratios. The final parameterization takes transmission intensity and the ratios as inputs and produces estimates of infections, deaths, admissions, and cases. The three parameterizations will be explained in detail in the context of their usage in following sections.

For our initial condition, we find the first day in each location with more than 50 infections ($t_{0}$) using the naïve ratio estimate from the statistical model and the corresponding measure data. Then we set

$$S_{\emptyset,u,a}\left( t_{0} \right)=N_{a}- E_{a}^{+}\left( t_{0} \right)-\left( \frac{E_{a}^{+}(t_{0})}{\beta_{0}} \right)^{1/\alpha}$$

$$E_{A,u,a}\left( t_{0} \right)=E_{a}^{+}(t_{0})$$

$$I_{A,u,a}\left( t_{0} \right)=\left( \frac{E_{a}^{+}\left( t_{0} \right)}{\beta_{0}} \right)^{1/\alpha}$$

where $E_{a}^{+}\left( t_{0} \right)=\frac{N_{a}}{N}E^{+}\left( t_{0} \right)$ is the total number of infections on the first day of the pandemic in age group $a$ and $\beta_{0}$ is an initial estimate of transmission intensity (assumed to be 2.0 in the first model parameterization and taken from prior model results in the second). All other SEI compartments are set to 0. The model also has a boundary condition that requires intervention when new variants invade a particular location. The invasion date $\tau$ for a variant is determined by our model described in section 5.1. At this point we adjust the system state so that $\delta=min(max \left( \pi E, 1 \right), 0.005*S)$ people move from the susceptible compartment to exposed with the new variant. Additionally, we send $\left( \frac{\delta}{2} \right)^{1/\alpha}$ people from the appropriate susceptible to infectious with the new variant. This mirrors the way we initialize the system. Here $\pi$ is a proportion sampled from $U[0.01, 0.10]$ and used to encode our observation that the speed of escape variant invasion is positively correlated with the size of the outbreak in a location where it invades.

To solve our system of integro-differential equations, we used a custom implementation of the classic 4^th^ order Runge-Kutta solver.^38^ The standard implementation is expanded both to allow boundary conditions beyond the initial condition to handle the emergence of new variants and to allow the periodic computation of the integral terms on a coarser step size than the non-integral terms. We used a step size of 0.1 days and recomputed the infection-derived immunity integrals $\chi^{I}$ and $\chi^{Z}$ every tenth time step. The model was written using Python 3.7 and optimized with NumPy^39^ and Numba.^40^ All modelling code is available online.^41^ Comprehensive diagnostics for our model and all future scenarios can be found in Appendix 4.

### Section 5 Estimating infections in the past

The past infection estimation process produces robust estimates of SARS-CoV-2 infections through the full history of the pandemic by location. It also produces posterior estimates of past infection-detection, infection-hospitalization, and infection-detection ratios that link the modelled infections to reported cases, deaths, and hospital admissions. We begin by using each of cases, deaths, and hospital admissions to make independent measure-specific estimates of transmission intensity using a statistical model of the historical ratios and the first parameterization of the SEI model. These estimates go through a calibration and model selection process that removes invalid or poorly behaved models from the results. The resulting transmission intensity estimates from each measure-specific model are averaged to produce a single estimate of transmission intensity that incorporates data from all measures. This averaged estimate is run through the second parameterization of the SEI model along with all available cases, deaths, and hospital admissions data to produce a full time series of infections, cases, deaths, hospital admissions by variant and vaccine status and the corresponding posterior estimates of the IDR, IFR, and IHR.

#### Section 5.1 Measure-specific estimates of $\boldsymbol{\beta}$

We make an initial estimate of transmission intensity $\beta_{fit}^{Z}(t)$ for each of cases, deaths, and hospital admissions for each location. The first step of the estimation process is to use the statistical model of historical ratios described in section 5 to produce a first pass estimate of the matched epidemiological ratio among the vaccine- and infection-naïve population experiencing an ancestral-type SARS-CoV-2 infection $\theta_{\emptyset,A,u}^{Z}$. This initial estimate of the ratio is done using a seroprevalence data subset to the period before vaccination or variant invasion where we can safely assume all infections correspond to our target denominator. The first parameterizations of the SEI model then uses this initial ratio estimate, $\theta_{\emptyset,A,u}^{Z}$, and the measure data used to generate it, $Z$, to generate a first pass estimate of variant and vaccine-status specific results. To do this, we can write the increment in the measures as

$$\frac{dZ}{dt}= \sum_{x,y,v,a} \frac{dZ_{x,y,v,a}}{dt}$$

$$= \sum_{x,y,v,a} \theta_{x,y,v,a}\lambda_{y}S_{x,y,v,a}$$

$$=\frac{\beta^{Z}}{N} \sum_{x,y,v,a} \theta_{x,y,v,a}\kappa_{y}^{I}I_{y}^{\alpha}S_{x,y,v,a}$$

Here $\frac{dZ}{dt}$ corresponds directly to an (appropriately scaled) version of our input data. On the right-hand side of the expression, everything inside the summation can be computed as a function of the input parameters, initial ratio estimation, and transmission history up to the current point in time. We can therefore rewrite this expression as

$$\beta^{Z}=\frac{N\frac{dZ}{dt}}{\sum_{x,y,v,a} \theta_{x,y,v,a}\kappa_{y}^{I}I_{y}^{\alpha}S_{x,y,v,a}}$$

to solve for the current transmission intensity. This scheme allows us to start with an initial condition and then calibrate transmission intensity to the measure data each time step.

The important outputs from this first run of the system are the subset of infections and the input measure $Z$ that are among the target denominator of individuals experiencing an ancestral infection without a vaccination or prior infection. This initial estimate is run through the historical ratio estimation model a second time, this time using all seroprevalence data to produce a more robust estimate of the target epidemiological ratio. The second-pass ratio and the measure are sent through the first parameterization of the SEI system a second time to produce final estimates of measure-specific transmission intensity $\beta_{fit}^{Z}\left( t \right)$ by location.

#### Section 5.2 Model ensemble and estimation of variant-specific outcome risk

To capture the uncertainty in the modelling assumptions, data, and parameterizations, we use an ensemble approach that generates 100 iterations (or *draws*) of $\beta_{fit}^{Z}(t)$ for each of cases, deaths, and hospital admissions. These iterations are created by varying the input data, model composition, and model parameterization. In addition to incorporating the uncertainty in assay sensitivity, total COVID mortality scalars, durations between exposure and reported outcomes, and ratio covariates as described in Barber, et al,^2^ we also incorporate uncertainty in transmission dynamics parameters, cross-variant immunity, variant-specific transmission intensity, and variant-specific risk ratios for hospitalization and death. The sampling distributions of these parameters are summarized in table S3.

The effects of many of these parameters are poorly specified in isolation and are often in conflict in particular locations. In some situations, the estimates of transmission intensity over time, $\beta_{fit}^{Z}\left( t \right),$ produced by these methods revealed that the case, hospitalization, and/or death data available for certain geographies were not compatible with the global risk ratio parameterization, $\kappa_{x}^{Z}$, described above – characterized by having large unexplained decreases or increases in estimates of $\beta_{fit}^{Z}\left( t \right)$ over a period associated with the invasion of a new variant. In these instances, we approximate the relative deviation in $\beta_{fit}^{Z}\left( t \right)$ and use that value to scale $\kappa_{x}^{Z}(l)$ for measure $Z$, location $l$, variant $x$.

In other situations, the conflict was so extreme that it caused the model to infect more people than exist in a location in a time period incompatible with any evidence on waning immunity. In these latter situations, we found the unrealistic estimates were a result of edge case samples in the joint distribution of the model parameters. For example, a single draw for a location may select a large total-COVID scalar and both a small death risk ratio $\kappa_{x}^{D}$ for Delta and Omicron, turning each reported death into tens or hundreds of thousands of infections. As we generally assume parameters to be independently distributed unless we have compelling evidence otherwise, this outcome is not totally unexpected.

To address these issues, we developed an iterative calibration approach with an algorithm designed to estimate an empirical risk profile for each variant that was consistent with the epidemiological surveillance data on which our model relies, and also allowed us to calibrate those risk parameters to each location.

##### Section 5.2.1 Global Calibration

For each non-ancestral variant of COVID-19, we estimated the risk of hospitalization (IHR) and death (IFR) relative to the ancestral variant for use in the transmission model (Section 6). We assessed the Alpha, Beta, Gamma, and Delta variants in the order in which they invaded. To evaluate a given variant, we first estimated infections using our measure-specific model of past transmission (section 7.1), running each location through to the date a subsequent variant invaded. Estimates of infections based on hospital admissions and deaths excluded seroprevalence studies that took place after the invasion of the variant being evaluated and were specified such that the IHR and IFR for that variant was the same as ancestral. Estimates of infections based on reported cases used all available seroprevalence data through the variant wave we were parameterizing, thus leveraging the relationship between more contemporary seroprevalence data and testing rates in the IDR time series (which we did not observe to be otherwise influenced by the invasion of more severe variants). For each location and draw, we then used the formula

$$r_{m}=\frac{I_{m,v,o}}{I_{m,v,e} \left( \frac{I_{m,p,o}}{I_{m,p,e}} \right)}$$

to define a risk ratio, $r$, for each measure, $m$, as the ratio of infections, $I$, due to a given variant,$v$, that are observed in the equal-risk model, o, to the infections due to that variant expected by the case-based model, $e$, after controlling for the relative difference in the observed and expected models during the period before the most recent variant invasion, $p$. We then examined the distribution of these values across the full set of location-draws, as well as just the draws in the subset of locations with seroprevalence data, to define plausible ranges from which to sample the IHR and IFR risk ratios for that variant (table S3). After parameterizing a given variant in the sequence, we moved to the next.

Due to the declining reliability of reported cases during the Omicron and BA.5 era, we were not able to simply use cases to derive expected infections for these variants, and in fact needed to parameterize the change in IDR in addition to IHR and IFR. In chronological order of invasion, we again ran our model of past transmission through the period in which a variant was dominant, excluding the relevant seroprevalence data and using ancestral-level risk levels for all three measure-specific models. These were used as observed infections in the risk ratio formula. We then extracted only the time period before the invasion of the variant in question to use in our regression model (section 8.1) and to subsequently build a counterfactual forecast (section 8.2 – 8.4) of infections for the variant based on each of the case-, hospitalization-, and death-based past time series. We used these counterfactual infections as expected infections in the risk ratio formula and determined the sampling range for the risk ratios in the same manner.

##### Section 5.2.2 Local Calibration

The degradation of reporting standards and inconsistent definitions of data during the Omicron and BA.5 era, as well as the reduced frequency of severe outcomes due to these milder variants, led to contextual variation in the epidemiological surveillance data that was exogenous to our model system, and time series that often resembled a stochastic process. Additionally, these variants are highly transmissible and were not met with much opposition in the way of non-pharmaceutical interventions, meaning most locations experienced surges that are difficult to parameterize – underestimates of past infections (and thus existing natural immunity) turn into implausible resurgent forecasts, while overestimates can inappropriately deplete the susceptible population. Addressing this required that we calibrate the case, hospital admission, and death data was incorporated into our model for each location (first for Omicron, then for BA.5).

We first modeled transmission through the time period in which variant we were calibrating was dominant, using the global risk parameters derived in the previous step as well as seroprevalence data from that time period, to be used as observed infections in the measure-specific risk calculation. The same counterfactual estimate of infections that was produced in the global risk assessment step was used as expected infections again here. We used the average infections across draws for each location in the formula, thus producing one scalar quantity in each location to shift the entire distribution of risk sampled at the global level. In a subset of locations, we were unable to calculate a robust risk ratio using this framework – typically, this was either a location without any seroprevalence data and was incompatible with the out-of-sample predictions from the baseline ancestral ratio model, or one in which the use of global parameters and local data in the observed model was too unstable to yield a baseline estimate of infections. In these instances, we approximate the relative deviation between observed and expected for use in local calibration for any offending variant.

#### Section 5.3 Model averaging and final estimates

To produce our final variant- and vaccine-specific estimates of cases, deaths, hospital admissions, infections, we need to build a composite estimate of transmission intensity $\beta_{fit}$. We do this by first decomposing each measure-specific estimate of transmission intensity, $\beta^{Z}$, into two values by day – total infections, $E^{+,Z}$, and a second quantity $Y^{Z}=\left( \frac{E^{+,Z}}{\beta^{Z}} \right)^{\frac{1}{\alpha}}$. We combine the measure-specific values using a simple average from the start of the time series up until 30 days from the end. In order to mitigate the effect of compositional bias due to different lags from exposure to each measure, as well as any variation in the recency of reporting for each measure, we also fit a spline regression model using a custom modelling framework called RegMod, ^42^ to the daily difference in the natural log of either value $E^{+,Z}$ or $Y^{Z}$ over time using a cubic spline with knots every five days and a constraint that the slope in the terminal interval be linear. We use the predictions from this model to stream out the last 30 days starting from the end of the averaged portion of the time series, resulting in composite estimates $E^{+}$and $Y$. From these we recover a composite model of $\beta_{fit}=\frac{E^{+}}{Y^{\alpha}}$ that averages the influence of the case, death, and hospital admission data in each location.

We then use the composite estimate of transmission intensity along with all case, death, and hospital admission data in each location to produce final estimates of variant and vaccine-status specific estimates of cases, deaths, admissions, and infections using the second parameterization of the transmission system. In this parameterization, the transmission dynamics of the SEI model are completely determined by the initial condition and the composite transmission intensity. We next write the equation for each measure

$$\frac{dZ}{dt}= \sum_{x,y,v,a} \frac{dZ_{x,y,v,a}}{dt}$$

$$= \sum_{x,y,v,a} \theta_{x,y,v,a}\lambda_{y}S_{x,y,v,a}$$

$$= \theta_{\emptyset,A,u}^{Z}\sum_{x,y,v,a} \kappa_{y}^{Z}\left( 1- \eta_{y,v,a}^{Z} \right)\left( 1- \chi_{x,y,a}^{Z} \right)\zeta_{a}^{Z}\lambda_{y}S_{x,y,v,a}$$

Then, as with the first parameterization, we are in a situation where $\frac{dZ}{dt}$ on the left-hand side is a known input value and everything inside the summation is a function of model parameters or transmission dynamics on previous time steps. We can thus rewrite the expression to solve for the epidemiological rate

$$\theta_{\emptyset,A,u}^{Z}=\frac{\frac{dZ}{dt}}{\sum_{x,y,v,a} \kappa_{y}^{Z}\left( 1- \eta_{y,v,a}^{Z} \right)\left( 1- \chi_{x,y,a}^{Z} \right)\zeta_{a}^{Z}\lambda_{y}S_{x,y,v,a}}$$

for each of the input measures $Z$. One way to interpret this parameterization is that we are absorbing all the discrepancies between the case-, death-, and hospital admission-specific models of transmission into our estimation of the epidemiological ratios. The full history of vaccine status and variant specific transmission that leverages all available reported data in each location.

### Section 6 Forecasting infections, deaths, admissions, and cases

In order to forecast the future of the pandemic, we first need to forecast the transmission intensity. For each of the 100 model draws we regress the historical transmission intensity fit to cases, deaths, and hospital admissions in the previous step, $\beta_{fit}\left( l,t \right)$ onto several of the model drivers described in section 6. The regression is done with linear mixed-effects model with constraints and Gaussian priors and produces a model of transmission intensity $\hat{\beta}(l,t)$. This modelled transmission intensity allows us to forecast transmission intensity as a function of the forecasts of our model drivers. We next produce our final model of transmission intensity $\beta(l, t)$ by aligning our regression model $\hat{\beta}(l, t)$ with our fit to past infections $\beta_{fit}\left( l, t \right)$ on the last day of infection data from our past infections model. We then adjust the long term trajectory of $\beta(l, t)$ to account for unexplained variation in recent trends transmission intensity using the regression residuals. Finally, we feed the forecast transmission intensity back into the third parameterization of our SEI model with projections of vaccine delivery and variant spread to produce forecasts of the COVID-19 epidemic over the next six months.

#### Section 6.1 Regression of transmission intensity onto model covariates

The next modelling step is to regress our fit betas against key model drivers by minimizing

$$f\left( \boldsymbol{\alpha} \right)\mathbf{=}\frac{1}{2}\sum_{l,t} \left( \ln\beta_{fit}\left( l, t \right)-\ln\hat{\beta}\left( l, t;\boldsymbol{\alpha} \right) \right)^{2}+\frac{1}{2}\sum_{l, c} \left( \frac{\alpha_{l,c}-\mu_{c}}{\sigma_{c}} \right)^{2}$$

subject to constraints of the form

$${lower}_{c}<\alpha_{l,c}<{upper}_{c}$$

Here $\boldsymbol{\alpha}$ is a vector of coefficients of the model covariates. The first term of the optimization is a standard sum of squared errors with

$$\ln\hat{\beta}\left( l, t;\boldsymbol{\alpha} \right)=\alpha_{intercept,l}+\sum_{c\in I} \alpha_{c}x_{c}\left( l \right)+\sum_{c\in V} \alpha_{c}x_{c}(l, t)$$

That is, our model of $\beta_{fit}(l, t)$ is a location-specific random intercept, a set of location-specific and time-invariant covariates $I$ with fixed effects for the coefficients, and a set of location-specific and time varying covariates $V$ with fixed effects for the coefficients. The second term of the objective function is a set of Gaussian priors on the coefficients. These priors vary by covariate but are not location specific. Regression constraints, priors, and effect sizes can be found in table S4. Given the nature of the constraints for the problem, we solve our regression using the reference implementation of the L-BFGS-B algorithm^43^ wrapped in the Python library SciPy.^44^

#### Section 6.2 Forecasting transmission intensity

For our final model of transmission intensity, we want to use our best fit to the infection data $\beta_{fit}(t)$ in the past and then leverage our model of transmission intensity as a function of the model drivers $\hat{\beta}(t)$ to tell us how transmission will proceed in the future. To do so, we need to address two issues – aligning $\beta_{fit}\left( t \right)$ and $\hat{\beta}(t)$ on the last day of data $T$, and using the unexplained variance in the model to shape the long-term trajectory.

The first issue is straightforward as we can just intercept shift $\hat{\beta}(t)$ in log-space so that it matches $\beta_{fit}\left( t \right)$ on day$T$. This approach has two large downsides though. It makes the entire forecast model of transmission intensity very sensitive to variations in the last week of the epidemiological data and the model driver data, both of which are frequently subject to reporting lags and other data quality issues. It also ignores mid-term trends in the regression residuals which frequently encode important features of the epidemic not captured by our model drivers such as the emergence of a new, unclassified variant (resulting in a large positive residual) or a breakdown of our assumptions about homogeneous mixing in a location (resulting in a large negative residual).

Our approach then is to use a second scale factor based on the recent history of the infection-weighted regression residuals and then transition linearly from the initial intercept shift to our final scale factor. Formally, we start with the residuals of the regression

$$r\left( t \right)=E^{+}\left( t \right)\cdot ln \left( \frac{\beta_{fit}\left( t \right)}{\hat{\beta}\left( t \right)} \right)$$

We then compute a final scale factor

$$s_{f}=\frac{1}{b-a}\sum_{\tau=a}^{b} r(t-\tau)$$

And define

$$s\left( t \right)= \left\{ \begin{aligned} r\left( t \right), t\leq T \\ r\left( t \right)+\frac{s_{f}-r\left( T \right)}{w}\left( t-T \right), T<t\leq T+w \\ s_{f}, t>T+w \end{aligned} \right.$$

So that our final predicted beta, which we write simply as $\beta(t)$ can be written as

$$\beta\left( t \right)=exp(s\left( t \right))\hat{\beta}\left( t \right)$$

The window over which we average the residuals, governed by the parameters $a$ and $b$, as well as the window over which the scale factor transitions, $w$, were all initially selected by out-of-sample predictive validity testing^1^ and have subsequently been tuned as a part of our weekly estimation process to handle aberrations in the residuals introduced by variants. Their current distributions can be found in table S3.

#### Section 6.3 Forecasting IDR, IHR and IFR

In each location, we must project the IDR, IHR, and IFR associated with ancestral infections among the unvaccinated with no prior infection, from which we can derive ratios conditioned on variant-type and immune status in the forecast. We set the projected ratio to be equal to the cumulative observed ratio over the final 180 days of the past. In order to avoid disjoints at the start of the forecast period, we linearly transition from the posterior ratio on the final day of the past to the projected level over the first 60 days of the forecast.

#### Section 6.4 SEI forecast

Once we have a parametric forecast of transmission intensity $\beta(t)$ and a forecast of the IDR, IFR, and IHR, we run them through the third parameterization of the SEI model alongside forecasts of future vaccination and variant spread. The third parameterization is the “natural” parameterization of the system, in which all input parameters are elements of the right-hand side of the set of equations described in section 6. This process produces 100 forecasts of vaccine-status and variant-specific infections, cases, deaths, and hospital admissions. Comprehensive diagnostics for our forecasts of all scenarios can be found in Appendix 4.

#### Section 6.5 Limitations

There are a number of simplifications made within our modelling formulation. First, the transmission model ignores the import and export of infections from locations. This assumption generally leads to an underestimation of infections. Larger, denser locations will attract people and therefore have the potential to reintroduce local transmission of the virus. When in the middle of a surge in infections, these same locations are also likely to introduce infections into less-populous neighbouring regions. The most important aspect of this assumption, however, is related to the importation of new variants which looks much more like a stochastic process. We somewhat mitigate this effect by informing our long-range forecasts of transmission intensity using the $\beta$ residuals from the regression.

The second important simplification we make is to assume a well-mixed population. This is a persistent problem in locations with small populations. It also poses challenges in the initial stages of an outbreak in larger locations where the spatial heterogeneity of the population can have a large impact on how quickly and completely the virus takes root.

### Section 7 Sources of uncertainty

The primary source of uncertainty in our models is in the reported case, death, admission, and serology data used to produce past infections. As described in section 7.2, we account for this uncertainty by generating 100 posterior draws of infection trajectories to sample the noise in the reported data. Each draw from the distribution of infection trajectories is paired with a sample of the SEI transmission parameters described in table S3 and that uncertainty is propagated through the fit, regression, and forecast stages of the transmission model to produce 100 forecast trajectories of infections, deaths, cases, and hospital admissions.

### Section 8 Predictive validity

We have previously demonstrated our model performance in a US-specific scenarios paper^1^ and in a broader model comparison project^45^. While many of our data sources and modelling assumptions are consistent with these prior analyses, we’ve expanded our modelling framework to account for several salient features of the pandemic that were not previously present. In order to evaluate the model’s performance with these new features, we ran an out-of-sample predictive validity test by holding out eight weeks of reported case, death, and admissions data from May 5, 2022 to June 30, 2022, and using our forecasting model to fill in the period. We then compared the difference in predicted total cases, admissions, and deaths over the holdout period to those reported. This was run for 100 draws in the same ensemble framework as our normal estimation process (described in section 7.2). At the global level, we underestimated deaths by -11% (-31% - 36%), underestimated admissions by -11% (-32% - 35%) and overestimated cases by 1.0% (-31%-54%).

We evaluated model performance in all 176 countries and territories and the 206 subnational units in our model and the detailed results of the analysis can be found in appendix 5. We had notable deviations (> 40%) from reported data in North America and Western Europe where we did not capture the second small surge widely attributed to the B.A.2 subvariant of Omicron. We also significantly overestimated the Omicron surge in China, as our assumptions about the impact of the lockdown were too conservative (thought notably more in line with reality than other contemporary analyses.^46^

17	Nassereldine H, Stein C, COVID-19 Forecasting Team, Lim SS. COVID-19 vaccine effectiveness after second dose by variants and main outcomes: a systematic review and meta-analysis. (To be submitted).

18	Stein C, Nassereldine H, Sorensen, R.J.D., et al. Past SARS-CoV-2 infection protection against reinfection: a systematic review and meta-analysis. *The Lancet* (In press).

19	Rader B, White LF, Burns MR, *et al.* Mask-wearing and control of SARS-CoV-2 transmission in the USA: a cross-sectional study. *Lancet Digit Health* 2021; **3**: e148–57.

20	Mitze T, Kosfeld R, Rode J, Wälde K. Face masks considerably reduce COVID-19 cases in Germany. *Proc Natl Acad Sci* 2020; **117**: 32293–301.

21	Hale T, Angrist N, Hale AJ, *et al.* Government responses and COVID-19 deaths: Global evidence across multiple pandemic waves. *PLOS ONE* 2021; **16**: e0253116.

22	Desvars-Larrive A, Dervic E, Haug N, *et al.* A structured open dataset of government interventions in response to COVID-19. *Sci Data* 2020; **7**: 285.

23	Adolph C, Amano K, Bang-Jensen B, Fullman N, Wilkerson J. Pandemic Politics: Timing State-Level Social Distancing Responses to COVID-19. *J Health Polit Policy Law* 2021; **46**: 211–33.

24	WNTRAC. https://ibm.github.io/wntrac/ (accessed June 6, 2022).

25	Feb 10 P, 2022. State COVID-19 Data and Policy Actions. KFF. 2022; published online Feb 10. https://www.kff.org/coronavirus-covid-19/issue-brief/state-covid-19-data-and-policy-actions/ (accessed June 6, 2022).

26	Vos T, Lim SS, Abbafati C, *et al.* Global burden of 369 diseases and injuries in 204 countries and territories, 1990–2019: a systematic analysis for the Global Burden of Disease Study 2019. *The Lancet* 2020; **396**: 1204–22.

27	Troeger C, Khalil I, Blacker B, et al. Quantifying risks and interventions that have affected the burden of lower respiratory infections among children younger than 5 years: an analysis for the Global Burden of Disease Study 2017. *Lancet Infect Dis* 2020; **20**: 60–79.

28	Troeger C, Blacker B, Khalil I, et al. Estimates of the global, regional, and national morbidity, mortality, and aetiologies of lower respiratory infections in 195 countries, 1990-2016: a systematic analysis for the Global Burden of Disease Study 2016. *Lancet Infect Dis* 2018; **18**: 1191–210.

29	Khan AJ, Hussain H, Omer SB, *et al.* High incidence of childhood pneumonia at high altitudes in Pakistan: a longitudinal cohort study. *Bull World Health Organ* 2009; **87**: 193–9.

30	Pérez-Padilla R, García-Sancho C, Fernández R, Franco-Marina F, López-Gatell H, Bojórquez I. The impact of altitude on hospitalization and hospital mortality from pandemic 2009 influenza A (H1N1) virus pneumonia in Mexico. *Salud Publica Mex* 2013; **55**: 92–5.

31	Pérez-Padilla R, Franco-Marina F. The impact of altitude on mortality from tuberculosis and pneumonia. *Int J Tuberc Lung Dis Off J Int Union Tuberc Lung Dis* 2004; **8**: 1315–20.

32	Global Burden of Disease Collaborative Network. Global Burden of Disease Study 2015 (GBD 2015) Covariates 1980-2015. Seattle, WA: Institute for Health Metrics and Evaluation, 2016 http://ghdx.healthdata.org/record/ihme-data/gbd-2015-covariates-1980-2015 (accessed July 8, 2020).

33	Reitsma MB, Kendrick PJ, Ababneh E, *et al.* Spatial, temporal, and demographic patterns in prevalence of smoking tobacco use and attributable disease burden in 204 countries and territories, 1990–2019: a systematic analysis from the Global Burden of Disease Study 2019. *The Lancet* 2021.

34	Murray CJL, Aravkin AY, Zheng P, *et al.* Global burden of 87 risk factors in 204 countries and territories, 1990–2019: a systematic analysis for the Global Burden of Disease Study 2019. *The Lancet* 2020; **396**: 1223–49.

35	WorldPop. WorldPop Population Dataset. 2019; published online Oct 4. https://www.worldpop.org/project/categories?id=3 (accessed Oct 4, 2019).

36	World Pop. Population counts. 2021. https://www.worldpop.org/project/categories?id=3 (accessed July 4, 2021).

37	Wang H, Abbas KM, Abbasifard M, *et al.* Global age-sex-specific fertility, mortality, healthy life expectancy (HALE), and population estimates in 204 countries and territories, 1950–2019: a comprehensive demographic analysis for the Global Burden of Disease Study 2019. *Lancet* 2020; **396**: 1160–203.

38	Press WH, Teukolsky, Saul A, Vetterling, William T, Flannery, Brian P. Section 17.1 Runge-Kutta Method. In: Numerical Recipes: The Art of Scientific Computing, 3rd edition. Cambridge, UK ; New York: Cambridge University Press, 2007.

39	Harris CR, Millman KJ, van der Walt SJ, *et al.* Array programming with NumPy. *Nature* 2020; **585**: 357–62.

40	Lam SK, Pitrou A, Seibert S. Numba: A llvm-based python jit compiler. In: Proceedings of the Second Workshop on the LLVM Compiler Infrastructure in HPC. 2015: 1–6.

41	Collins JK, Bannick M, Zheng P, *et al.* IHME COVID-19 forecasting model. 2021; published online July 7. DOI:10.5281/zenodo.5081067.

42	Peng Zheng, Jiawei He. ihmeuw-msca/regmod. 2022; published online April 5. https://github.com/ihmeuw-msca/regmod (accessed May 23, 2022).

43	Morales JL, Nocedal J. Remark on ‘algorithm 778: L-BFGS-B: Fortran subroutines for large-scale bound constrained optimization’. *ACM Trans Math Softw* 2011; **38**: 7:1-7:4.

44	Virtanen P, Gommers R, Oliphant TE, *et al.* SciPy 1.0: fundamental algorithms for scientific computing in Python. *Nat Methods* 2020; **17**: 261–72.

45	Friedman J, Liu P, Troeger CE, *et al.* Predictive performance of international COVID-19 mortality forecasting models. *Nat Commun* 2021; **12**: 2609.

46	Cai J, Deng X, Yang J, *et al.* Modeling transmission of SARS-CoV-2 Omicron in China. *Nat Med* 2022; : 1–1.

47	Faria NR, Mellan TA, Whittaker C, *et al.* Genomics and epidemiology of the P.1 SARS-CoV-2 lineage in Manaus, Brazil. *Science* 2021; **372**: 815–21.

48	Pearson C, Russell T, Davies N, et al. Estimates of severity and transmissibility of novel SARS-CoV-2 variant 501Y.V2 in South Africa. *CMMID Repos Peer Rev* 2021; published online Jan 11.

49	Prete CA, Buss LF, Buccheri R, *et al.* Reinfection by the SARS-CoV-2 Gamma variant in blood donors in Manaus, Brazil. *BMC Infect Dis* 2022; **22**: 127.

50	Allen H, Vusirikala A, Flannagan J, et al. Increased household transmission of COVID-19 cases associated with SARS-CoV-2 Variant of Concern B.1.617.2: a national case-control study. London, UK: National Infection Service, Public Health England, 2021.

51	Challen R, Brooks-Pollock E, Read JM, Dyson L, Tsaneva-Atanasova K, Danon L. Risk of mortality in patients infected with SARS-CoV-2 variant of concern 202012/1: matched cohort study. *BMJ* 2021; **372**: n579.

52	Davies NG, Abbott S, Barnard RC, *et al.* Estimated transmissibility and impact of SARS-CoV-2 lineage B.1.1.7 in England. *Science* 2021; **372**: eabg3055.

53	Fort H. A very simple model to account for the rapid rise of the alpha variant of SARS-CoV-2 in several countries and the world. *Virus Res* 2021; **304**: 198531.

54	Leung K, Shum MH, Leung GM, Lam TT, Wu JT. Early transmissibility assessment of the N501Y mutant strains of SARS-CoV-2 in the United Kingdom, October to November 2020. *Eurosurveillance* 2021; **26**: 2002106.

55	Kim MS, Seong D, Li H, *et al.* Comparative effectiveness of N95, surgical or medical, and non-medical facemasks in protection against respiratory virus infection: A systematic review and network meta-analysis. *Rev Med Virol* 2022; **32**: e2336.

56	Abaluck J, Kwong LH, Styczynski A, *et al.* Impact of community masking on COVID-19: A cluster-randomized trial in Bangladesh. *Science* 2022; **375**: eabi9069.

### Section 10 Supplementary Tables

Table S2 Vaccine efficacy by coronavirus variant, available, data, and modelled estimates

Prevention of severe disease can be interpreted as the prevention of hospitalisation and death.

| Vaccine | Ancestral and Alpha variant | | Beta, Gamma, and Delta variants | | Omicron variant | |
| --- | --- | --- | --- | --- | --- | --- |
|  | Efficacy at preventing severe disease | Efficacy at preventing infection | Efficacy at preventing severe disease | Efficacy at preventing infection | Efficacy at preventing severe disease | Efficacy at preventing infection |
| AstraZeneca | 94% | 63% | 94% | 69% | 71% | 36% |
| Tianjin CanSino | 66% | 62% | 64% | 61% | 48% | 32% |
| CoronaVac | 50% | 47% | 49% | 46% | 37% | 24% |
| Covaxin | 78% | 73% | 76% | 72% | 57% | 38% |
| Johnson & Johnson | 86% | 72% | 76% | 64% | 57% | 33% |
| Moderna | 97% | 92% | 97% | 91% | 73% | 48% |
| Novavax | 89% | 83% | 86% | 82% | 65% | 43% |
| Pfizer/BioNTech | 95% | 86% | 95% | 84% | 72% | 44% |
| Sinopharm | 73% | 68% | 71% | 67% | 53% | 35% |
| Sputnik-V | 92% | 86% | 89% | 85% | 67% | 44% |
| Other mRNA vaccines | 91% | 86% | 88% | 85% | 67% | 45% |
| All other vaccines | 75% | 70% | 73% | 69% | 55% | 36% |

Table S3 SEI transmission model parameter distributions.

| Parameter | Value | Source | Description |
| --- | --- | --- | --- |
| $\tau_{C}$ | U[10, 14] | Sensitivity Analysis^2^ | Time from exposure to a reported case. |
| $\tau_{H}$ | U[10, 14] | Sensitivity Analysis^2^ | Time from exposure to a hospital admission. |
| $\tau_{D}$ | U[12, 16] | Sensitivity Analysis^2^ | Time from hospital admission to death. |
| $\kappa_{x}^{I}$ | 1.0 - ancestral  U[1.2, 1.6] – Alpha  U[1.0, 1.4] – Beta  U[1.0, 1.4] – Gamma  U[1.6, 2.0] – Delta  U[2.5, 3.5] – Omicron | Literature^47–50^ and model calibration | Increase in transmission intensity relative to the ancestral variant |
| $\phi_{x,y}$ | 1.0 – unless otherwise noted  U[0.7, 0.9] – Beta, Gamma, and Delta vs. Alpha or ancestral  U[0.4, 0.6] – Omicron vs all other variants. | Literature^47–49^ and model calibration | Maximum cross-variant immunity |
| $\kappa_{x}^{H}, \kappa_{x}^{D}$ | 1.64 (95% CI 1.32-2.04) | Literature^51^ | Increase in risk of hospitalization and death for all non-Omicron variants. |
| $\kappa_{o}^{H}$ | 20% of Delta variant | Model calibration | Reduction in the risk of hospitalization due to an Omicron infection relative to the risk of hospitalization with a Delta infection. |
| $\kappa_{o}^{D}$ | 10% of Delta variant | Model calibration | Reduction in the risk of death due to an Omicron infection relative to the risk of death with a Delta infection. |
| $\alpha$ | $U\left[ 0.9, 1.0 \right]$ | OOS Fitting^1^ | Adjustment to mitigate well-mixed assumption. |
| $\sigma_{x}$ | $U[1/4, 1]$ – Omicron  $U[1/5,1/3]$ – All other variants | Literature^1^ | Reciprocal of the incubation period. |
| $\gamma$ | 0.5 | Literature^1,52–54^ | Reciprocal of the infectious period. |
| $\pi$ | $U\left[ 0.01, 0.1 \right]$ | Assumed | Proportion of non-escape exposed $E$ introduced into escape exposed $\tilde{E}$ on the day an escape variant invades a location. |
| $a$ | $U[1, 7]$ | OOS Fitting^1^ | Lower bound of residual averaging window in the adjustments to predicted transmission intensity. |
| $b$ | $U\left[ a+21, 42 \right]$ | OOS Fitting^1^ | Upper bound of residual averaging window in the adjustments to predicted transmission intensity. |
| $w$ | $42$ | OOS Fitting^1^ | Window size for the application of the shift from the transmission intensity on the final reported day to the residual average. |

Table S4 SEI Regression priors and coefficients

| Covariate | Constraints | Prior (mean, sd) | Result (mean, sd) | Note |
| --- | --- | --- | --- | --- |
| Intercept | N/A | N/A | Location-specific | Random effect by location |
| Pneumonia Seasonality (Section 5.4) | $[0.0, +\infty)$ | N/A | (0.17, 0.13) | Time-varying |
| Mandate Index  (Section 5.7) | $(-\infty, 0.0]$ | N/A | (-0.25, 0.22) | Time-varying |
| Mask Usage (Section 5.6) | $(-\infty, 0.0]$ | (-0.61, 0.01) | (-0.35, 0.09) | Time-varying; Prior based on literature^55,56^ |
| Air Pollution (P.M. 2.5)  (Section 5.8.4) | $[0.0, +\infty)$ | N/A | (7.1e-5, 4.8e-4) | Time-invariant |
| Smoking Prevalence  (Section 5.8.3) | $[0.0, +\infty)$ | N/A | (1.6e-3, 7.2e-3) | Time-invariant |
| LRI Mortality  (Section 5.8.1) | $[0.0, +\infty)$ | N/A | (3.6e-5, 1.8e-4) | Time-invariant |
| Altitude  (Section 5.8.2) | $[0.0, +\infty)$ | N/A | (6.9e-4, 1.6e-3) | Time-invariant |
| Population Density  (Section 5.8.5) | $\left[ 0.0, +\infty\right)$ | N/A | (5.0e-6, 2.1e-5) | Time-invariant |

### Section 11 Supplementary Figures

Figure S1. SEI model accounting for multiple vaccination rounds and SARS-CoV-2 variants

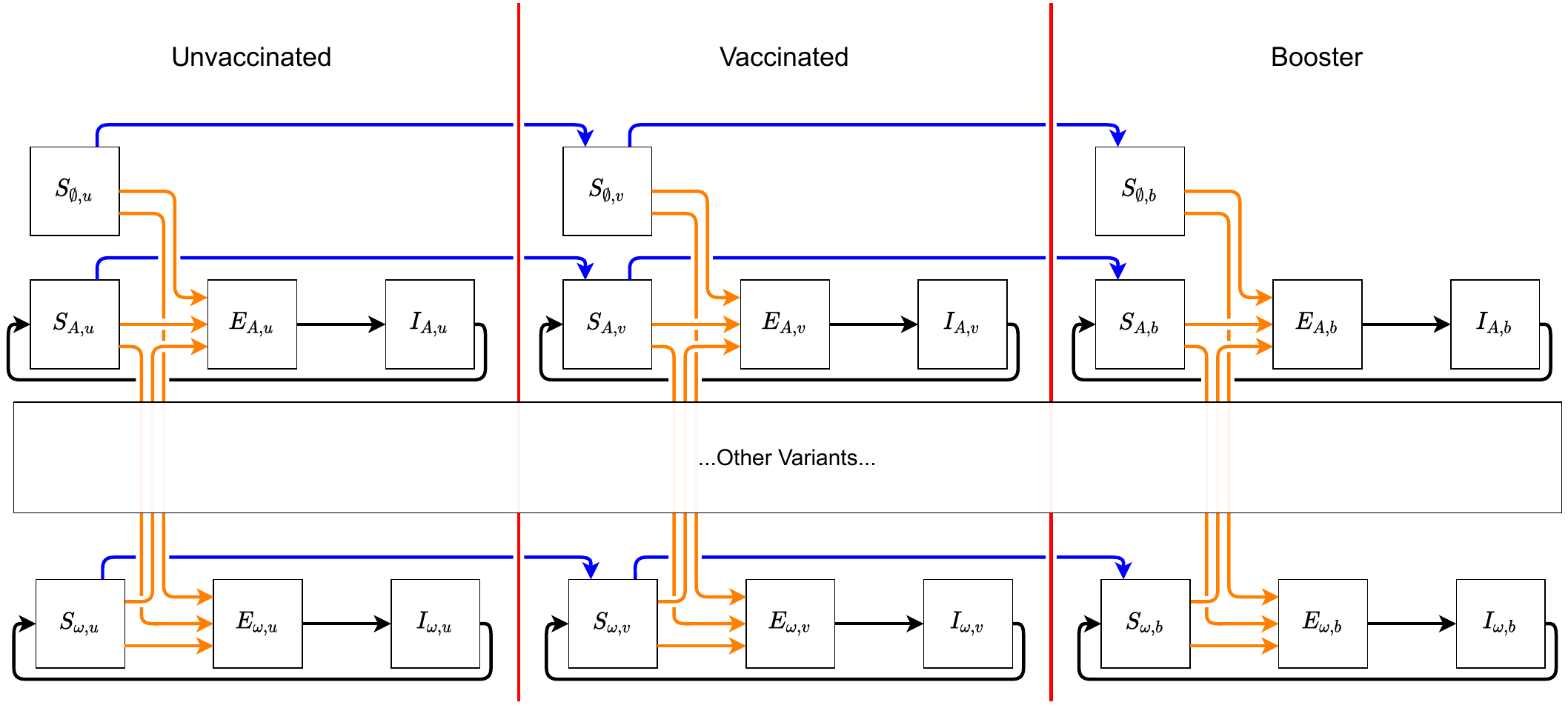

Figure S2. Total COVID-19 hospital admissions, June 1, 2022, through November 30, 2022, across four intervention scenarios, globally and by WHO region
